# Dietary staple type, protein quality and child wasting and stunting across 127 LMICs — an ecological cross-sectional study

**DOI:** 10.64898/2026.07.13.26357913

**Authors:** Rama Krishna Sanjeev, Bindu Krishnan, Reka Karuppusami, T Madhusudan, Dhasarathi Kumar, Junaid Khan, Alex Joseph, Mohan Balakrishnan, Jacques L Tamuzi

## Abstract

**Background:** Child stunting and wasting affect millions globally, with unexplained heterogeneity across LMICs. Staple foods — rice, wheat, maize, sorghum, millet and cassava — differ in protein quality (DIAAS) and zinc bioavailability. We hypothesised that these differences explain heterogeneity in undernutrition prevalence.

**Methods:** This ecological cross-sectional analysis included 127 LMICs (wasting) and 126 LMICs (stunting) using JME 2025 anthropometric data, GBD 2023 estimates and FAOSTAT 2019–2023 five-year mean food balances. Each staple was analysed individually using robust MM-estimation (primary) and quantile regression (secondary), with fixed covariates selected after circularity, correlation and multicollinearity assessment. Five sensitivity analyses tested robustness. Two secondary cross-reference analyses — WHZ, MUAC and oedema across 46 nationally representative surveys and undernutrition in infants under 6 months across 56 DHS datasets — were cross-referenced with dominant dietary staple supply. A post-hoc cross-reference analysis examined maternal short stature and low BMI by dominant dietary staple using DHS data.

**Findings:** Sorghum (β=0·361, p<0·001) and rice (β=0·086, p=0·004) were positively associated with wasting while Maize was negatively associated (β=−0·139, p=0·004). Cassava showed a positive, borderline association with stunting (β=0·975, p=0·092). HIV showed a negative wasting association reversing on exclusion of high-HIV countries. Secondary analyses confirmed MUAC-predominant wasting in cassava- and maize-dominant countries and WHZ-predominant wasting in sorghum- and millet-dominant countries.

**Interpretation:** The above findings are consistent with staple-specific growth phenotypes. Currently DHS and MICS surveys collect height and weight only — incorporating MUAC and individual staple dietary data appears imperative. Preventive measures may differ by staple context.

**Funding:** No funding was received.

**Research in context:** *Evidence before this study:* We searched PubMed using the terms “dietary staple”, “protein quality”, “DIAAS”, “child wasting”, “child stunting”, “ecological”, “epidemiology” and combinations thereof, with no language or date restrictions. Frongillo et al (1997) examined country-level determinants of stunting and wasting using energy availability as the only dietary variable, identifying unexplained residuals for India and Bangladesh. Martorell and Young (2012) documented higher wasting in India than Guatemala despite similar stunting, pointing to maternal nutritional factors. A research prioritisation exercise involving 146 global nutrition experts (Frison et al, 2020) identified ten priority areas for wasting prevention; dietary protein quality featured in none of them. Garenne(2009) identified two populations in Niakhar(Senegal) and Bwamanda(DRC) consuming staples Sorghum/millet and Cassava/maize respectively having vastly different wasting profiles. Specifically they had higher Mid-upper-arm-circumference(MUAC) based wasting in Bwamanda and lower Weight for height Z(WHZ) scores in Niakhar with differing age profiles of malnourished children. No prior ecological study has examined whether the specific type of dietary staple — and its protein quality and zinc bioavailability profile — is associated with child wasting or stunting prevalence and differing anthropometric profiles across a representative sample of LMICs.

*Added value of this study:* This is the first ecological analysis to examine the independent associations of six individual dietary staples — wheat, rice, maize, millet, sorghum and cassava — with child wasting and stunting prevalence across 127 LMICs (126 for stunting), using robust MM-estimation, quantile regression and five pre-specified sensitivity analyses. Sorghum and rice protein supply independently predicted higher wasting; maize independently predicted lower wasting. Cassava showed a positive, borderline association with stunting (β=0·975, p=0·092). These findings are consistent with each staple driving a distinct growth phenotype determined by its protein quality (DIAAS) and zinc bioavailability profile, explaining the long-documented geographic dissociation between wasting and stunting that conventional socioeconomic and disease models have failed to account for. We have additionally integrated two large datasets of child wasting by MUAC and WHZ as well as wasting and stunting (WaSt) prevalence across countries, with our dataset, to demonstrate these differences between staples. Cross-referencing the Grellety-Golden dataset (46 surveys) with dominant dietary staple showed MUAC-predominant wasting in cassava- and maize-dominant countries and WHZ-predominant wasting in sorghum- and millet-dominant countries (Kruskal-Wallis p=0·005). Cross-referencing the Kerac et al dataset (56 DHS surveys) showed that WaSt prevalence in infants under 6 months was more than double in sorghum- and millet-dominant countries compared with cassava- and maize-dominant countries, suggesting these staple-specific growth phenotypes are established from the earliest weeks of life. We also undertook a post-hoc analysis cross-referencing maternal short stature and low BMI with staple type, establishing a statistically significant negative relationship of maize with maternal low BMI, corroborating our primary robust MM-regression finding of a negative association of maize with child wasting.

*Implications for all the available evidence:* These findings have direct implications for global nutritional surveillance — DHS and MICS surveys aggregate all cereals and all roots and tubers into single food groups, precluding individual-level testing of the staple-specific protein quality hypothesis documented here. Reclassification of dietary assessment modules to capture individual staple types and preparation methods is a research priority. The use of WHZ as the sole wasting metric introduces systematic bias — height in the denominator may overestimate wasting prevalence in sorghum- and millet-dominant Sahelian populations, while underestimating it in maize- and cassava-dominant populations where linear growth deficits artificially elevate WHZ. Incorporating MUAC alongside WHZ in surveillance and programme assessment is essential to capture the full burden of wasting across differing staple-linked growth phenotypes. For policy, targeted pulse or micronutrient supplementation in cereal or tuber dominant populations could help address undernutrition.

## Introduction

Globally, stunted and wasted children numbered 150·2 million and 42·8 million respectively.^1^ Victora, analysing WHO Global Nutritional Status data across multiple world regions, documented wide variation in the proportion of child wasting at comparable levels of stunting.^2^ This was reiterated in the landmark Lancet 2008 series on maternal and child undernutrition, which noted that countries with similar stunting levels can have several-fold differences in the prevalence of severe wasting, and additionally reported a higher prevalence of low birthweight and intrauterine growth restriction in South-Central Asia and Africa.^3^ A later analysis by Martorell and Young, comparing India and Guatemala using WHO 2006 growth standards, highlighted similar stunting levels in both countries contrasting with higher child wasting, lower BMI and higher anaemia among Indian mothers.^4^ The higher prevalence of wasting in early infancy in India, alongside these maternal nutritional indicators, pointed to a role for maternal nutritional factors in determining both intrauterine growth and early postnatal lean mass accretion.^4^

Frongillo et al examined these differences at country level through socioeconomic, geographic and political lenses, with dietary factors limited to energy availability, concluding that stunting and wasting may have partly distinct aetiologies — wasting in India being higher than predicted while Bangladesh showed lower-than-expected wasting despite comparable poverty.^5^ A research prioritisation exercise involving 146 global nutrition experts identified ten priority areas for wasting prevention; dietary protein quality featured in none of them.^6^ Dietary zinc deficiency, linked to staple phytate content through food balance sheet analysis, has an established independent association with child stunting at country level.^7^ A published peer review of the subnational analysis that preceded this study noted that ‘the field of nutrition and child anthropometrics has ironically not focused as much on food supply as on other factors.’^8^

Within India this dissociation between stunting and wasting prevalence is replicated sub-nationally — peninsular states with higher SDI values, including Maharashtra, Gujarat and Karnataka, show paradoxically higher wasting than lower-SDI northern states such as Bihar and Uttar Pradesh.^9^ A plausible dietary explanation lies in the relatively greater consumption of sorghum and millet in peninsular rural populations compared with the rice- and wheat-dependent north.^10^ These staple cereals and tubers vary markedly in protein quality — as measured by the Digestible Indispensable Amino Acid Score (DIAAS), the FAO-recommended metric for protein adequacy — with sorghum having among the lowest of any major staple and cassava providing negligible protein, yet comprehensive young child DIAAS determinations remain unavailable for many staples — itself a critical evidence gap.^11,12^

Garenne et al compared two distinct African populations for types of severe wasting; sorghum and millet consuming Niakhar in Senegal as well as maize and cassava consuming Bwamanda in Democratic Republic of Congo(DRC).^13^ Niakhar had higher severe wasting with WHZ deficit (WHZ<−2·0: 16% vs 12% in Bwamanda), while higher MUAC deficit and oedema was found in Bwamanda (MUAC<125mm: 29% vs 9% in Niakhar). Strikingly, despite Bwamanda children being lighter overall (WAZ<−2·0: 46% vs 32% in Niakhar) and far more stunted (HAZ<−2·0: 51% vs 24%), their WHZ deficit was lower than Niakhar’s — because their short stature (high HAZ deficit) attenuates the weight-for-height denominator, partially compensating the weight deficit in the WHZ calculation and masking true lean mass depletion. This directly illustrates the height denominator mechanism further discussed herein. In high-stunting cassava- and maize-dominant populations, WHZ systematically underestimates true wasting burden. Only staple food differences and differing adult population body shape and stature distinguished the two populations. Grellety and Golden analysed patterns of child undernutrition across a 46 country dataset to document differing prevalences of either MUAC or WHZ predominant or both type of wasting among them.^14^ Similarly, Kerac et al have analysed prevalence of stunting, wasting or underweight in under 6 month infants in a 56 country dataset.^15^

We hypothesise that differences in dietary staple type — reflecting wide variation in protein quantity, quality and zinc availability — may explain the substantial heterogeneity in child wasting and stunting prevalence observed across LMICs. We test this hypothesis through an ecological analysis of 127 LMICs (126 for stunting) linking FAOSTAT food balance sheet data to GBD 2023 estimates and Joint Malnutrition Estimates(JME) 2025 anthropometric surveillance data. Since global JME wasting estimates are defined exclusively by WHZ and the distinct phenotypic expression of undernutrition by dietary staple type necessitates assessment beyond this single metric.We cross-referenced our findings with the Grellety-Golden and Kerac et al datasets incorporating MUAC, oedema and growth in early infancy. We also examined associations between staple type and maternal anthropometry — maternal short stature and low BMI — by post-hoc analysis using DHS data.

## Methods

### Study design

This study is an ecological cross-sectional analysis, reported in accordance with the STROBE guidelines (S Table 1), of 132 eligible low- and middle-income countries (LMICs) defined by the OECD Development Assistance Committee list of Official Development Assistance recipients (S Table 4), of which 127 provided complete wasting data and 126 stunting data.^16,17^

## Data sources

### Dependent variables

Child stunting and wasting prevalence from the Joint Malnutrition Estimates 2025 (JME 2025) were used as dependent variables; the most recent unmodelled survey estimate was preferred to avoid circularity with GBD-derived covariates.^1^

### Care and disease covariates

Covariates were grouped into dietary, care and disease categories per the UNICEF framework.^18^ Disease covariates were under-5 prevalence of lower respiratory infections, diarrhoeal diseases, malaria and HIV/AIDS (GBD 2023, GHDx).^19^ Care covariates were the Socio-demographic Index and UHC effective coverage index (GBD 2023). Healthcare Access and Quality index, WASH summary exposure values, tuberculosis prevalence and intestinal nematode prevalence were excluded on circularity grounds (S Table 8).

### Dietary variables

Protein supply (g/capita/day) for wheat, rice, maize, millet, sorghum and cassava was extracted from FAOSTAT food balance sheets as five-year means for 2019–2023 to better match anthropometric outcomes of under-5 undernutrition and counterbalance year-on-year fluctuations.^20,21^ Animal protein was summed across 14 food categories, total energy availability in kcal/capita/day and non-cereal plant protein were also extracted. Animal protein and total energy were included as fixed covariates to control for dietary context; non-cereal plant protein was excluded (Spearman r<0·10 with both outcomes; S Table 8). The biological rationale linking each staple to child growth outcomes — including published DIAAS values (range 29 for sorghum to 68 for pearl millet) and phytate:zinc molar ratios across staples — is documented in S Section E (S Tables 19–20), drawing on the published protein quality and zinc bioavailability literature.

### Covariate selection

Selection followed a five-step process (S Figure 1; S Tables 7 & 8): circularity assessment; theoretical framework; Spearman correlation with each outcome; inter-variable correlation; and Variance Inflation Factor assessment (threshold 5). SDI was retained in the stunting model only; non-cereal plant protein was excluded (Spearman r<0·10 with both outcomes).

### Analytical framework

The conceptualisation of study variables is detailed in S Section B. One cereal or tuber per model with separate fixed covariate sets for stunting and wasting were used to improve the events-per-variable ratio, avoid inter-staple collinearity, and isolate each staple’s independent contribution. The wasting model included UHC effective coverage index, LRI prevalence, diarrhoea prevalence, HIV prevalence, animal protein supply and total energy availability. The stunting model additionally included SDI and malaria prevalence, and excluded HIV prevalence on collinearity grounds (Spearman HIV–SDI r=−0·567; S Table 8). Six models were run per outcome. Primary robust MM-estimation was supplemented by quantile regression, and five sensitivity analyses tested robustness to extreme observations, data quality, geographic restriction, disease confounding and differential country population size.

### Statistical analysis

Five sensitivity analyses were conducted: S1 excluded the five highest-wasting countries; S2 excluded 11 countries with unavailable or invalid FAOSTAT data (confirmed FAO Rome, personal communication 2025); S3 was restricted to Sub-Saharan Africa; S4 excluded countries with child HIV above the 75th percentile; and S5 applied population weighting using UN IGME 2023 under-5 estimates.

Robust MM-estimation (robustbase package, KS2014 settings) was the primary method, combining a high-breakdown-point S-estimate with a high-efficiency M-estimate.^22^ Quantile regression (Q_0·25_, Q_0·50_, Q_0·75_; 1000 xy-bootstrap replications) was the secondary method. All analyses used R version 4·5·3.^23^

A p-value below 0·05 was considered statistically significant and below 0·10 was considered borderline significant, in keeping with the ecological and hypothesis-generating objectives of the analysis.

Geographic visualisations were produced using ggplot2, sf, rnaturalearth and viridis packages in R (S Figures 1–11).

Two secondary cross-reference analyses are reported in S Section F: WHZ-MUAC discordance across 46 countries (Grellety and Golden, BMC Nutr 2016) and WaSt in infants under 6 months across 56 countries (Kerac et al, BMJ Global Health 2025), both cross-referenced with dominant dietary staple protein supply from the present dataset. One post-hoc cross-reference analysis examined associations between staple type and maternal anthropometry (adult short stature and low BMI) using DHS StatCompiler data across 57 countries (S Section F.3).^14,15^

### Ethics

This study involved secondary analysis of publicly available, aggregated data from global databases. No individual-level or identifiable data were used, and ethical approval was not required.

## Results

Descriptive statistics of the study dependent and independent variables are provided in Table 1; full distributional statistics including normality assessment are provided in S Table 6. The results of primary robust-MM estimation are provided in Table 2. The secondary quantile regression and sensitivity analysis results are provided in S Tables 10–12. The top ten countries by staple protein supply, with corresponding maps, are presented in S Tables 13–18 and S Figures 1–11. Results of the secondary cross-reference analyses against the Grellety & Golden and Kerac et al datasets, together with a summary of the nutritional profile and anthropometric associations across all six staples, are presented in S Tables 21– 24 and S Figures 13–15.

**Table 1.**
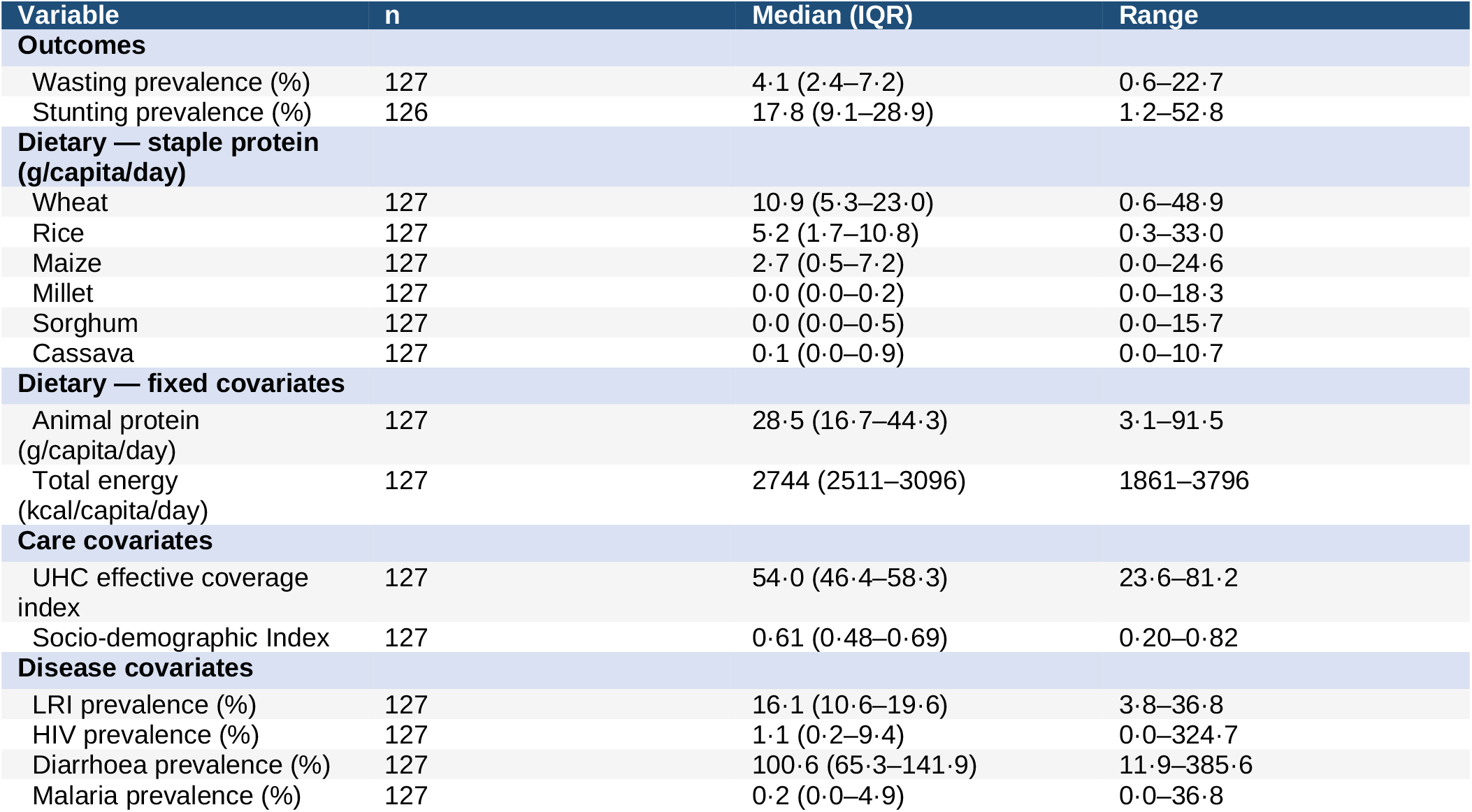
Descriptive statistics for the analytical sample (127 LMICs for wasting, 126 for stunting). Values are median (IQR) unless otherwise stated. LRI=lower respiratory infection. Wasting and stunting from JME 2025; disease covariates from GBD 2023; dietary variables from FAOSTAT 2019–2023.

**Table 2.** Primary robust MM-estimation results — association between dietary staple protein supply and child wasting (Panel A, n=127 LMICs) and stunting (Panel B, n=126 LMICs). Non-consuming countries are coded as zero supply for each staple. β = unstandardised regression coefficient (change in outcome prevalence per 1 g/capita/day increase in staple protein supply). 95% CIs calculated as β ± 1·96 × SE; full results including all covariate coefficients and standard errors are in S Table 10. Each model includes fixed covariates (see Methods). *p<0·05; **p<0·01; ***p<0·001; †p<0·10. Model R^2^ is shown for each panel.

| Staple model | $\beta$ (MM-estimate) | 95% Confidence interval | p-value |
| --- | --- | --- | --- |
| Panel A. Child wasting |  |  |  |
| Wheat | −0.028 | −0.072 to 0.017 | 0.222 |
| Rice | 0.086 | 0.029 to 0.142 | 0.004 ** |
| Maize | −0.139 | −0.232 to −0.046 | 0.004 ** |
| Millet | 0.190 | −0.016 to 0.396 | 0.073 † |
| Sorghum | 0.361 | 0.165 to 0.557 | <0.001 *** |
| Cassava | 0.039 | −0.274 to 0.351 | 0.809 |
| Model R<sup>2</sup> | 0.350–0.406 | Range across six staple models |  |
| Panel B. Child stunting |  |  |  |
| Wheat | −0.040 | −0.170 to 0.091 | 0.553 |
| Rice | −0.075 | −0.240 to 0.091 | 0.379 |
| Maize | 0.124 | −0.127 to 0.376 | 0.333 |
| Millet | −0.025 | −0.695 to 0.646 | 0.943 |
| Sorghum | 0.249 | −0.298 to 0.797 | 0.374 |
| Cassava | 0.975 | −0.149 to 2.098 | 0.092 † |
| Model R<sup>2</sup> | 0.699–0.709 | Range across six staple models |  |

### Study population

The analytical sample comprised 127 countries for wasting and 126 for stunting, after exclusion of Somalia, the Democratic People’s Republic of Korea and the Palestinian Territories (FAOSTAT food balance sheet data unavailable for all three; Somalia’s most recent JME survey also dates to 2009, a 15-year temporal mismatch); full details of country exclusions are provided in S Table 5. Stunting ranged from 1·2% to 52·8% and wasting from 0·6% to 22·7% across included countries.

#### Primary analysis — wasting

#### Staple protein associations

Sorghum and rice protein supply showed statistically significant positive associations with wasting (sorghum β=0·361, p<0·001; rice β=0·086, p=0·004), with the highest model R^2^ for sorghum at 0·406. Maize showed a significant negative association (β=−0·139, p=0·004). Millet showed a borderline positive association (β=0·190, p=0·073). Wheat and cassava were not significantly associated with wasting. Model R^2^ ranged from 0·350 to 0·406 across staple models (Table 2).

#### Fixed covariates — wasting

UHC index showed a consistent negative association with wasting across all staple models. HIV prevalence showed a biologically implausible negative association (primary β=−2·093 to −2·906, all p≤0·012), reversing to positive in S4 (β=7·6 to 12·3, all p=0·34–0·56), confirming geographic confounding (S Section D, Panel E). The country-level composition underlying this reversal is detailed in S Table 12, Panel F.

#### Primary analysis — stunting

No staple protein supply variable reached conventional significance for stunting in the primary analysis, though cassava showed a borderline positive association (β=0·975, p=0·092†) with the highest model fit (R^2^=0·709), consistent with its near-zero protein content driving chronic energy-protein deficit preferentially affecting linear growth. Model R^2^ ranged from 0·699 (millet) to 0·709 (cassava) across the six staple models.

LRI prevalence was the dominant predictor of stunting, with a significant positive coefficient across all six staple models in the primary analysis (β=63·4 to 67·1, all p<0·001; S Table 12, Panel D).

#### Quantile regression — wasting

Quantile regression broadly confirmed the primary findings. Maize showed a significant negative association with wasting across all three quantiles (Q_0·25_, Q_0·50_, Q_0·75_; all p<0·05), with the magnitude of the association increasing at higher quantiles (β=−0·109 to −0·259). Rice showed a significant positive association at Q_0·25_ only (β=0·075, p=0·002), with non-significant positive associations at Q_0·50_ and Q_0·75_. Sorghum showed a positive association that increased in magnitude across quantiles (β=0·265 to 0·546), reaching borderline significance at Q_0·50_ and Q_0·75_. Cassava showed a borderline positive association at Q_0·25_ only (β=0·291, p=0·084†). Wheat and millet showed no significant associations at any quantile (S Table 11, Panel A).

For stunting, no staple protein supply variable reached conventional significance at any quantile. Maize showed a borderline positive association at Q_0·25_ only (β=0·362, p=0·054†), consistent in direction with the primary analysis. Cassava showed consistently positive, increasing-magnitude but non-significant coefficients across all three quantiles (β=0·884 to 1·455). Wheat, rice, millet and sorghum showed no significant associations at any quantile (S Table 11, Panel B).

Quantile regression of fixed covariates for wasting (S Table 12, Panel A) showed LRI prevalence positively associated across most staples, strongest at Q_0·50_ (e.g. wheat β=15·125, p=0·006; cassava β=13·712, p=0·015) and attenuating or reversing at Q_0·75_. The UHC index showed a consistent negative association across all staples and quantiles, while HIV prevalence remained negative but non-significant throughout, mirroring the primary analysis. Animal protein supply showed a consistent negative association, reaching significance at Q_0·25_ for maize (β=−0·049, p=0·003) and wheat (β=−0·039, p=0·020).

For stunting (S Table 12, Panel B), LRI prevalence was strongly and consistently positively associated with stunting across all six staple models and all three quantiles, with magnitude increasing from Q_0·25_ to Q_0·75_ (e.g. wheat β=56·218 to 94·353; cassava β=40·271 to 69·4), confirming LRI as the dominant covariate identified in the primary analysis. The Socio-demographic Index showed a consistent negative association, significant for most staples at Q_0·25_ and Q_0·50_. Total energy supply showed a small but consistently negative association that strengthened to reach significance at Q_0·75_ for all six staples. Animal protein supply showed no consistent pattern and was non-significant throughout.

#### Sensitivity analyses

All five sensitivity analyses used robust MM-estimation. Full results are in S Table 12, Panels A–E. S1 (excl. top 5 wasting outliers, n=122/121): sorghum-wasting weakened but remained significant (β=0·221, p=0·038); rice (β=0·092, p=0·001) and maize (β=−0·128, p=0·005) remained significant; millet-wasting newly reached borderline significance (β=0·191, p=0·055); cassava-stunting remained significant (β=1·168, p=0·035).

S2 (excl. countries with invalid FAOSTAT data, n=118/117): rice strengthened (β=0·103, p<0·001); maize was essentially unchanged (β=−0·137, p=0·004); sorghum weakened but remained significant (β=0·217, p=0·046); millet-wasting newly reached borderline significance (β=0·194, p=0·056); cassava-stunting reached borderline significance (β=0·984, p=0·089†).

S3 (sub-Saharan Africa only, n=47/47): sorghum-wasting strengthened (β=0·404, p=0·008); wheat showed a sign reversal from the primary analysis, becoming strongly positive (β=0·344, p<0·001); two new stunting associations emerged in this Africa-restricted sample: sorghum (β=0·655, p=0·032) and rice, which reversed direction to become protective (β=−0·428, p=0·014); cassava-stunting remained non-significant (β=1·067, p=0·138).

S4 (excl. high-HIV countries, n=95/94): sorghum-wasting strengthened to p<0·001 (β=0·462, p<0·001); rice (β=0·068, p=0·062) and maize (β=−0·130, p=0·069) remained attenuated to borderline significance; millet-wasting newly reached significance (β=0·287, p=0·044), and sorghum-stunting newly reached borderline significance (β=0·606, p=0·071); the HIV coefficient reversed from negative to positive (though non-significant), consistent with geographic confounding (S Table 12, Panel E).

S5 (population-weighted, n=127/126): maize (β=−0·131, p<0·001) and rice (β=0·044, p=0·028) wasting associations were retained; sorghum-wasting was attenuated but remained highly significant (β=0·249, p<0·001). Millet-wasting and cassava-wasting newly emerged as highly significant (β=0·208, p<0·001 and β=0·278, p<0·001 respectively). For stunting, rice (β=−0·133, p=0·012, protective) and maize (β=0·195, p=0·022) were retained, while sorghum (β=0·715, p<0·001) and cassava (β=1·134, p<0·001) showed substantially stronger positive associations; most strikingly, millet-stunting reversed from a small protective association to the largest positive coefficient in the analysis (β=2·954, p<0·001).

#### LRI-stunting robustness

LRI prevalence remained a positive predictor of stunting across all five sensitivity analyses and all six staple models (β ranging from 30·2 to 77·7), reaching p<0·001 in S1, S2, S4 and S5 for all staples; in S3 (sub-Saharan Africa only, smallest sample) all staples remained significant (p<0·05) except rice (p=0·068) — the most robust covariate finding in the analysis.

#### Geographic distribution

Choropleth maps of staple protein supply and child undernutrition prevalence are presented in S Figures 1–11. The top 10 consuming countries for each staple are presented in S Tables 13–18.

### Secondary cross-reference analyses

The Grellety and Golden WHZ-MUAC discordance dataset (n=47 nationally representative surveys) showed that sorghum and millet protein supply were positively correlated with WHZ-predominant wasting (r=0·295, p=0·047 and r=0·383, p=0·009 respectively), while cassava protein supply was negatively correlated (r=−0·295, p=0·047), indicating MUAC-predominant wasting in cassava-dominant countries.

Rice showed a positive correlation (r=0·443, p=0·002). Stunting prevalence was inversely correlated with the WHZ-MUAC difference (r=−0·463, p=0·001), consistent with the height denominator effect — countries with higher stunting tending toward MUAC-predominant wasting. In the Kerac et al dataset of 56 LMIC DHS surveys in infants under 6 months, mean WaSt was more than double in sorghum- and millet-dominant countries (1·80%) compared with maize- and cassava-dominant countries (0·67%), alongside higher mean wasting prevalence (15·1–20·8% vs 5·4–8·1%), while stunting was lower in sorghum/millet countries (11·2–14·8%) than in maize/cassava countries (19·1–22·0%). In the post-hoc maternal anthropometry analysis (n=57 countries, DHS StatCompiler), maize protein supply was negatively associated with maternal thinness (β=−0·427, p<0·001), while millet (β=0·386, p=0·096†) and sorghum (β=0·523, p=0·083†) showed positive associations; rice protein supply was positively associated with maternal short stature (β=0·101, p=0·011). Full results are presented in S Section F and S Tables 21–24.

## DISCUSSION

Our primary analysis findings indicate that among the dietary staples, both sorghum and rice protein availability are positively associated with child wasting, while millet had a borderline positive association and maize showed a consistent negative association. Cassava showed a borderline positive association with stunting (β=0·975, p=0·092). These patterns are consistent with each staple driving a characteristic growth phenotype determined by its protein quality (DIAAS) and zinc bioavailability: protein quality deficit in sorghum and millet producing lean mass depletion and wasting; maize, with a relatively higher DIAAS (36) compared with sorghum and cassava, conferring a protein quality advantage contributing to its negative wasting association; and protein quantity and zinc deficit in cassava producing linear growth impairment and stunting. These phenotypes explain the geographic dissociation between wasting and stunting prevalences documented by Victora and Frongillo — not as random variation, but as the predictable nutritional consequence of the dominant dietary staple consumed.^2,5,24^

Cassava showed a borderline positive stunting association in primary analysis, significant in S1 and S5. Its near-zero protein content and very low zinc content (S Table 20) produce chronic energy-protein deficit, causing stunting; wasting from lean mass deficit is detectable through lowered MUAC in cassava-consuming countries in our secondary analysis (S Table 21, Panels A,B,C). Cassava consumption has also been implicated in kwashiorkor in DRC, in consonance with its very low protein-energy ratio and higher oedema prevalence in cassava-consuming countries (S Figure 14).^12,25^ Commensurate with the contrasting phenotypes observed by Garenne et al (2009) across sorghum/millet- and cassava/maize-consuming populations, the two cassava-dominant countries in the Kerac cross-reference — Burundi (stunting 25·9%, WaSt 0·6%) and DRC (stunting 18·2%, WaSt 1·2%) — have substantially higher stunting and lower WaSt than sorghum/millet-dominant countries (WaSt mean 1·80%), pointing to cassava-associated stunting attenuating WHZ-measured wasting.^13^ Stunting caused by dietary deficits linked to staples such as cassava may contribute to underestimation of wasting by WHZ: a shorter height denominator in the WHZ calculation partially compensates the weight deficit, producing a less severely depressed WHZ score despite true lean mass depletion — henceforth termed the height denominator effect. This is directly illustrated by Garenne et al (2009), who documented that Bwamanda (cassava/maize-consuming, DRC) children had substantially higher stunting (HAZ<−2: 51% vs 24% in Niakhar) but lower WHZ-detected wasting (WHZ<−2: 12% vs 16%) than Niakhar (sorghum/millet-consuming, Senegal) children, despite Bwamanda children being lighter overall (mean WAZ: −1·884 vs −1·304 in Niakhar).^13^

Sorghum showed the most robust wasting association, significant across all five sensitivity analyses (Table 2 and S Table 12, Panel A), with the highest model fit (R^2^=0·406). The medium to very high wasting in the top ten sorghum-consuming countries (S Table 13) directly reflects this. The low DIAAS (~29; S Table 19), high phytate:zinc ratio (S Table 20) and lysine limitation — critical for lean tissue synthesis — are consistent with the FAO monograph noting that sorghum may be unable to meet even maintenance requirements in adults.^26^ WHZ-predominant wasting in sorghum-dominant countries in our cross-reference analysis (S Table 21, Panels A,B,C) and higher WaSt in early infancy (S Table 22) further confirm lean mass depletion as the dominant phenotype. The directionally inconsistent stunting association (negative in S1/S2, positive in S3/S5) is paradoxical given sorghum’s highest phytate:zinc ratio of all six staples (48·9; S Table 20), which might be expected to drive zinc-deficiency stunting as seen with maize. This apparent inconsistency deserves closer scrutiny and may be linked to food processing or other undefined factors. The high collinearity with the co-consumed Sahelian staple,millet (r=0·675, S Table 7), which has higher zinc content and lower phytate:zinc ratio, further destabilises the stunting signal across sensitivity specifications. The WHZ-predominant wasting pattern in sorghum-dominant countries, rather than the MUAC predominance seen in maize and cassava, is consistent with relatively preserved stature in these populations, where protein quality deficit rather than zinc-mediated linear growth impairment appears to be the dominant mechanism.^13^ Millet showed a borderline positive wasting association, reaching significance in S4 and S5, with findings broadly mirroring sorghum given their high geographic collinearity (r=0·675, S Table 7). Interpretation is constrained by FAOSTAT’s undifferentiated millet aggregate, which encompasses species with near-tenfold DIAAS variation — from proso millet (DIAAS 7) to pearl millet (DIAAS 68; S Table 19).^11^

Rice showed a consistent positive wasting association in primary analysis and three of five sensitivity analyses. Its moderate DIAAS (~47) but low protein content per unit energy means total indispensable amino acid delivery is constrained by quantity, predisposing to lean mass depletion. The consistently negative stunting direction, reaching significance in S3 and S5, probably reflects fish complementation in major rice-consuming countries — exemplified by Bangladesh, which has substantially lower wasting and stunting than neighbouring India despite a considerably lower SDI.^5,27^ Rice’s low phytate:zinc molar ratio (20·1; S Table 20) makes rice-dominant diets more amenable to nutritional complementation than sorghum- or maize-dominant diets, and WHZ-predominant wasting in the Grellety cross-reference (S Table 21) is consistent with relatively preserved stature in rice-consuming populations — lean mass loss from lower protein content is therefore more readily detectable by WHZ when height is maintained. This contrasts with Gasgruber et al.^28^ who reported an inverse association between rice consumption and height; however, that analysis compared rice specifically against wheat, whereas our analysis situates rice within a six-staple comparison spanning all major cereals and cassava. The direction of association for any single staple is therefore sensitive to the comparator set, and rice’s relatively favourable stunting association here reflects its position relative to a broader reference group rather than a contradiction of Gasgruber’s narrower wheat-comparison finding.^28^

Maize showed a consistent negative association with wasting (primary analysis, all quantile regression quantiles, and four of five sensitivity analyses) alongside a consistently positive stunting association across primary analysis and all five sensitivity analyses, reaching significance in S5 and borderline significance in S2 and at Q0·25 in quantile regression. This pattern is consistent with two complementary mechanisms: a genuine protective effect on wasting through adequate weight gain, and a height denominator effect whereby linear growth impairment attenuates the WHZ denominator, masking true lean mass depletion.

Maize’s DIAAS (36) exceeds sorghum’s (29) and cassava’s (17), though it remains lower than wheat’s (48) and rice’s (47); its higher protein quantity (9·5g/100g vs rice’s 6·7g/100g) may partially offset this shortfall relative to rice specifically. However, maize’s low zinc content (1·8mg/100g) and high phytate:zinc molar ratio (45·5, second-highest of the six staples; S Table 20) could plausibly also contribute to stunting via impaired zinc-dependent linear growth. A randomized trial testing this directly in a maize-dependent Guatemalan population found no improvement in linear growth from either low-phytate maize or zinc supplementation, arguing against zinc bioavailability as the operative mechanism.^26^

By contrast, a quality protein maize trial in Ethiopia found that children in conventional (ordinary) maize households showed significantly worsening height-for-age (HAZ −1·30 to −1·54, p=0·015) alongside simultaneous, non-significant improvement in weight-for-height (WHZ −0·32 to −0·18) — directly demonstrating the height denominator effect at the individual level.^25^

Our post-hoc analysis corroborates the wasting-protective effect, with maize protein supply negatively associated with maternal thinness (BMI<18·5; β=−0·427, p<0·001; S Table 24). As an adult outcome, this should be extrapolated to child growth mechanisms with caution, given differing nutritional requirements and more variable food quantity consumed relative to requirement in adults; however, maize’s non-significant negative coefficient with maternal short stature — the opposite direction a height-denominator artifact would predict — suggests a genuine nutritional association rather than a stature-driven artifact.

The Grellety-Golden and Kerac cross-references together provide convergent support from independent datasets.^14,15^ WHZ-MUAC discordance data across 46 surveys confirmed sorghum/millet countries show WHZ-predominant wasting while maize/cassava countries show MUAC-predominant wasting (Kruskal-Wallis H=10·5, p=0·005). WaSt was more than double in sorghum/millet-dominant than cassava/maize-dominant countries in infants under 6 months (1·80% vs 0·67%); since infants under 6 months have not yet begun complementary feeding, this differential probably reflects maternal dietary protein quality transmitted through breast milk — Dror and Allen reported that breast milk amino acid composition responds to maternal dietary intake, though breast milk zinc is largely refractory to maternal zinc intake.^29^ The post-hoc maternal anthropometry analysis (n=57, DHS StatCompiler) showed a highly significant negative association between maize supply and maternal thinness (β=−0·427, p<0·001), independently corroborating maize’s wasting-protective signal.

Among fixed covariates, UHC index showed a consistent negative association with wasting across all staple models. LRI prevalence showed a consistent positive association with stunting across all sensitivity analyses (β=30·2 to 77·8, all p<0·001), consistent with evidence that repeated LRI episodes impair linear growth through inflammatory suppression of IGF-1.^30^ This association warrants caution since GBD LRI estimates are modelled using SDI, which is itself associated with stunting; the LRI-stunting coefficient may therefore partly reflect shared SDI dependence.^31^

These findings collectively underscore three surveillance priorities: first, the inclusion of MUAC alongside WHZ as a mandatory wasting metric in DHS and MICS surveys to capture the full burden of wasting across staple-linked growth phenotypes; second, the disaggregation of the current single cereal and single roots-and-tubers food group into individual staple types in dietary assessment modules;^32,33^ and third, the separate documentation of millet species consumed, given the near-tenfold variation in DIAAS between proso-millet (DIAAS 7) and pearl millet (DIAAS 68) that is entirely obscured by aggregate FAOSTAT millet reporting. More broadly, the use of WHZ alone as the global wasting indicator in JME and SDG monitoring systematically underestimates true wasting burden in cassava- and maize-dominant populations where MUAC-identified and oedematous wasting predominate.

### Limitations

This study is an ecological analysis based on country-level food balance sheet data and is subject to the ecological fallacy — country-level associations cannot be used to generate conclusions on individual health outcomes but only to generate hypotheses requiring confirmation in individual-level studies. The inability to test these associations sub-nationally is a key limitation: DHS and MICS surveys record all cereal consumption within a single undifferentiated food group and aggregate all roots and tubers, precluding staple-specific analysis at individual level. The FAOSTAT millet variable aggregates all millet species despite substantial variation in DIAAS values (7 to 68; S Table 21). Lack of DIAAS data for key staples in infants and young children under typical preparation conditions is a further limitation. Food processing methods substantially alter amino acid and zinc bioavailability and cannot be captured in food balance sheet data. The use of WHZ as the wasting metric misses MUAC-identified and oedematous wasting, potentially underestimating true wasting burden in cassava- and maize-dominant populations. The denominator effect, wherein short stature linked to a staple’s nutritional profile as in cassava could reduce WHZ-based wasting prevalence, while an opposite effect may operate in millet-dominant Sahelian populations where better zinc-supported linear growth elevates WHZ, is a further limitation of WHZ as the sole wasting metric. Temporal mismatches between anthropometric survey years and FAOSTAT reference periods represent a residual limitation. The post-hoc maternal anthropometry cross-reference reflects adult, not child, outcomes; extrapolating maternal short stature and thinness associations to child growth mechanisms should be interpreted cautiously, since adult nutritional requirements differ from those of young children, and the quantity of food actually consumed relative to requirement is considerably more variable in adults, reflecting heterogeneous dietary access, activity levels, and intra-household food allocation. The LRI-stunting association may partly reflect shared SDI dependence as described above. Other macro- and micronutrients and genetic factors contribute to child anthropometry but have not been factored in this analysis The one-staple-per-model design, while necessary to avoid inter-staple collinearity, means that each staple coefficient reflects not only the nutritional properties of that staple but also the dietary and socioeconomic context of countries where it dominates — including the effects of co-consumed staples that are correlated with it. The millet-sorghum collinearity (r=0·675, S Table 7) is a specific example: their near-identical Sahelian geographic distribution means their individual coefficients cannot be cleanly separated. The independent contributions of co-consumed staples cannot be disaggregated without individual-level dietary data.

## Data Availability

The analytical dataset, population-weighted dataset, master food balance sheet dataset, and all R analysis and map generation code will be made available on Mendeley Data upon posting of this preprint (DOI to be assigned). Data are deidentified country-level aggregates from publicly available sources (JME 2025, GBD 2023, FAOSTAT 2019-2023) and will be freely available without restriction. Requests for data prior to formal deposit may be directed to the corresponding author at.

## Contributors

RKS conceived and designed the study, developed the hypothesis, acquired and cleaned all data, conducted all statistical analyses, interpreted the results, and wrote the manuscript. RKS directly accessed and verified the underlying data reported in the manuscript. BK contributed to hypothesis development, data collection and cleaning, interpretation of results, and review of the manuscript. RK contributed to data collection, cleaning, statistical analysis and review. DK contributed to data cleaning, statistical analysis and interpretation of results. JK contributed to statistical analysis. TM contributed to concept development, overseeing the study, and drafting the manuscript. AJ provided critical feedback and contributed to the analysis. MB contributed to statistical analysis. JLT contributed to data collection, checking of the manuscript, and review. All named authors reviewed and approved the final manuscript and accept responsibility for the decision to submit for publication.

## Data sharing

The analytical dataset (validation_dataset_final_127LMIC_corrected.csv), population-weighted dataset (validation_dataset_with_population_127LMIC_corrected.csv), master food balance sheet dataset (master_dataset_Food_balances_corrected.xlsx), and all R analysis and map generation code will be made available on Mendeley Data upon posting of the preprint (DOI to be assigned). Data are deidentified country-level aggregates from publicly available sources (JME 2025, GBD 2023, FAOSTAT 2019–2023) and will be freely available without restriction. Requests for data prior to formal deposit may be directed to the corresponding author at.

## Artificial intelligence

Claude (Anthropic, version claude-sonnet-4, 2025–26) was used to assist with language editing, grammar checking, and R code debugging during manuscript preparation. Claude was not used in study design, data analysis, interpretation of results, or generation of scientific conclusions. All analytical code was written, verified and executed by the corresponding author. All scientific interpretations and conclusions are the sole responsibility of the authors. Prompts and outputs are available on request.

## Acknowledgements and funding sources

No funding was used from any external sources for the study. We acknowledge the contribution through guidance for the paper of Dr. Nirupama Shivakumar, Prof. Hans Henrik Stein, Prof. Glenda Courtney-Martin and Prof. Mark Manary. We acknowledge the guidance of Dr Akshay Dinesh for the maps. The scientific hypothesis and core analytical concept underlying this manuscript were conceived by the corresponding author. Valuable input was received from the GBD 2021 LMIC Child Malnutrition Collaborators, which helped improve this work.

