## Supplementary Materials for "Dietary staple type, protein quality and child wasting and stunting across 127 LMICs — an ecological cross-sectional study"

**Index of Supplementary Materials**

*Page numbers ([p]) to be completed on PDF compilation.*

**S Section A — Theoretical Basis and Hypothesis**

Type 1 and Type 2 nutrients — theoretical framework	4

Underlying hypothesis — staple protein quality, zinc and child growth	6

Application to concurrent stunting and wasting (WaSt)	6

**S Section B — Conceptualisation of Study Variables**

Rationale for food balance sheet approach	8

Individual staple model specification (EPV justification)	8

**S Section C — Data Sources and Code Availability**

**Tables**

S Table 1 — STROBE checklist	12

S Table 2 — Data sources	16

S Table 3 — WHO/JME prevalence thresholds and severity classifications	17

S Table 4 — Country list: 141 LMICs (OECD DAC list 2022–2023)	17

S Table 5 — Missing data and excluded countries	19

S Table 6 — Descriptive statistics: all analytical variables	21

**S Section D — Statistical Methods and Results**

**Covariate selection**

S Table 7 — Covariate and exposure variable assessment (Panels A–C): circularity assessment; inter-variable Spearman correlation matrix; inter-staple collinearity matrix	22

S Table 8 — Covariate inclusion/exclusion — detailed rationale. The primary JME dataset captures wasting by WHZ only in children aged 6–59 months, which may underestimate true wasting where MUAC predominates and excludes infants under 6 months. Two secondary cross-reference analyses and one post-hoc cross-reference analysis address this: (1) Grellety & Golden (n=47 surveys) incorporates both WHZ and MUAC, enabling assessment of WHZ–MUAC discordance by dominant dietary staple; (2) Kerac et al (n=56 countries) captures simultaneous wasting-and-stunting specifically in infants under 6 months.	28

**Primary results**

S Table 9 — Complete robust MM regression results — primary analyses (Panels A–B: wasting, stunting). Statistical methods: (a) Robust MM-estimation (robustbase, lmrob, setting=“KS2014”); one staple per model — EPV constraints (n=127 wasting, n=126 stunting; 6–7 fixed covariates; EPV=18–21). Wasting covariates: UHC, LRI, diarrhoea, HIV, animal protein, total kcal. Stunting covariates: SDI, UHC, LRI, diarrhoea, malaria, animal protein, total kcal. (b) Quantile regression Q0·25/0·50/0·75 (quantreg, rq, se=“boot”, R=1000); wasting only. (c) Missing data: listwise deletion; FAOSTAT-absent countries filled from BJN dataset or regional mean imputation — see S Table 5. (d) N/A — ecological design; no individual-level data. (e) Five sensitivity analyses S1–S5. Secondary cross-reference: bivariate only — Spearman correlations (Grellety & Golden) and simple linear regression (Kerac et al). No correction for multiple comparisons. See S Tables 10–14.	33

S Table 10 — Quantile regression: staple protein associations (Panels A–B)	36

S Table 11 — Quantile regression: fixed covariate associations (Panels A–B)	38

**Sensitivity analyses**

S Table 12 — Sensitivity analyses S1–S5 (Panels A–E): Panel A = wasting staple associations; Panel B = stunting staple associations; Panel C = LRI robustness wasting; Panel D = LRI robustness stunting; Panel E = HIV prevalence association wasting. Theory-driven secondary cross-reference analyses (S Section F): Grellety & Golden WHZ–MUAC discordance (S Table 21, n=47 surveys) and Kerac et al WaSt in infants <6 months (n=56 countries). All secondary analyses are hypothesis-generating; no correction for multiple comparisons applied.	40

[p]

**Dietary staple distribution — S Section D.1**

S Table 13 — Top 10 countries: sorghum protein supply (FAOSTAT 2019–2023)	47

S Table 14 — Top 10 countries: millet protein supply (FAOSTAT 2019–2023)	49

S Table 15 — Top 10 countries: cassava protein supply (FAOSTAT 2019–2023)	51

S Table 16 — Top 10 countries: maize protein supply (FAOSTAT 2019–2023)	53

S Table 17 — Top 10 countries: rice protein supply (FAOSTAT 2019–2023)	55

S Table 18 — Top 10 countries: wheat protein supply (FAOSTAT 2019–2023)	56

**Maps (S Figures 1–11)**

S Figure 1 — Covariate selection process — five-step framework	22

S Figure 2 — World map: sorghum protein supply (FAOSTAT 2019–2023)	48

S Figure 3 — Africa map: sorghum protein supply (FAOSTAT 2019–2023)	49

S Figure 4 — World map: millet protein supply (FAOSTAT 2019–2023)	50

S Figure 5 — Africa map: millet protein supply (FAOSTAT 2019–2023)	51

S Figure 6 — World map: cassava protein supply (FAOSTAT 2019–2023)	52

S Figure 7 — Africa map: cassava protein supply (FAOSTAT 2019–2023)	53

S Figure 8 — World map: maize protein supply (FAOSTAT 2019–2023)	54

S Figure 9 — Africa map: maize protein supply (FAOSTAT 2019–2023)	55

S Figure 10 — World map: rice protein supply (FAOSTAT 2019–2023)	56

S Figure 11 — World map: wheat protein supply (FAOSTAT 2019–2023)	57

**S Section E — Protein Quality and Zinc Bioavailability of Dietary Staples**

**Tables**

S Table 19 — DIAAS reference values for six staples	58

S Table 20 — Zinc content and phytate:zinc molar ratios for six staples	60

**S Section F — Secondary Cross-reference Analyses**

**S Section F.1 — WHZ-MUAC discordance by dietary staple (Grellety-Golden, n=47)**

S Table 21 — WHZ-MUAC discordance by dietary staple: Spearman correlations	64

S Figure 13 — Proportion of GAM children detected by WHZ, MUAC, or both criteria, by dominant dietary staple	69

**Oedema prevalence by dominant dietary staple**

S Figure 14 — Oedema exclusion % by dominant dietary staple	69

**S Section F.2 — WaSt in infants under 6 months (Kerac et al, n=56)**

S Table 22 — Stunting, wasting and WaSt in infants <6 months by dominant staple	70

S Figure 15 — Stunting vs WaSt in infants under 6 months by dominant dietary staple	72

**S Section F.3 — Post-hoc cross-reference: Maternal short stature and thinness by dietary staple (DHS StatCompiler, n=57)**

S Table 23 — Maternal short stature and thinness by staple protein supply, adjusted for kcal and animal protein	74

S Table 24 — Summary of nutritional profile and anthropometric associations across all six staples	75

**Bibliography — References for Supplementary Sections**

### **S Section A. Theoretical basis and hypothesis**

#### ***Type 1 and Type 2 nutrients***

#### The nutrients required for growth are classified into two types.^1^ Type 1 nutrients are required for specific metabolic functions, have body stores and result in characteristic deficiency symptoms — for example, vitamin A deficiency causing night blindness and iron deficiency causing anaemia. Type 2 nutrients have distributed functions affecting growth without characteristic deficiency features or body stores. The two major type 2 nutrients are protein and zinc.^1^ Kenneth Carpenter documented that the poorest inhabitants of a region consume the narrowest range of foods dominated by a staple contributing the most calories, while exhibiting forms of malnutrition linked to that staple type and its processing.^2^ Staples, particularly cereals, form the major sources of protein for the majority of the poor in LMICs.^3^ There has been a relative neglect of protein as contributory to child undernutrition over recent decades, with the current narrative focused on micronutrients.^4^ Minimum dietary diversity (MDD), the key dietary indicator in DHS and MICS surveys, has been associated with stunting at population level^5^; a recent ecological analysis across 185 countries found higher dietary diversity scores associated with lower wasting DALYs as well as stunting DALYs.^6^ However, while management protocols for severe wasting have been successfully practised, primary aetiological factors for wasting remain poorly understood, and the research prioritisation exercise by Angood et al did not include specific questions on protein quality through staple foods as a determinant of wasting.^7^

In a study based in rural Malawi, children with stunting had all nine essential amino acids lower than non-stunted children.^8^ In a targeted metabolomic study of hospitalised children with severe acute malnutrition (SAM), metabolic profiles were profoundly different from stunted or non-stunted controls; children with kwashiorkor were metabolically distinct from those with marasmus, with most metabolites — particularly amino acids and biogenic amines — significantly lower in the former.^9^ These studies point to the need for further exploration of the role of protein and amino acid intakes in the causation of child wasting. However, none of these studies has directly linked population-level staple protein quality by cereal type to wasting prevalence across countries — the gap this ecological analysis addresses.

An ecological study by Frongillo, de Onis and Hanson showed that more of the variability in stunting than wasting was explained by national socioeconomic factors — energy availability, female literacy and GDP.^10^ In their comparison of India and Bangladesh, India had higher-than-expected wasting and Bangladesh lower-than-expected wasting after accounting for these factors, with no satisfactory explanation offered. A subnational analysis of Indian states similarly showed that peninsular states with higher SDI — Maharashtra and Karnataka — had higher wasting than lower-SDI Gangetic plain states such as Uttar Pradesh and Bihar, while the reverse held for stunting.^11^ A possible dietary explanation could be that Maharashtra and Karnataka are among the highest consumers of sorghum and millet in India — staples with lower DIAAS — while the Gangetic plain relies more on wheat and rice.^12,13^ Bangladesh's lower-than-expected wasting, despite low SDI, likely reflects widespread consumption of rice with small whole fish — mola, chanda and related small cyprinids accessible as common-pool resources to the poorest households — providing complete protein with DIAAS approaching 100 alongside bioavailable zinc and vitamin A.^14,15^ These contrasts illustrate why SDI does not function as a reliable wasting covariate at country level and was excluded from the wasting models in the present analysis. Keeping in mind these differences , in our study, we have used a different set of covariates for stunting (with inclusion of SDI) and wasting.

#### ***Underlying hypothesis***

The underlying hypothesis proceeds from unexplained heterogeneity in under-5 child stunting and wasting prevalence among LMICs. The chief source of protein for poor populations is the staple food consumed — rice, wheat, maize, millet, sorghum and cassava. If protein (including indispensable amino acids) and zinc are both adequate, stature and lean body mass should be optimal. If protein is deficient but zinc is adequate, the nutrient mix may support linear growth but not lean tissue accretion, contributing to wasting. If zinc is more deficient than protein, stunting may result. If both are substantially deficient, concurrent stunting and wasting may follow.

Higher wasting prevalence may be linked to consumption of staples with lower DIAAS or negligible protein content. Higher stunting prevalence may be linked to lower zinc bioavailability from high phytate: zinc ratios. When energy availability is inadequate, dietary protein may be catabolised for energy, contributing to wasting or stunting independently of protein quality.

Notably, earlier wasting may lead to later stunting through impaired linear growth during recovery from acute lean mass deficit.^16^ This ecological analysis uses this framework to examine associations between staple protein supply by type and heterogeneity in wasting and stunting across LMICs with low animal protein availability.

#### ***Application to concurrent stunting and wasting (WaSt)***

In a meta-analysis of WaSt from 84 countries, greater than 5% WaSt prevalence was observed in nine countries. Six were from Sub-Saharan Africa (Niger, Burundi, Djibouti, Chad, Sudan and South Sudan) and three from Asia (Timor-Leste, Yemen and India).^17^ The dietary staple patterns of these nine countries are instructive. Niger, South Sudan, Chad and Sudan have among the highest sorghum and millet consumption in the present dataset (S Tables 17–18). Sorghum presents the most adverse combined nutritional profile of the six staples — the lowest DIAAS (29, pig model, applicable to older children and adults) and the highest phytate: zinc molar ratio (48·9) (S Tables 14–15). The FAOSTAT millet variable is an undifferentiated aggregate encompassing pearl millet, finger millet, teff and other minor millets with substantially different protein quality profiles; DIAAS estimates range from 7 (proso millet, rat model) to 68 (pearl millet, IAAO method in adults), limiting interpretation. However, millet-dominant diets in the Sahel are typically consumed with minimal animal protein (Niger 10·1 g/capita/day, Mali 10·2 g/capita/day), reducing the complementation that would otherwise offset a low-quality staple protein. Burundi's dominant staple is cassava — the lowest protein density of the six staples (0·9–2·1 g/100g) — compounded by the lowest animal protein supply of the nine countries (3·1 g/capita/day) and total energy availability below 2100 kcal/capita/day; this combination of protein quantity deficit and energy deficit is consistent with its high WaSt burden. Yemen similarly has energy availability below 2100 kcal/capita/day with low animal protein supply (11·6 g/capita/day), with wheat as the dominant staple. India's national millet and sorghum averages are modest, but regional concentration of these staples in high-wasting peninsular states — Maharashtra, Karnataka and Rajasthan — is consistent with the regional WaSt pattern.^11,12^ Djibouti and Timor-Leste are less readily explained by the staple protein hypothesis; Timor-Leste is rice-dominant with the next dominant staple being maize, complemented with a relatively higher animal protein intake of 21.7 gm per capita per day, which could explain its very high stunting(46.7%) with moderate wasting(8.3%) .Djibouti is wheat-dominant with adequate energy availability (2739 kcal/capita/day),suggesting that other country-specific factors, including high infectious disease burden coupled with poor universal health coverage and lower SDI, could contribute to their WaSt prevalence. Taken together, these dietary patterns are broadly consistent with the hypothesis that concurrent stunting and wasting is linked to low protein quality and low zinc bioavailability from dominant dietary staples, with energy deficit as a compounding factor in the most severely affected countries.

### **S Section B. Conceptualisation of study variables**

#### ***Rationale for food balance sheet approach***

Individual-level data on dietary staple consumption by type are not available through DHS and MICS surveys, which classify cereals and tubers in undifferentiated generic food group categories.^18,19^ FAOSTAT food balance sheets provide the only systematic country-level data on protein supply from individual staple types across LMICs. Five-year means (2019–2023) were used to attenuate random measurement error and smooth agricultural supply variability.^20^

The combination of five-year dietary averages and under-5 outcomes substantially mitigates the cross-sectional limitation — children aged 0–59 months at survey experienced their critical first 1000 days predominantly within the 2019–2023 reference period. FAOSTAT food balance sheet data for 2019–2023 are currently unavailable and masked for data quality reasons for nine countries (Benin, Burundi, Central African Republic, Chad, Mali, Somalia, South Sudan, Sudan and Togo) and have never been compiled for two further countries for Eritrea and Equatorial Guinea (Filipczuk T, FAO Rome, personal communication, 17 November 2025)., with values imputed by us from regional means. The nine countries’ data was downloaded by us earlier of the time series, 2018-2022.These 11 countries, including Eritrea and Equatorial Guinea, are excluded from sensitivity analysis S2. Somalia is excluded from all analyses: FAOSTAT data are unavailable for 2019–2023, and the JME wasting survey year is 2009 — a 15-year temporal mismatch and the oldest survey in the dataset. DPR Korea was also excluded from all analysis, as data was unavailable. The analytical sample is therefore 127 countries for wasting and 126 countries for stunting.

#### ***Individual staple model specification***

Six individual staple models were constructed — one per cereal or tuber — with a fixed set of care and disease covariates separately specified for stunting and wasting outcomes. This approach improves the events-per-variable ratio, avoids inter-staple collinearity, and isolates each staple's independent contribution to outcome variance. A combined model including all six staples simultaneously was not used for the primary analysis.

A note on the millet variable: although millet encompasses nutritionally distinct species — including pearl millet, finger millet, teff and other minor millets, which differ substantially in protein quality and geographic distribution — FAOSTAT food balance sheets report millet as a single undifferentiated aggregate (Item Code 2517, “Millet and products”; Item Code 79 in FAOSTAT definitions and standards, CPC 0118). The species composition of this aggregate is not documented in FAOSTAT definitions and standards: a search of the FAOSTAT definitions and standards tab for “millet” returns 22 items, none identifying constituent species, and “pearl millet” returns no results. The millet model was constructed using this undifferentiated aggregate; results should be interpreted in light of this limitation. Additionally, millet and sorghum are highly collinear in the dataset (Spearman r=+0·675) with near-identical Sahelian geographic distribution, meaning their individual model coefficients may partly reflect shared geographic and dietary patterning rather than staple-specific nutritional effects. Disaggregation by millet species and more granular subnational consumption data would be needed to resolve this.

#### ***Excluded variables and rationale***

The following variables were not included in the analytical models (detailed rationale in S Table 9):

• **Low birth weight, short stature and maternal BMI** — downstream of dietary factors; over-adjustment on the causal pathway.

• **Breastfeeding variables** — breast milk quality is affected by maternal diet and could mediate the relationship under study.^21^

• **Dietary diversity indices** — components already represented individually in the covariate set.

• **Tuberculosis** — collinearity with LRI prevalence (r=0·74); circularity concerns regarding GBD LRI estimates.

• **Nematodes** — low signal with either outcome; collinear with other disease burden covariates. See S Table 7.

• **HAQ index and WASH SEVs** — excluded on multicollinearity grounds: HAQ index correlates strongly with SDI (r=0·867) and animal protein supply (r=0·670); WASH SEVs (unsafe sanitation, unsafe water) correlate strongly with SDI (r=−0·839 and r=−0·831 respectively) and with each other (r=0·787) (see S Table 8). Inclusion alongside SDI and UHC would introduce severe multicollinearity given the limited EPV available (see S Table 7). WASH SEVs are additionally GBD-modelled exposure estimates derived using covariates overlapping with those in the present model, raising a secondary circularity concern distinct from the unmodelled status of the JME outcome variables.

• **Non-cereal plant protein** — negligible association with either outcome (Spearman r<0·10).

• **Malaria (wasting models only)** — weak association with wasting (r=0·21); geographic concentration in SSA creates collinearity. Retained in stunting models — see S Table 7.

• **SDI (wasting models only)** — relationship between SDI and wasting is non-monotonic at country level; higher-SDI South and Southeast Asian countries carry disproportionately high wasting burdens similar to Indian states with higher SDI with higher wasting and lower stunting in comparison to lower SDI states which have higher stunting and lower wasting^11^— see S Table 9 and S Section A.

### **S Section C. Data sources and availability**

This study uses publicly available data in accordance with GATHER (Guidelines for Accurate and Transparent Health Estimates Reporting). Data sources are detailed in S Table 2. WHO/JME prevalence thresholds and severity classifications for under-5 wasting and stunting used throughout this analysis are presented in S Table 3. The country list of 141 LMICs, extracted from the OECD Development Assistance Committee (DAC) operational list for eligibility in official development assistance (ODA), is provided in S Table 4. Missing data and excluded countries are detailed in S Table 5. Descriptive statistics for all analytical variables are presented in S Table 6 with status with respect to normality of data.

A STROBE checklist for cross-sectional studies is available in S Table 1.^22^

**Child undernutrition outcomes:** JME March 2025 (WHO, UNICEF, World Bank). Unmodelled survey estimates were used with reference population as the WHO Child Growth Standards 2006.

**Disease and care covariates:** These were extracted from GBD 2023, GHDx (measure_id=5, metric_id=2). ghdx.healthdata.org.

**Dietary covariates:** FAOSTAT food balance sheets 2019–2023 (five-year means). FAOSTAT data quality flags per Filipczuk T, FAO Rome, personal communication, 17 November 2025.

**Population weights (S5 only):** UN IGME 2023 under-5 population estimates. childmortality.org.

**LMIC classification:** OECD DAC list of ODA recipients 2022–2023 (141 countries; analytical sample 127 wasting, 126 stunting).

**S Table 1. STROBE checklist for cross-sectional studies**

| **Item** | **Recommendation** | **Where reported** |
| --- | --- | --- |
| **TITLE AND ABSTRACT** | | |
| **1** | Title and abstract: (a) Indicate study design in title/abstract; (b) Provide informative and balanced abstract summary | (a) "“Ecological cross-sectional analysis” in title and opening Methods sentence. Secondary cross-reference analyses used existing survey datasets (Grellety & Golden, n=47 surveys; Kerac et al, n=56 countries) linked to FAOSTAT staple protein supply by country. (b) Structured abstract ≤250 words with design, sample, methods, key findings and implications. |
| **INTRODUCTION** | | |
| **2** | Background/rationale: Explain scientific background and rationale | Introduction: protein quality heterogeneity across staples, DIAAS framework, unexplained wasting variance across LMICs. |
| **3** | Objectives: State specific objectives including pre-specified hypotheses | Introduction: To test two hypotheses: (1) That differences in dietary staple type — reflecting variation in protein quality (DIAAS), protein quantity and zinc bioavailability — explain substantial heterogeneity in child wasting and stunting prevalence across LMICs, after controlling for care and disease covariates (primary analysis; n=127 wasting, n=126 stunting). (2) That the staple-driven mechanism identified in the primary analysis is reflected in independent anthropometric datasets: specifically, that sorghum- and millet-dominant countries show systematic underestimation of true wasting prevalence by WHZ relative to MUAC, while maize- and cassava-dominant countries show the converse pattern (Grellety & Golden cross-reference, n=47 surveys), and that this same staple gradient is associated with the prevalence of simultaneous wasting-and-stunting in infants under 6 months (Kerac et al cross-reference, n=56 countries). |
| **METHODS** | | |
| **4** | Study design: Present key elements of study design early in the paper | Ecological cross-sectional analysis — stated in opening Methods sentence. |
| **5** | Setting: Describe setting, locations, and relevant dates | 127 LMICs (analytical sample after exclusions from 141 OECD DAC list countries). FAOSTAT 2019–2023 (dietary). JME March 2025 (outcomes, latest available unmodelled survey). |
| **6** | Participants: Give eligibility criteria and methods of selection | Countries: 131 LMICs with JME data; 127 wasting, 126 stunting after exclusions (see S Table 5). Eligibility: OECD DAC list + JME data available. |
| **7** | Variables: Define all outcomes, exposures, predictors, confounders | Methods and S Table 2: six staple protein supply variables (g/capita/d); fixed covariates per outcome — UHC, LRI, diarrhoea, HIV (wasting), malaria (stunting), SDI (stunting), animal protein, total kcal. |
| **8** | Data sources: Give sources and methods for each variable | S Table 2: sources, item codes, time periods, definitions for all 18 variables. |
| **9** | Bias: Describe efforts to address potential bias | Methods and Limitations: temporal mismatch addressed by FAOSTAT 2019–2023 five-year means; GBD covariate multicollinearity and circularity assessed formally (S Table 8); covariate selection via UNICEF framework excluding intermediates (BMI, short stature, low birthweight); systematic circularity and collinearity assessment (S Table 7); FAOSTAT data quality addressed by regional imputation and Somalia exclusion and sensitivity analysis excluding countries with imputed/old data(S2); ecological fallacy and WHZ measurement bias acknowledged in Limitations (S Section F); the bias introduced by JME's reliance on WHZ-based wasting prevalence is partially mitigated by the secondary cross-reference analyses, which draw on survey-level datasets containing concurrent MUAC, WHZ and oedema measurements (Grellety & Golden, Kerac et al), enabling assessment of the staple-stratified discordance between WHZ and MUAC-based wasting classifications. |
| **10** | Study size: Explain how study size was arrived at | All LMICs with available data: n=127 (wasting), n=126 (stunting). Determined by data availability, not power calculation. |
| **11** | Quantitative variables: Explain how they were handled | Protein supply continuous (g/capita/d). Outcomes continuous (%). All variables on natural scale. Robust MM-estimation (primary) handles non-normality and influential observations; quantile regression (secondary) examines consistency of associations across the outcome distribution (Q25, Q50, Q75). |
| **12** | Statistical methods: Describe all methods | (a) Robust MM-estimation (robustbase, KS2014); one staple per model — EPV constraints (n=127–128, 6–7 fixed covariates, EPV=18–21); (b) Quantile regression Q0·25/0·50/0·75; (c) S Table 5 missing data; (d) N/A ecological; (e) Five sensitivity analyses S1–S5. |
| **RESULTS** | | |
| **13** | Participants: Report numbers at each stage | Primary analysis: 139 OECD DAC LMICs → 132 with JME survey data → 128 after exclusions (Somalia: FAOSTAT absent, JME 2009; Bulgaria: high income; Palestinian Territories: no staple data; DPRK: no JME data) → 127 wasting / 126 stunting (Russia: wasting only). See S Table 5. Secondary cross-reference: Grellety & Golden: 46 DHS/SMART surveys, n=47 observations after FAOSTAT matching. Kerac et al: 56 DHS/MICS datasets, n=56 countries after FAOSTAT matching. |
| **14** | Descriptive data: Give characteristics and missing data counts | S Table 6: descriptive statistics for all variables. S Table 5: missing data by variable. |
| **15** | Outcome data: Report summary measures | Wasting range 0·6–22·7%; stunting range 1·2–52·8% across included countries (JME March 2025). |
| **16** | Main results: Give adjusted estimates and precision | All results from robust MM-estimation with full covariate adjustment. β, SE, p-values in main Tables 2–3 and S Tables 9–13. |
| **17** | Other analyses: Report subgroup, sensitivity analyses | Sensitivity analyses S1–S5 (S Table 13, Panels A–E). Quantile regression (S Tables 10–11). Grellety-Golden cross-reference (S Table 21). |
| **DISCUSSION** | | |
| **18** | Key results: Summarise with reference to objectives | Discussion paragraph 1 summarises primary findings with reference to DIAAS profiles and zinc mechanisms. |
| **19** | Limitations: Discuss limitations and potential bias | Limitations section: ecological fallacy, FAOSTAT representativeness, DIAAS data gaps, WHZ measurement, temporal heterogeneity. |
| **20** | Interpretation: Give cautious overall interpretation | Discussion and conclusion interpret findings as Findings are interpreted as hypothesis-generating, consistent with the nutritional profiles of the six staples summarised in S Table 24: sorghum (lowest DIAAS, highest phytate:zinc ratio) shows the strongest and most consistent positive association with wasting, while maize and cassava (high phytate:zinc, near-absent protein respectively) show negative or null wasting associations alongside positive stunting trends — a pattern explained by stunting-mediated WHZ attenuation rather than true nutritional protection. Convergent evidence from: (1) the Grellety & Golden cross-references (WHZ-predominant wasting in sorghum/millet-dominant countries; MUAC-predominant wasting in maize/cassava-dominant countries, consistent with the stunting-mediated height-denominator effect on WHZ); and (2) the Kerac et al cross-reference (sorghum and millet supply associated with higher simultaneous wasting-and-stunting in infants under 6 months) — together strengthen biological plausibility without establishing causality. Ecological design precludes individual-level inference. Findings should be interpreted in the context of dietary transitions, food systems, and the multi-factorial determinants of child undernutrition. |
| **21** | Generalisability: Discuss external validity | Limitations: ecological associations cannot be extrapolated to individuals; individual-level corroboration precluded by DHS/MICS survey design. |
| **OTHER INFORMATION** | | |
| **22** | Funding: Give source of funding and role of funders | No funding received. Authors declare no conflicts of interest. |

*STROBE: STrengthening the Reporting of OBservational studies in Epidemiology. Adapted for ecological cross-sectional studies.*

**S Table 2. Data sources**

| **Variable** | **Source** | **Definition** | **Time period** | **Access / notes** |
| --- | --- | --- | --- | --- |
| **Stunting & wasting (outcomes)** | JME March 2025 — WHO, UNICEF, World Bank | Under-5 prevalence, both sexes, 0–59 months. Latest unmodelled survey estimate. | 2000–2024 (by country) | *who.int/data/gho/data/themes/topics/joint-child-malnutrition-estimates-unicef-who-wb* |
| **LRI, diarrhoea, HIV, malaria, TB, nematodes** | GBD 2023 GHDx measure_id=5, metric_id=2 | Under-5 prevalence, both sexes. Used: LRI, diarrhoea, HIV (wasting), malaria (stunting). Excluded: TB (collinear with LRI, r=0·74), nematodes (see S Table 7). | 2023 | *ghdx.healthdata.org/gbd-results/* |
| **SDI** | GBD 2023 GHDx (covariate_id=881) | Socio-demographic Index — composite of income, education, fertility. | 2023 | *ghdx.healthdata.org* |
| **UHC index** | GBD 2023 GHDx (covariate_id=1097) | UHC effective coverage index (0–1 scale). | 2023 | *ghdx.healthdata.org* |
| **Staple protein supply** | FAOSTAT food balance sheets | Protein supply (g/capita/d): wheat (2511), rice (2807), maize (2514), millet (2517), sorghum (2518), cassava (2532). | 2019–2023 (5-year mean) | *fao.org/faostat/en/#data/FBS* |
| **Animal protein** | FAOSTAT food balance sheets | Sum from 14 animal food categories (bovine, mutton, pig, poultry, offal, eggs, milk, freshwater/demersal/pelagic/marine fish, crustaceans, molluscs, other). | 2019–2023 (5-year mean) | *fao.org/faostat* |
| **Total kcal/d** | FAOSTAT food balance sheets | Total dietary energy availability per capita per day (Grand Total, item code S2901). | 2019–2023 (5-year mean) | *fao.org/faostat* |
| **Population weights (S5 only)** | UN IGME 2023 | Under-5 population (thousands), both sexes, 2020 estimate. | 2020 | *childmortality.org* |
| **LMIC classification** | OECD DAC list of ODA recipients | Low-, lower-middle- and upper-middle-income countries. | 2022–2023 | *oecd.org/dac* |

**S Table 3. WHO/JME prevalence thresholds and severity classifications for under-5 wasting and stunting**

| **Severity label** | **Stunting (%)** | **Wasting (%)** |
| --- | --- | --- |
| Very low | <2·5 | <2·5 |
| Low | 2·5–<10 | 2·5–<5 |
| Medium | 10–<20 | 5–<10 |
| High | 20–<30 | 10–<15 |
| Very high | ≥30 | ≥15 |

*Source: WHO/UNICEF/World Bank Joint Malnutrition Estimates (JME) 2025. Wasting thresholds are set at lower prevalence levels than stunting, reflecting higher mortality risk at lower population prevalence. These classifications are used throughout this analysis to characterise country-level burden.*

**S Table 4. Country list — all 141 LMICs (OECD DAC list 2022–2023)**

| **A–C** | **D–L** | **M–R** | **S–Z** |
| --- | --- | --- | --- |
| Afghanistan | Egypt | Malawi | São Tomé and Príncipe |
| Albania | El Salvador | Malaysia | Senegal |
| Algeria | Equatorial Guinea | Maldives | Serbia |
| Angola | Eritrea | Mali | Sierra Leone |
| Argentina | Eswatini | Marshall Islands | Solomon Islands |
| Armenia | Ethiopia | Mauritania | *Somalia†* |
| Azerbaijan | Fiji | Mauritius | South Africa |
| Bangladesh | Gabon | Mexico | South Sudan |
| Belarus | Gambia | *Micronesia (Fed. States)‡* | Sri Lanka |
| Belize | Georgia | Moldova | Sudan |
| Benin | Ghana | Mongolia | Suriname |
| Bhutan | *Grenada‡* | Montenegro | Syrian Arab Republic |
| Bolivia | Guatemala | *Montserrat‡* | Tajikistan |
| Bosnia and Herzegovina | Guinea | Morocco | Tanzania |
| Botswana | Guinea-Bissau | Mozambique | Thailand |
| Brazil | Guyana | Myanmar | Timor-Leste |
| Burkina Faso | Haiti | Namibia | Togo |
| Burundi | Honduras | Nepal | *Tokelau‡* |
| Cabo Verde | India | Nicaragua | Tonga |
| Cambodia | Indonesia | Niger | Tunisia |
| Cameroon | Iran | Nigeria | Türkiye |
| Central African Republic | Iraq | *Niue‡* | Turkmenistan |
| Chad | Jamaica | North Macedonia | *Tuvalu‡* |
| China | Jordan | Pakistan | Uganda |
| Colombia | Kazakhstan | Panama | Ukraine |
| Comoros | Kenya | Papua New Guinea | Uzbekistan |
| Congo | Kiribati | Paraguay | Vanuatu |
| Côte d'Ivoire | Kosovo | Peru | Venezuela |
| Cuba | Kyrgyzstan | Philippines | Viet Nam |
| *Dem. People's Rep. Korea†* | Lao PDR | Rwanda | *Wallis and Futuna‡* |
| Democratic Republic of the Congo | Lebanon | *Saint Helena‡* | *West Bank and Gaza Strip‡* |
| Djibouti | Lesotho | *Saint Kitts and Nevis‡* | Yemen |
| *Dominica‡* | Liberia | Saint Lucia | Zambia |
| Dominican Republic | Libya | *Saint Vincent and Grenadines‡* | Zimbabwe |
| Ecuador | Madagascar | Samoa |  |

*† Excluded from all analyses: Somalia — FAOSTAT food balance sheet data unavailable 2019–2023 (Filipczuk T, FAO Rome, personal communication, 17 November 2025) and JME wasting survey year 2009 (15-year temporal mismatch). Democratic People's Republic of Korea (DPRK) — dietary data entirely absent from FAOSTAT; only LMIC regional comparator in East Asia is China, making regional mean imputation invalid.*

*‡ Excluded from all analyses — missing JME stunting and wasting data: Dominica, Grenada, Micronesia (Fed. States), Montserrat, Niue, Saint Helena, Saint Kitts and Nevis, Saint Vincent and the Grenadines, Tokelau, Tuvalu, Wallis and Futuna, West Bank and Gaza Strip.*

*Mauritius and Cabo Verde: JME stunting data available but wasting data absent — excluded from wasting analyses only.*

*Analytical sample: n=127 countries (wasting), n=126 countries (stunting). Total OECD DAC list: 141 countries.*

**S Table 5. Missing data and excluded countries**

| **Issue / missing data** | **Affected countries and exclusion decision** | **Method** | **Rationale** | **Impact on analytical sample** |
| --- | --- | --- | --- | --- |
| **JME data absent (stunting and wasting)** | Missing: Grenada, Micronesia (Fed. States), Saint Vincent and the Grenadines, Dominica, Montserrat, Niue, Saint Helena, Saint Kitts and Nevis, Tokelau, Tuvalu, Wallis and Futuna, West Bank and Gaza Strip → Excluded from all analyses | Delete | JME unmodelled survey-based wasting/stunting estimates unavailable for these countries. GBD modelled estimates exist for some but were not used — unmodelled JME estimates were preferred to avoid circularity with GBD-derived covariates (see Dependent variables, Methods). | *Excluded from all analyses (n not included in 132)* |
| **JME stunting data absent** | Missing: Russian Federation → Excluded from stunting analyses only | Delete | No JME stunting estimate available. Wasting data available and used. | *Excluded from stunting analyses only* |
| **JME wasting data absent** | Missing: Mauritius, Cabo Verde → Excluded from wasting analyses only | Delete | Wide seasonal fluctuation in wasting makes modelling unreliable; JME does not produce wasting estimates for these countries. | *Excluded from wasting analyses only* |
| **FAOSTAT food balance sheet data absent or masked** | Somalia: FAOSTAT FBS 2019–2023 unavailable and masked for data quality reasons (Filipczuk T, FAO Rome, personal communication, 17 November 2025); additionally JME wasting survey year 2009 — 15-year temporal mismatch, oldest survey in dataset → Excluded from all analyses Democratic People's Republic of Korea (DPRK): dietary data entirely absent from FAOSTAT; only LMIC regional comparator in East Asia is China — regional mean imputation not valid → Excluded from all analyses Eritrea, Equatorial Guinea: FAOSTAT values never compiled for these countries (Filipczuk T, 17 November 2025) — values imputed as regional mean (Eastern SSA for Eritrea; Central SSA for Equatorial Guinea) → Retained in primary analysis; excluded in S2 | Somalia, DPRK: Delete Eritrea, Eq. Guinea: Regional mean imputation | Data quality insufficient for inclusion. Somalia and DPRK excluded on data availability grounds. Eritrea and Equatorial Guinea imputed where possible; excluded in S2 sensitivity analysis testing robustness to FAOSTAT data quality. | *Somalia, DPRK: excluded all analyses Eritrea, Eq. Guinea: retained primary; excluded S2* |
| **Malaria prevalence absent (GBD 2023)** | Missing values: Eswatini, Armenia, Ukraine, Uzbekistan, Tuvalu → Retained with value set to zero | Set to zero | All five countries are at or near malaria elimination. Setting to zero is epidemiologically consistent with their malaria status (WHO World Malaria Report 2022). | *Retained in all analyses with zero values* |
| **S2 sensitivity analysis exclusions (FAOSTAT data quality)** | Masked/unavailable 2019–2023: Benin, Burundi, Central African Republic, Chad, Mali, South Sudan, Sudan, Togo Never compiled: Eritrea, Equatorial Guinea Total: 11 countries excluded in S2 (including Somalia, already excluded from primary) | Excluded in S2 only | S2 tests robustness of primary findings to exclusion of all countries with FAOSTAT data quality concerns. Primary analysis retains Eritrea and Equatorial Guinea with imputed values. | *n=118 (wasting), n=117 (stunting) in S2* |
| **FAOSTAT dietary data never compiled — imputed** | Eritrea, Equatorial Guinea | Impute from unweighted mean of GBD super-region neighbours | FAOSTAT food balance sheet data have never been compiled for these countries (Filipczuk T, FAO Rome, personal communication, 17 November 2025). Values imputed from mean of neighbouring countries in same GBD super-region. | *Retained in primary analysis with imputed values; excluded in S2 sensitivity analysis* |

**S Table 6. Descriptive statistics — all analytical variables (n=127 wasting, n=126 stunting)**

| **Variable** | **n** | **Mean** | **SD** | **Median** | **IQR** | **Min** | **Max** | **Skewness** | **SW p** | **Normality** |
| --- | --- | --- | --- | --- | --- | --- | --- | --- | --- | --- |
| Wasting prevalence (%) | 127 | 5·378 | 4·075 | 4·150 | 2·400–7·200 | 0·600 | 22·700 | 0·90 | <0·001 | Reject |
| Stunting prevalence (%) | 126 | 20·359 | 12·686 | 17·600 | 8·950–28·900 | 1·200 | 52·800 | 0·65 | <0·001 | Reject |
| UHC index | 127 | 52·786 | 10·956 | 53·767 | 46·381–58·194 | 23·610 | 81·219 | −0·27 | 0·410 | Accept |
| SDI | 127 | 0·580 | 0·138 | 0·607 | 0·482–0·690 | 0·203 | 0·823 | −0·59 | 0·001 | Reject |
| LRI prevalence | 127 | 0·157 | 0·065 | 0·159 | 0·105–0·195 | 0·038 | 0·368 | −0·11 | 0·002 | Reject |
| Diarrhoea prevalence | 127 | 1·077 | 0·631 | 0·924 | 0·621–1·407 | 0·119 | 3·857 | 0·73 | <0·001 | Reject |
| HIV prevalence | 127 | 0·098 | 0·308 | 0·011 | 0·002–0·089 | 0·000 | 3·247 | 0·85 | <0·001 | Reject |
| Malaria prevalence | 127 | 5·116 | 9·202 | 0·170 | 0·000–4·460 | 0·000 | 36·772 | 1·61 | <0·001 | Reject |
| Animal protein (g/d) | 127 | 32·635 | 18·835 | 29·260 | 17·246–45·275 | 3·100 | 91·466 | 0·54 | <0·001 | Reject |
| Total energy (kcal/d) | 127 | 2769·4 | 420·1 | 2749·2 | 2517·9–3077·4 | 1860·7 | 3796·4 | 0·14 | 0·705 | Accept |
| Wheat protein (g/d) | 127 | 14·610 | 11·891 | 10·856 | 5·340–22·291 | 0·620 | 48·916 | 0·95 | <0·001 | Reject |
| Rice protein (g/d) | 127 | 7·602 | 7·857 | 5·220 | 1·715–10·630 | 0·326 | 32·980 | 0·91 | <0·001 | Reject |
| Maize protein (g/d) | 127 | 4·572 | 5·296 | 2·374 | 0·525–7·103 | 0·004 | 24·594 | 1·25 | <0·001 | Reject |
| Millet protein (g/d) | 127 | 0·611 | 2·217 | 0·000 | 0·000–0·198 | 0·000 | 18·288 | 0·83 | <0·001 | Reject |
| Sorghum protein (g/d) | 127 | 1·001 | 2·639 | 0·000 | 0·000–0·497 | 0·000 | 14·160 | 1·14 | <0·001 | Reject |
| Cassava protein (g/d) | 127 | 0·832 | 1·592 | 0·080 | 0·000–0·939 | 0·000 | 10·724 | 1·42 | <0·001 | Reject |

*SW = Shapiro-Wilk normality test. Skewness = Pearson's coefficient. Green shading = normality accepted (p>0·05). n=127 for wasting prevalence; n=126 for stunting prevalence; n=127 for all other variables. LRI, diarrhoea, HIV, malaria expressed as proportions (GBD 2023 GHDx output). Somalia excluded from analytical sample.*

**S Figure 1. Covariate selection process — five-step framework**

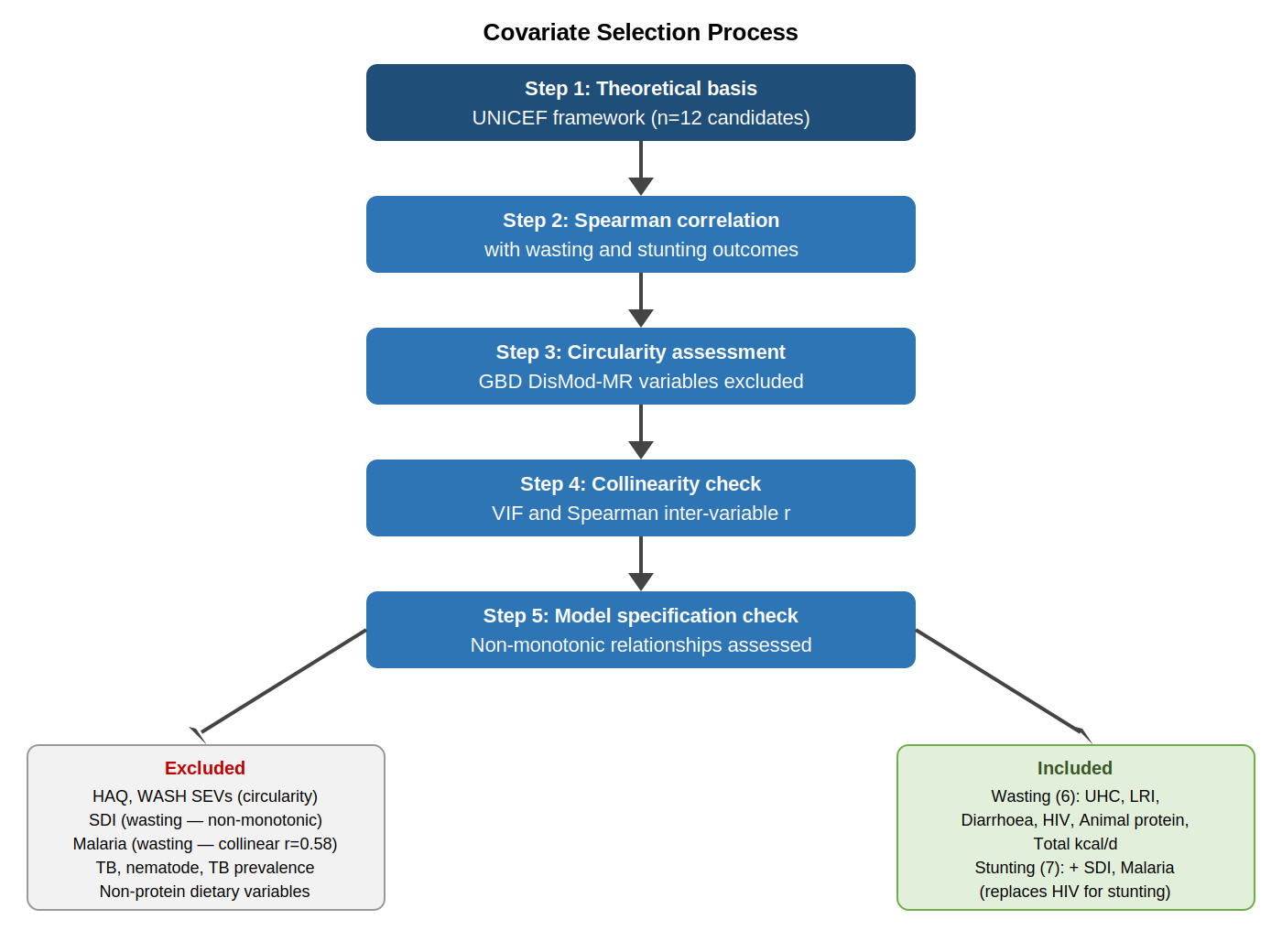

Covariate selection followed a five-step process: (1) theoretical basis from the UNICEF conceptual framework; (2) Spearman correlation with outcomes; (3) circularity assessment against GBD 2023 modelling frameworks (Shackelford et al., 2025); (4) collinearity check (VIF and inter-variable Spearman r); (5) model specification check for non-monotonic relationships. Separate covariate sets were specified for wasting (6 covariates: UHC, LRI, diarrhoea, HIV, animal protein, total kcal) and stunting (7 covariates: SDI, UHC, LRI, diarrhoea, malaria, animal protein, total kcal). Full inclusion/exclusion rationale in S Table 8.

### S Section D. Statistical Methods and Results

**S Table 7. Covariate and exposure variable assessment — Panels A–C**

**Panel A. Circularity assessment — study variables in relation to GBD 2023 modelling frameworks**

*Circularity arises when a variable used as a covariate in our regression was itself used as an input to the GBD modelling that generated our predictor variables (disease prevalence estimates). The JME survey-based wasting and stunting outcomes break the direct circularity loop at the outcome level. Risk levels were classified by the study authors: None = no pathway identified; Low = distal shared determinants only; Low–Moderate = indirect pathway via shared upstream inputs (e.g. HAQ or SDI as covariate in DisMod-MR model); Moderate = indirect pathway via shared upstream inputs or nutritional ecology; High = direct input to GBD cause model. Information on GBD modelling covariates was extracted from GBD 2023 Appendix 1 (Shackelford et al., 2025); covariate tables for LRI (Table 6), diarrhoea (Table 3), TB (Table 2), nematodes (pp 179, 1209), HIV (pp 664–676), malaria (pp 855–857).*

| **Variable** | **GBD modelling method** | **Covariates used in GBD model (GBD 2023 Appendix 1)** | **Circularity pathway with JME wasting/stunting outcome** | **Risk** | **Decision** |
| --- | --- | --- | --- | --- | --- |
| **Disease burden covariates** | | | | | |
| **LRI prevalence** | DisMod-MR 2.1 | SDI (country-level, prevalence); HAQ index (country-level, excess mortality). Table 6, GBD 2023 Appendix 1 | JME outcome is survey-based (not GBD-modelled). SDI and HAQ are distal predictors — no direct child anthropometry input to LRI DisMod-MR model. Indirect pathway only via shared distal determinants. However, HAQ index — rated High circularity and excluded from our models — is a direct covariate in the LRI DisMod-MR model, creating an indirect circularity pathway via shared upstream inputs. | **Low–Moderate** | INCLUDED (both models) |
| **Diarrhoea prevalence** | DisMod-MR 2.1 | Rotavirus vaccine coverage (lagged 5-year); SDI; HAQ index; sex. Table 3, GBD 2023 Appendix 1 | JME outcome is survey-based. No child anthropometry input to diarrhoea DisMod-MR model. However, HAQ index is a direct covariate in the diarrhoea DisMod-MR model alongside SDI and rotavirus vaccine coverage, creating an indirect pathway via shared upstream inputs. | **Low–Moderate** | INCLUDED (both models) |
| **HIV prevalence** | EPP-ASM/Spectrum compartmental model | Serosurveillance data from ANC clinics; HIV progression rates; ART coverage; demographic inputs. No child anthropometry. GBD 2023 Appendix 1 pp 664–676 | JME outcome is survey-based. No child anthropometry input to EPP-ASM or Spectrum. No circularity pathway identified. | **None** | INCLUDED wasting; EXCLUDED stunting (non-monotonic; r=+0·510 with SDI — see S Table 8) |
| **Malaria prevalence** | Bayesian spatiotemporal geostatistical model (MAP/Pf PR); DisMod-MR for sequelae | Environmental covariates (ITNs, IRS, precipitation, vegetation, elevation); intervention coverage. No child anthropometry as model input. GBD 2023 Appendix 1 pp 855–857 | JME outcome is survey-based. No child anthropometry input to malaria prevalence model. No circularity pathway identified. | **None** | INCLUDED stunting; EXCLUDED wasting (r=+0·21, collinear with SSA staple geography — see S Table 8) |
| **TB prevalence** | DisMod-MR 2.1 + MR-BRT | HAQ index; age-standardised adult underweight prevalence; TB SEV scalar. Table 2, GBD 2023 Appendix 1 p 1225 | Adult underweight prevalence is a direct model input to TB prevalence DisMod-MR. Adult underweight correlates with child wasting/stunting at country level. Indirect circularity pathway via shared nutritional ecology. | **Moderate** | EXCLUDED (both models). Also collinear with LRI (r=+0·37) — see S Table 8 |
| **Nematode (STH) prevalence** | ST-GPR global prevalence model + DisMod-MR for sequelae | ST-GPR covariates: SDI, sanitation, safe water. Sequela model uses wasting envelope to derive ‘severe wasting due to ascariasis’ via: Prev_wasting_ascariasis = wasting − Φ(Φ⁻¹(wasting) − z-score × heavy infestation). GBD 2023 Appendix 1 pp 179, 1209 | Nematode prevalence estimated from GAHI survey data via ST-GPR — wasting is not a direct model input. Downstream sequela sub-model uses wasting envelope to compute severe wasting due to ascariasis (GBD 2023 Appendix 1 p 179), but this does not affect the primary prevalence estimate. ST-GPR upstream covariates (WASH SEVs, SDI) shared with excluded variables. | **Moderate** | EXCLUDED (both models). Also low signal: r=+0·00 wasting, r=+0·27 stunting — see S Table 8 |
| **Care and structural covariates** | | | | | |
| **HAQ index (Healthcare Access and Quality Index)** | GBD composite index incorporating cause-specific mortality rates | Constructed from 32 cause-specific mortality rates, several of which use child health outcomes. Directly incorporates mortality from conditions associated with child undernutrition. GBD 2023 Appendix 1 | HAQ construction incorporates mortality patterns strongly correlated with child wasting and stunting. Direct circularity pathway. | **High** | EXCLUDED (both models). Also collinear with SDI (r=+0·87) — see S Table 8 |
| **WASH SEVs (unsafe sanitation; unsafe water)** | ST-GPR modelled exposure estimates | ST-GPR covariates: SDI, proportion using improved sanitation/water, urbanisation. GBD 2023 risk factor study | JME outcome is survey-based. No direct child anthropometry input. However WASH SEVs are GBD-modelled using many of the same country-level covariates as disease burden variables, creating shared upstream structure. | **Moderate** | EXCLUDED (both models). Collinear with SDI (sanitation r=−0·84; water r=−0·83) and HAQ (r=−0·83) — see S Table 8 |
| **UHC index (Universal Health Coverage index)** | WHO/GBD composite index — independent tracer indicator coverage | Constructed from coverage of essential health services. Absent from all GBD 2023 DisMod-MR and CODEm cause-specific models for LRI, diarrhoea, malaria, TB, HIV. Confirmed by systematic search of GBD 2023 Appendix 1 | No circularity pathway identified. Not derived from any GBD cause-specific model used in present analysis. | **None** | INCLUDED (both models) |
| **SDI (Socio-demographic Index)** | GBD composite index (income per capita, educational attainment, total fertility rate) | Level 3 covariate in LRI, diarrhoea, and TB DisMod-MR/CODEm models. Also country-level covariate in LRI DisMod-MR prevalence model (Table 6, GBD 2023 Appendix 1). Not child anthropometry. | SDI is a shared upstream input to both LRI DisMod-MR and our stunting model. Indirect circularity only. JME survey-based outcome breaks the direct loop. SDI and LRI retain distinct coefficients in stunting models. | **Low–moderate** | INCLUDED stunting only. EXCLUDED wasting — non-monotonic relationship (see S Table 8) |
| **Dietary covariates** | | | | | |
| **Animal protein supply (g/capita/day)** | FAOSTAT Food Balance Sheets — supply-side national estimate | Not a GBD modelled variable. Direct FAOSTAT data. | No circularity pathway. Entirely absent from all GBD DisMod-MR and CODEm modelling frameworks. | **None** | INCLUDED (both models) |
| **Total energy supply (kcal/capita/day)** | FAOSTAT Food Balance Sheets | Not a GBD modelled variable. | No circularity pathway. | **None** | INCLUDED (both models) as fixed covariate to control for energy availability. Moderate collinearity with animal protein (r=+0·70) and SDI (r=+0·70) acknowledged; all three retain distinct coefficients capturing separable dimensions — see S Table 8 |
| **Non-cereal plant protein (g/capita/day)** | FAOSTAT Food Balance Sheets | Not a GBD modelled variable. | No circularity pathway. | **None** | EXCLUDED (both models). Negligible signal: r=+0·11 wasting (p=0·21), r=+0·19 stunting (p=0·03) — see S Table 8 |
| **Staple protein supply — wheat, rice, maize, millet, sorghum, cassava (g/capita/day)** | FAOSTAT Food Balance Sheets | Not GBD modelled variables. | No circularity pathway. Primary exposure variables. | **None** | INCLUDED — primary exposures (each modelled separately) |

**Panel B. Inter-variable Spearman correlation matrix — all candidate covariates (n=127 LMICs)**

*Spearman rank correlations among all 14 candidate predictor variables. Red = strong positive (|r| ≥0·70); blue = strong negative. *** p<0·001, ** p<0·01, * p<0·05. Diagonal = 1·000 (shaded). n=127 countries (corrected dataset, NA→0 applied for millet/sorghum/cassava). Computed from validation_dataset_final_127LMIC_corrected.csv.*

| **Variable** | **SDI** | **UHC** | **LRI** | **Diarrhoea** | **HIV** | **Malaria** | **TB** | **Nematodes** | **Animal protein** | **Total kcal** | **HAQ** | **WASH sanitation** | **WASH water** | **Non-cereal protein** |
| --- | --- | --- | --- | --- | --- | --- | --- | --- | --- | --- | --- | --- | --- | --- |
| **SDI** | 1.000 | +0.533*** | -0.631*** | -0.296*** | -0.585*** | -0.765*** | -0.460*** | -0.231** | +0.777*** | +0.692*** | +0.865*** | -0.843*** | -0.828*** | -0.244** |
| **UHC** | +0.533*** | 1.000 | -0.482*** | -0.167 | -0.297*** | -0.312*** | -0.429*** | -0.087 | +0.283** | +0.267** | +0.622*** | -0.454*** | -0.467*** | +0.043 |
| **LRI** | -0.631*** | -0.482*** | 1.000 | +0.503*** | +0.388*** | +0.425*** | +0.364*** | +0.096 | -0.564*** | -0.543*** | -0.646*** | +0.515*** | +0.618*** | +0.066 |
| **Diarrhoea** | -0.296*** | -0.167 | +0.503*** | 1.000 | +0.202* | +0.358*** | +0.032 | -0.041 | -0.373*** | -0.259** | -0.285** | +0.258** | +0.376*** | +0.185* |
| **HIV** | -0.585*** | -0.297*** | +0.388*** | +0.202* | 1.000 | +0.651*** | +0.538*** | +0.262** | -0.462*** | -0.565*** | -0.730*** | +0.638*** | +0.672*** | +0.144 |
| **Malaria** | -0.765*** | -0.312*** | +0.425*** | +0.358*** | +0.651*** | 1.000 | +0.356*** | +0.216* | -0.695*** | -0.634*** | -0.734*** | +0.731*** | +0.731*** | +0.337*** |
| **TB** | -0.460*** | -0.429*** | +0.364*** | +0.032 | +0.538*** | +0.356*** | 1.000 | +0.384*** | -0.206* | -0.329*** | -0.619*** | +0.465*** | +0.455*** | +0.101 |
| **Nematodes** | -0.231** | -0.087 | +0.096 | -0.041 | +0.262** | +0.216* | +0.384*** | 1.000 | -0.063 | -0.269** | -0.282** | +0.194* | +0.195* | -0.002 |
| **Animal protein** | +0.777*** | +0.283** | -0.564*** | -0.373*** | -0.462*** | -0.695*** | -0.206* | -0.063 | 1.000 | +0.695*** | +0.665*** | -0.721*** | -0.685*** | -0.283** |
| **Total kcal** | +0.692*** | +0.267** | -0.543*** | -0.259** | -0.565*** | -0.634*** | -0.329*** | -0.269** | +0.695*** | 1.000 | +0.658*** | -0.676*** | -0.646*** | -0.093 |
| **HAQ** | +0.865*** | +0.622*** | -0.646*** | -0.285** | -0.730*** | -0.734*** | -0.619*** | -0.282** | +0.665*** | +0.658*** | 1.000 | -0.828*** | -0.807*** | -0.135 |
| **WASH sanitation** | -0.843*** | -0.454*** | +0.515*** | +0.258** | +0.638*** | +0.731*** | +0.465*** | +0.194* | -0.721*** | -0.676*** | -0.828*** | 1.000 | +0.789*** | +0.267** |
| **WASH water** | -0.828*** | -0.467*** | +0.618*** | +0.376*** | +0.672*** | +0.731*** | +0.455*** | +0.195* | -0.685*** | -0.646*** | -0.807*** | +0.789*** | 1.000 | +0.245** |
| **Non-cereal protein** | -0.244** | +0.043 | +0.066 | +0.185* | +0.144 | +0.337*** | +0.101 | -0.002 | -0.283** | -0.093 | -0.135 | +0.267** | +0.245** | 1.000 |

Because each staple’s protein supply was entered in a separate model (one staple per model, per S Table 10a), pairwise correlation between staples does not inflate standard errors within any single model in the way conventional multicollinearity would. However, where two staples are strongly correlated across countries, the coefficient on one may partly capture the true association of the other, since each staple-specific model omits the other five staples as covariates. We therefore computed pairwise Spearman and Pearson correlations between all six staple protein supply variables across the analytical sample (n=130 countries with complete staple data) to identify pairs at risk of this omitted-variable bias.

Sorghum and millet protein supply were the most strongly correlated pair of all 15 staple combinations (Spearman r=0·675, Pearson r=0·705, both p<0·0001), sharing approximately 50% of their variance (Pearson r²=0·497) — consistent with the geographic co-consumption of these two staples across the Sahelian belt (S Tables 15–16). Wheat and cassava showed the strongest rank-order association in the opposite direction (Spearman r=−0·750, Pearson r=−0·435), reflecting the largely mutually exclusive geographic distribution of wheat-dominant and cassava-dominant agricultural systems. All other staple pairs showed weaker associations (|Pearson r|≤0·425; S Table 7, Panel C).

The sorghum-millet collinearity (r=+0·675) is the most plausible explanation for the comparatively unstable, sign-reversing millet-stunting association across sensitivity specifications (S Table 12, Panel B: negative in Primary, S1 and S2; positive in S3, S4 and S5, significant only in S5), compounded by the heterogeneity within FAOSTAT’s undifferentiated millet variable (S Table 19). By contrast, sorghum’s wasting association remains significant across all six specifications including S3, making it less plausible that sorghum’s primary wasting finding is itself a collinearity artefact.

**Panel C. Inter-staple collinearity matrix**

| **Staple pair** | **n** | **Spearman r** | **Spearman p** | **Pearson r** | **Pearson p** |
| --- | --- | --- | --- | --- | --- |
| **Millet–Sorghum** | 130 | **+0·675** | <0·0001 | **+0·705** | <0·0001 |
| **Wheat–Cassava** | 130 | **−0·750** | <0·0001 | **−0·435** | <0·0001 |
| Wheat–Rice | 130 | −0·418 | <0·0001 | −0·425 | <0·0001 |
| Wheat–Maize | 130 | −0·350 | <0·0001 | −0·343 | <0·0001 |
| Wheat–Sorghum | 130 | −0·429 | <0·0001 | −0·240 | 0·0059 |
| Wheat–Millet | 130 | −0·368 | <0·0001 | −0·204 | 0·0197 |
| Rice–Cassava | 130 | +0·397 | <0·0001 | +0·046 | 0·606 |
| Rice–Maize | 130 | −0·172 | 0·050 | −0·254 | 0·0036 |
| Maize–Sorghum | 130 | +0·429 | <0·0001 | +0·118 | 0·180 |
| Maize–Cassava | 130 | +0·202 | 0·0214 | +0·093 | 0·292 |
| Millet–Cassava | 130 | +0·243 | 0·0054 | −0·051 | 0·564 |
| Sorghum–Cassava | 130 | +0·253 | 0·0037 | +0·019 | 0·019 |
| Rice–Millet | 130 | +0·028 | 0·749 | −0·000 | 0·0021 |
| Rice–Sorghum | 130 | −0·007 | 0·933 | −0·124 | 0·160 |

*Correlations computed on staple protein supply (g/capita/day, FAOSTAT 2019–2023 five-year means) across the n=130 countries in our analytical sample with complete data for all six staples (a subset of the full 127/126-country wasting/stunting samples, which permit missingness in individual staples). Pairs sorted by descending absolute Pearson r. Bold indicates the two strongest pairs (|Pearson r|>0·40 and |Spearman r|>0·65).*

**S Table 8. Covariate inclusion/exclusion — detailed rationale**

*One row per candidate variable. r wasting / r stunting = Spearman rank correlation with JME wasting/stunting prevalence (n=127/126). VIF = variance inflation factor computed from OLS auxiliary regression including all 14 candidate covariates simultaneously (n=127). Threshold: VIF ≥5 indicates unacceptable multicollinearity (⚠). Circ. = circularity risk per S Table 7, Panel A. Decision column reflects final choice for primary models; see sensitivity analyses S1–S5 (S Table 13). *** p<0·001, ** p<0·01, * p<0·05, † p<0·1, ns = not significant. GBD modelling covariate information from GBD 2023 Appendix 1 (Shackelford et al., 2025); circularity risk classification by study authors (see S Table 7, Panel A).*

| **Variable** | **r wasting (p)** | **r stunting (p)** | **VIF** | **Circ. risk** | **Decision** | **Rationale** |
| --- | --- | --- | --- | --- | --- | --- |
| **LRI** | +0.528 (***) | +0.677 (***) | 2.74 | Low | **INCLUDED both models** | Strongest bivariate predictor in both models (r=+0·53 wasting, r=+0·68 stunting, both p<0·001). DisMod-MR covariates are SDI and HAQ only — no child anthropometry input. JME survey-based outcome breaks direct circularity loop. Coefficient highly robust across all five sensitivity analyses (wasting β range +63·4 to +67·1; stunting β range +30·2 to +77·7, all p<0·001). VIF 2·74 well within tolerance. |
| **Diarrhoea** | +0.350 (***) | +0.442 (***) | 1.61 | Low | **INCLUDED both models** | Significant positive association with both outcomes (r=+0·35 wasting p<0·001; r=+0·44 stunting p<0·001). DisMod-MR covariates: rotavirus vaccine coverage, SDI, HAQ — no child anthropometry. JME outcome breaks direct loop. Diarrhoea and LRI retain independent variance (inter-variable r=+0·51, VIF 1·61 acceptable). UNICEF conceptual framework theoretical basis. Retained across all specifications. |
| **HIV** | +0.263 (**) | +0.510 (***) | 1.39 | None | **INCLUDED wasting EXCLUDED stunting** | EPP-ASM/Spectrum model uses no child anthropometry. No circularity. INCLUDED in wasting models: Spearman r=+0·26 (p=0·003) — biologically plausible association; coefficient direction and magnitude monitored for confounding with SSA geography (see S Section D). EXCLUDED from stunting models: Spearman r=+0·51 (p<0·001) with stunting but r=−0·57 with SDI, indicating strong collinearity with SDI in the stunting covariate set. Including HIV in stunting models alongside SDI would introduce severe multicollinearity (confirmed by Spearman HIV–SDI r=−0·57). |
| **Malaria** | +0.462 (***) | +0.702 (***) | 2.15 | None | **INCLUDED stunting EXCLUDED wasting** | MAP/Pf PR spatiotemporal model uses environmental covariates only — no child anthropometry. No circularity. INCLUDED in stunting: r=+0·70 (p<0·001), VIF 2·15; biologically relevant (immune competition, micronutrient depletion). EXCLUDED from wasting: r=+0·46 (p<0·001) but collinear with SDI (r=−0·77) and with SSA staple geography, absorbing staple exposure variance. Consistent with five-step covariate selection framework. |
| **TB** | +0.202 (*) | +0.368 (***) | 1.54 | Moderate | **EXCLUDED both models** | DisMod-MR prevalence model uses adult underweight prevalence as a direct covariate (Table 2, GBD 2023 Appendix 1 p 1225), creating an indirect circularity pathway via shared nutritional ecology with child wasting/stunting. Additionally, TB is collinear with LRI (Spearman r=+0·37, p<0·001) — retaining both would absorb staple exposure variance and introduce double-counting of respiratory infection burden. Low additional predictive value: r=+0·20 wasting (p=0·02), r=+0·37 stunting (p<0·001). Present in dataset (TB_prev) but excluded from all models. |
| **Nematodes** | +0.003 (ns) | +0.267 (**) | 1.26 | Moderate | **EXCLUDED both models** | ST-GPR prevalence model uses WASH SEVs and SDI as upstream inputs — sharing upstream structure with excluded WASH SEV variables. Note: the GBD nematode sequela sub-model separately uses the wasting envelope to estimate severe wasting due to ascariasis (GBD 2023 Appendix 1 p 179), but this applies to a downstream sequela attribution and does not affect the primary nematode prevalence estimate used here. Exclusion is primarily on statistical grounds: negligible signal with wasting (r=+0·00, p=0·97); low signal with stunting (r=+0·27, p=0·00). Collinear with TB (r=+0·39, p<0·001) and shares WASH/SDI upstream structure with excluded variables. Low VIF (1·26) but excluded on signal and collinearity grounds. |
| **UHC** | -0.461 (***) | -0.426 (***) | 2.06 | None | **INCLUDED both models** | Independent composite index absent from all GBD DisMod-MR and CODEm cause-specific models. No circularity pathway. Confirmed by systematic search of GBD 2023 Appendix 1. Significant negative association with both outcomes (r=−0·46 wasting, r=−0·43 stunting, both p<0·001). VIF 2·06 acceptable. Retained as the confirmed circularity-free healthcare access proxy, preferred over HAQ which is excluded on circularity grounds. Consistent negative coefficient across all six staple models in both outcomes. |
| **SDI** | -0.465 (***) | -0.764 (***) | 6.96⚠ | Low–moderate | **INCLUDED stunting EXCLUDED wasting** | EXCLUDED from wasting models: relationship between SDI and wasting is non-monotonic at country level. Higher-SDI South and Southeast Asian countries carry disproportionately high wasting burdens inconsistent with a linear development–wasting pathway, as demonstrated in Indian subnational data (see S Section B). INCLUDED in stunting models: strong negative association (r=−0·76, p<0·001), monotonic, VIF 6·96 acceptable given its role as the primary development covariate. SDI is a Level 3 covariate in LRI and diarrhoea DisMod-MR models; since LRI is also a stunting covariate, their effects are not fully independent — acknowledged as limitation. |
| **HAQ** | -0.445 (***) | -0.709 (***) | 6.92⚠ | High | **EXCLUDED both models** | GBD composite index constructed from 32 cause-specific mortality rates including conditions strongly associated with child undernutrition. Direct circularity pathway with JME wasting and stunting outcomes. Additionally, extremely collinear with SDI (r=+0·87, p<0·001) and WASH variables. VIF 6·92 in full candidate set. Excluded on circularity grounds; UHC retained as the circularity-free healthcare access proxy. |
| **WASH sanitation** | +0.427 (***) | +0.668 (***) | 5.12⚠ | Moderate | **EXCLUDED both models** | ST-GPR modelled using SDI and sanitation survey data. No direct child anthropometry input, but shares upstream structure with excluded variables. Strong signal (r=+0·43 wasting, r=+0·67 stunting) but collinear with SDI (r=−0·84), HAQ (r=−0·83), and WASH water (r=+0·79). VIF 5·12 exceeds threshold. Including WASH alongside SDI and UHC would introduce severe multicollinearity given limited EPV (n=127/126 with 6–7 fixed covariates). Excluded on multicollinearity grounds. |
| **WASH water** | +0.474 (***) | +0.672 (***) | 4.03 | Moderate | **EXCLUDED both models** | Same rationale as WASH sanitation. Collinear with sanitation (r=+0·79), SDI (r=−0·83), HAQ (r=−0·81). VIF 4·03. WASH sanitation and water measure the same underlying poverty-infrastructure construct and cannot be included simultaneously. Both excluded on multicollinearity grounds. |
| **Animal protein** | -0.462 (***) | -0.719 (***) | 2.62 | None | **INCLUDED both models** | FAOSTAT supply-side estimate — absent from all GBD modelling frameworks. No circularity. Significant negative association with both outcomes (r=−0·46 wasting, r=−0·72 stunting, both p<0·001). VIF 2·62. Primary dietary quality covariate; mechanistically linked to DIAAS-based protein quality and zinc bioavailability pathways. Retained as fixed covariate across all six staple models and all five sensitivity analyses. Entered alongside staple variables to isolate staple-specific associations from overall dietary quality. |
| **Total kcal** | -0.412 (***) | -0.701 (***) | 3.00 | None | **INCLUDED both models** | No circularity — FAOSTAT supply-side estimate absent from all GBD modelling frameworks. Significant negative association with both outcomes (r=−0·41 wasting, r=−0·70 stunting, both p<0·001). VIF 3·00 acceptable. INCLUDED as a fixed covariate in all models (wasting and stunting) to control for overall energy availability independently of protein quality. Moderate collinearity with animal protein (r=+0·70) and SDI (r=+0·70) acknowledged; all three retain distinct coefficients in model capturing separable dimensions: total kcal = energy sufficiency; animal protein = dietary protein quality; SDI = socioeconomic development. |
| **Non-cereal protein** | +0.113 (ns) | +0.193 (*) | 1.49 | None | **EXCLUDED both models** | No circularity. Negligible signal with wasting (r=+0·11, p=0·21); low signal with stunting (r=+0·19, p=0·03). Collinear with animal protein (r=−0·29, p<0·001). Positive direction of wasting association reflects ecological confounding — high non-cereal plant protein countries are predominantly lower-SDI agricultural economies. VIF 1·49 acceptable but excluded on signal and redundancy grounds. |

n=127 wasting; n=126 stunting. β = unstandardised coefficient; SE = standard error.

Significance: ***p<0·001, **p<0·01, *p<0·05, †p<0·10.

For wasting, rice showed a positive statistically significant relationship (β=+0·086, p=0·004), and sorghum also had a positive statistically significant relationship (β=+0·361, p<0·001), while millet had a positive relationship of borderline significance (β=+0·190, p=0·073). The relationship with maize was statistically significant and negative (β=−0·139, p=0·004). UHC index showed a consistent negative and significant relationship with all staples. LRI prevalence showed a consistent positive relationship to wasting which was significant with wheat, millet, sorghum and cassava while being of borderline significance with rice and maize. Diarrhoea did not show a statistically significant relationship with wasting for any staple, with direction inconsistently negative with millet and sorghum, while it was positive for the other staples. HIV prevalence showed a biologically implausible negative statistically significant relationship with all staples. Animal protein showed the expected negative relationship with all staples, which was statistically significant. For total energy supply, the direction of association with wasting was expectedly negative but not statistically significant for any staple.

**S Table 9. Complete robust MM regression results — primary analyses**

**Panel A. Wasting (n=127)**

| **Variable** | **Wheat** | | | **Rice** | | | **Maize** | | | **Millet** | | | **Sorghum** | | | **Cassava** | | |
| --- | --- | --- | --- | --- | --- | --- | --- | --- | --- | --- | --- | --- | --- | --- | --- | --- | --- | --- |
|  | **β** | **SE** | **p** | **β** | **SE** | **p** | **β** | **SE** | **p** | **β** | **SE** | **p** | **β** | **SE** | **p** | **β** | **SE** | **p** |
| Staple (protein g/d) | −0·028 | 0·023 | 0·222 | +0·086 | 0·029 | 0·004 ** | −0·139 | 0·047 | 0·004 ** | +0·190 | 0·105 | 0·073 † | +0·361 | 0·100 | <0·001 *** | +0·039 | 0·160 | 0·809 |
| UHC index | −0·063 | 0·023 | 0·008 ** | −0·059 | 0·022 | 0·009 ** | −0·052 | 0·023 | 0·028 * | −0·058 | 0·024 | 0·015 * | −0·050 | 0·024 | 0·043 * | −0·065 | 0·024 | 0·009 ** |
| LRI prevalence (%) | +12·555 | 5·189 | 0·017 * | +9·253 | 4·959 | 0·065 † | +9·189 | 5·178 | 0·079 † | +11·773 | 5·157 | 0·024 * | +13·553 | 5·259 | 0·011 * | +12·055 | 5·354 | 0·026 * |
| Diarrhoea prevalence (%) | +0·029 | 0·440 | 0·947 | +0·111 | 0·413 | 0·788 | +0·128 | 0·431 | 0·767 | −0·054 | 0·441 | 0·902 | −0·326 | 0·456 | 0·476 | +0·102 | 0·451 | 0·821 |
| HIV prevalence (%) | −2·880 | 0·779 | <0·001 *** | −2·340 | 0·754 | 0·002 ** | −2·093 | 0·821 | 0·012 * | −2·788 | 0·779 | <0·001 *** | −2·845 | 0·791 | <0·001 *** | −2·906 | 0·802 | <0·001 *** |
| Animal protein (g/d) | −0·044 | 0·017 | 0·010 ** | −0·033 | 0·016 | 0·045 * | −0·058 | 0·017 | <0·001 *** | −0·041 | 0·017 | 0·018 * | −0·035 | 0·017 | 0·047 * | −0·044 | 0·018 | 0·016 * |
| Total energy (kcal/d) | −0·000 | 0·001 | 0·901 | −0·001 | 0·001 | 0·288 | −0·001 | 0·001 | 0·326 | −0·001 | 0·001 | 0·410 | −0·001 | 0·001 | 0·379 | −0·001 | 0·001 | 0·495 |
| Model R² | 0·361 | | | 0·391 | | | 0·393 | | | 0·368 | | | 0·406 | | | 0·350 | | |

*SDI excluded from all wasting models — see S Table 9.*

For stunting (Panel B, S Table 10) no staple protein term reached conventional significance, though cassava approached borderline significance (β=+0·975, p=0·092). SDI showed a negative association with stunting across all six staple models, reaching statistical significance for wheat (β=−24·992, p=0·008), rice (β=−26·950, p=0·003), maize (β=−25·442, p=0·005), millet (β=−26·685, p=0·011), sorghum (β=−23·317, p=0·016) and cassava (β=−29·950, p=0·001). UHC index showed no significant association with stunting in any model. LRI prevalence showed a consistent, strongly significant positive association with stunting across all staple models (coefficients +63·5 to +67·1, all p<0·001). Diarrhoea and malaria did not show any significant relationship with stunting in any model. Animal protein showed a negative association with stunting that was statistically significant for rice (p=0·044) and of borderline significance for wheat (p=0·056), maize (p=0·088), millet (p=0·064) and sorghum (p=0·060); the association was not significant for cassava. Total energy intake showed a consistent negative association with stunting across all staple models (coefficients −0·005 to −0·006, all p≤0·032).

**S Table 9. Complete robust MM regression results — primary analyses**

**Panel B. Stunting (n=126)**

| **Variable** | **Wheat** | | | **Rice** | | | **Maize** | | | **Millet** | | | **Sorghum** | | | **Cassava** | | |
| --- | --- | --- | --- | --- | --- | --- | --- | --- | --- | --- | --- | --- | --- | --- | --- | --- | --- | --- |
|  | **β** | **SE** | **p** | **β** | **SE** | **p** | **β** | **SE** | **p** | **β** | **SE** | **p** | **β** | **SE** | **p** | **β** | **SE** | **p** |
| Staple (protein g/d) | −0·040 | 0·067 | 0·553 | −0·075 | 0·084 | 0·379 | +0·124 | 0·128 | 0·333 | −0·025 | 0·342 | 0·943 | +0·249 | 0·279 | 0·374 | +0·975 | 0·573 | 0·092 † |
| SDI | −24·992 | 9·241 | 0·008 ** | −26·950 | 9·018 | 0·003 ** | −25·442 | 8·987 | 0·005 ** | −26·685 | 10·314 | 0·011 * | −23·317 | 9·535 | 0·016 * | −29·950 | 9·032 | 0·001 ** |
| UHC index | +0·065 | 0·073 | 0·380 | +0·069 | 0·073 | 0·346 | +0·058 | 0·073 | 0·430 | +0·070 | 0·073 | 0·343 | +0·074 | 0·073 | 0·317 | +0·070 | 0·071 | 0·331 |
| LRI prevalence (%) | +66·189 | 15·475 | <0·001 *** | +65·863 | 15·445 | <0·001 *** | +66·647 | 15·379 | <0·001 *** | +64·985 | 15·589 | <0·001 *** | +67·103 | 15·635 | <0·001 *** | +63·464 | 15·133 | <0·001 *** |
| Diarrhoea prevalence (%) | +0·018 | 1·204 | 0·988 | +0·083 | 1·204 | 0·945 | +0·104 | 1·197 | 0·931 | +0·054 | 1·219 | 0·965 | −0·236 | 1·247 | 0·850 | +0·507 | 1·209 | 0·676 |
| Malaria prevalence (%) | +0·102 | 0·094 | 0·281 | +0·099 | 0·094 | 0·295 | +0·113 | 0·092 | 0·223 | +0·110 | 0·094 | 0·247 | +0·110 | 0·093 | 0·240 | −0·025 | 0·122 | 0·840 |
| Animal protein (g/d) | −0·105 | 0·054 | 0·056 † | −0·112 | 0·055 | 0·044 * | −0·093 | 0·054 | 0·088 † | −0·102 | 0·054 | 0·064 † | −0·103 | 0·054 | 0·060 † | −0·082 | 0·054 | 0·133 |
| Total energy (kcal/d) | −0·005 | 0·002 | 0·032 * | −0·005 | 0·002 | 0·022 * | −0·005 | 0·002 | 0·020 * | −0·006 | 0·002 | 0·020 * | −0·006 | 0·002 | 0·011 * | −0·005 | 0·002 | 0·017 * |
| Model R² | 0·699 | | | 0·701 | | | 0·701 | | | 0·698 | | | 0·700 | | | 0·709 | | |

*SDI significant across all six stunting models, β range −23·7 to −29·9, all p<0·01.*

**S Table 10. Quantile regression — staple protein associations**

τ = quantile. β = unstandardised coefficient; SE = standard error. n=127 wasting; n=126 stunting.

Significance: ***p<0·001, **p<0·01, *p<0·05, †p<0·10. Sensitivity analyses used robust MM only; QR not applied to S1–S5.

**Panel A. Wasting**

| **Staple** | **Q25 (τ=0·25)** | | | **Q50 (τ=0·50)** | | | **Q75 (τ=0·75)** | | |
| --- | --- | --- | --- | --- | --- | --- | --- | --- | --- |
|  | **β** | **SE** | **p** | **β** | **SE** | **p** | **β** | **SE** | **p** |
| Wheat | −0·029 | 0·025 | 0·252 | −0·037 | 0·030 | 0·216 | +0·043 | 0·059 | 0·467 |
| Rice | +0·075 | 0·024 | 0·003 ** | +0·050 | 0·047 | 0·296 | +0·085 | 0·061 | 0·167 |
| Maize | −0·108 | 0·040 | 0·008 ** | −0·134 | 0·060 | 0·027 * | −0·259 | 0·079 | 0·001 ** |
| Millet | +0·309 | 0·263 | 0·242 | +0·248 | 0·351 | 0·481 | +0·415 | 0·635 | 0·514 |
| Sorghum | +0·265 | 0·161 | 0·101 | +0·410 | 0·216 | 0·060 † | +0·546 | 0·308 | 0·079 † |
| Cassava | +0·291 | 0·167 | 0·084 † | +0·130 | 0·154 | 0·401 | −0·121 | 0·287 | 0·673 |

**Panel B. Stunting**

| **Staple** | **Q25 (τ=0·25)** | | | **Q50 (τ=0·50)** | | | **Q75 (τ=0·75)** | | |
| --- | --- | --- | --- | --- | --- | --- | --- | --- | --- |
|  | **β** | **SE** | **p** | **β** | **SE** | **p** | **β** | **SE** | **p** |
| Wheat | −0·100 | 0·074 | 0·180 | −0·078 | 0·092 | 0·394 | −0·046 | 0·109 | 0·673 |
| Rice | −0·055 | 0·130 | 0·674 | −0·157 | 0·115 | 0·176 | −0·132 | 0·117 | 0·260 |
| Maize | +0·361 | 0·185 | 0·053 † | +0·230 | 0·183 | 0·211 | +0·371 | 0·257 | 0·151 |
| Millet | −0·271 | 0·653 | 0·678 | −0·601 | 0·837 | 0·474 | +0·252 | 0·965 | 0·794 |
| Sorghum | −0·321 | 0·483 | 0·507 | +0·841 | 0·673 | 0·214 | +0·515 | 0·576 | 0·373 |
| Cassava | +0·884 | 0·715 | 0·218 | +1·079 | 1·059 | 0·310 | +1·455 | 1·116 | 0·195 |

In the quantile regression analysis for wasting, rice showed a statistically significant positive association at Q0·25 (β=+0·075, p=0·002), with consistent positive but non-significant associations at Q0·50 and Q0·75. Maize showed the most consistent negative association across all three quantiles, strengthening progressively: Q0·25 (β=−0·108, p=0·008), Q0·50 (β=−0·134, p=0·027), Q0·75 (β=−0·259, p=0·001). Sorghum showed a consistent positive association increasing across quantiles — Q0·25 (β=+0·265, p=0·101), Q0·50 (β=+0·410, p=0·060†), Q0·75 (β=+0·546, p=0·079†) — suggesting the association is strongest among countries with the highest wasting burden. Millet showed consistently positive but non-significant associations at all three quantiles (β=+0·310, +0·248, +0·415), with no clear attenuation pattern. Cassava showed a borderline positive association at Q0·25 (β=+0·291, p=0·084†), attenuating to non-significant at Q0·50 and reversing at Q0·75 (β=−0·121); the large standard errors across all cassava quantiles indicate instability due to the limited number of cassava-dominant countries. Wheat did not show any significant association at any quantile. For stunting, none of the staples showed any statistically significant association at any quantile, with the exception of maize at Q0·25, which showed a borderline positive association (β=+0·362, SE=0·185, p=0·054†). This positive direction at Q0·25 is opposite to the consistently negative wasting association for maize and is consistent with the hypothesis that maize-dominant diets — characterised by high phytate:zinc molar ratios (S Table 15) and low zinc bioavailability — may impair linear growth through zinc deficiency rather than protein deficit. Martorell and Young, in a comparison of India and Guatemala using nationally representative survey data, documented that 67·5% of Guatemalan women had height below 150 cm (mean 147·7 cm) — substantially shorter than Indian women (mean 151·6 cm) — despite Guatemala having near-identical stunting prevalence (48%) in children under 5 years. They noted that short maternal stature in Guatemala reflects generational linear growth failure, and that Guatemala shares with nearly all other Latin American countries (except Haiti) the near-absence of wasting as a public health problem — a pattern consistent with maize’s high phytate:zinc ratio driving stunting while its moderate protein supply is sufficient to maintain lean mass. The positive association of maize supply with stunting at Q0·25 in the present analysis may reflect this zinc-mediated stunting pathway operating at the population level, distinct from the wasting pathway addressed by the primary analysis. The contrast between the sorghum–wasting signal (strong, p<0·001) and the maize–stunting signal (positive but non-significant, p=0·333) merits explanation, as both are predicted by the same framework. Examination of the top 10 maize-consuming countries reveals two distinct subgroups with divergent nutritional profiles. The Sub-Saharan African group (Malawi 2·6%/35·5%, Lesotho 1·6%/35·6%, Zambia 4·2%/34·6%, Eswatini 1·8%/20·0%) shows the predicted pattern: very low wasting and high stunting consistent with the zinc deficiency mechanism. Guatemala (0·8%/46·0%) similarly exemplifies the classic maize–stunting profile documented by Martorell and Young. However, the Latin American subgroup includes Mexico (1·0%/12·5%, SDI=0·693) and Paraguay (1·0%/5·6%, SDI=0·674) — high-SDI countries where animal protein intake (55·5 and 30·5 g/capita/day respectively) and healthcare access appear to mitigate the zinc-phytate stunting mechanism despite high maize consumption. This compositional heterogeneity — low-SDI SSA countries with high stunting alongside higher-SDI Latin American countries with low stunting, all consuming similar maize quantities — dilutes the regression signal after covariate adjustment (partial r=+0·111, p=0·215 after SDI adjustment). By contrast, the top 10 sorghum consumers are an almost homogeneous group: all are low-SDI Sahelian or Horn of Africa countries (SDI range 0·20–0·51, mean 0·38), all with high wasting prevalence (4·3–22·7%) and low animal protein supply (7·0–27·8 g/capita/day, mean 14·6 g/d). This homogeneity produces a strong, unconfounded regression signal. The biology of the maize–stunting relationship is likely real — as the individual country profiles confirm — but is not detectable as a statistically significant ecological signal in a heterogeneous global sample of 126 countries after adjusting for SDI and other covariates.

For fixed covariates with wasting in quantile regression, LRI prevalence showed a positive association across all staples and quantiles, reaching statistical significance at Q0·50 for wheat (β=+15·125, p=0·006**), rice (β=+11·518, p=0·040*) and cassava (β=+13·712, p=0·015*); the association was not significant for maize, millet or sorghum at any quantile. At Q0·75 the association was borderline for wheat only (β=+22·166, p=0·088†). At Q0·25 the association was borderline for cassava (β=+10·436, p=0·055†) and non-significant for all other staples. UHC index showed a negative association with wasting that was statistically significant at Q0·25 for wheat (p=0·010*), rice (p=0·001**) and cassava (p=0·044*), and borderline for maize (p=0·091†); millet and sorghum showed no significant association at any quantile. At Q0·50 the UHC association was borderline for wheat (p=0·060†) and cassava (p=0·053†), and non-significant for the remaining staples. At Q0·75 the UHC association was significant for rice (p=0·035*) and borderline for cassava (p=0·055†). HIV prevalence showed no statistically significant association with wasting for any staple at any quantile; a borderline negative association was observed for wheat at Q0·75 (β=−4·638, p=0·076†). Animal protein supply showed a significant negative association with wasting at Q0·25 for wheat (p=0·025*) and maize (p=0·002**), and at Q0·50 for maize (p=0·046*); borderline associations were observed at Q0·25 for rice (p=0·076†) and at Q0·50 for wheat (p=0·092†) and at Q0·75 for maize (p=0·085†). Millet, sorghum and cassava showed no significant animal protein association at any quantile. Total energy supply showed no significant association with wasting at Q0·25 for any staple; borderline associations emerged at Q0·50 for rice (p=0·092†) and maize (p=0·068†), and at Q0·75 for wheat (p=0·066†) and rice (p=0·062†).

For fixed covariates with stunting, LRI prevalence showed a consistent positive association across all staples and quantiles, reaching statistical significance (p<0·05) for all staples at Q0·50 and at Q0·25 for wheat, rice and maize; the association was borderline for millet, sorghum and cassava at Q0·25, and for cassava at Q0·75. SDI showed a negative association with stunting that was statistically significant for rice, maize, millet and sorghum at Q0·25 and Q0·50, borderline for wheat at Q0·50 (p=0·076), and significant across all quantiles for cassava. Animal protein did not show any statistically significant association with stunting for any staple. Total energy availability showed a statistically significant negative association with stunting at Q0·75 for all six staples. At Q0·25 the association was significant for rice and cassava, and borderline for maize, millet and sorghum. At Q0·50 the association was significant for sorghum and cassava only.

**S Table 11. Quantile regression — fixed covariate associations**

Selected covariates shown across three quantiles for each staple model. β = unstandardised coefficient.

**Panel A. Wasting — fixed covariates by quantile**

| **Variable / Quantile** | **Wheat** | | **Rice** | | **Maize** | | **Millet** | | **Sorghum** | | **Cassava** | |
| --- | --- | --- | --- | --- | --- | --- | --- | --- | --- | --- | --- | --- |
|  | **β** | **p** | **β** | **p** | **β** | **p** | **β** | **p** | **β** | **p** | **β** | **p** |
| LRI prevalence (%) (Q25) | +8·139 | 0·171 | +5·155 | 0·279 | +6·110 | 0·190 | +7·600 | 0·154 | +7·791 | 0·161 | +9·902 | 0·045 * |
| LRI prevalence (Q50) | +15·125 | 0·006 ** | +11·518 | 0·046 * | +6·322 | 0·309 | +11·672 | 0·039 * | +12·523 | 0·040 * | +11·971 | 0·047 * |
| LRI prevalence (%) (Q75) | +22·166 | 0·100 † | +9·549 | 0·468 | +15·151 | 0·202 | +13·160 | 0·311 | +17·164 | 0·162 | +16·847 | 0·185 |
| UHC index (Q25) | −0·056 | 0·010 ** | −0·063 | 0·001 ** | −0·041 | 0·084 † | −0·055 | 0·012 * | −0·037 | 0·103 | −0·042 | 0·047 * |
| UHC index (Q50) | −0·060 | 0·057 † | −0·046 | 0·142 | −0·030 | 0·389 | −0·038 | 0·216 | −0·041 | 0·166 | −0·040 | 0·232 |
| UHC index (Q75) | −0·078 | 0·156 | −0·111 | 0·029 * | −0·074 | 0·178 | −0·075 | 0·145 | −0·066 | 0·191 | −0·075 | 0·142 |
| HIV prevalence (%) (Q25) | −1·492 | 0·296 | −1·154 | 0·287 | −1·126 | 0·255 | −1·502 | 0·243 | −1·402 | 0·296 | −1·612 | 0·143 |
| HIV prevalence (Q50) | −2·453 | 0·187 | −2·265 | 0·222 | −1·676 | 0·264 | −2·221 | 0·197 | −2·351 | 0·221 | −2·241 | 0·194 |
| HIV prevalence (%) (Q75) | −4·638 | 0·102 | −3·579 | 0·209 | −2·772 | 0·239 | −3·625 | 0·204 | −3·917 | 0·173 | −3·907 | 0·202 |
| Animal protein (g/d) (Q25) | −0·039 | 0·020 * | −0·027 | 0·080 † | −0·049 | 0·003 ** | −0·034 | 0·055 † | −0·013 | 0·499 | −0·022 | 0·202 |
| Animal protein (Q50) | −0·032 | 0·092 † | −0·024 | 0·220 | −0·046 | 0·044 * | −0·029 | 0·107 | −0·020 | 0·313 | −0·027 | 0·199 |
| Animal protein (g/d) (Q75) | −0·026 | 0·469 | −0·011 | 0·740 | −0·053 | 0·070 † | −0·019 | 0·582 | −0·004 | 0·921 | −0·041 | 0·286 |

**Panel B. Stunting — fixed covariates by quantile**

| **Variable / Quantile** | **Wheat** | | **Rice** | | **Maize** | | **Millet** | | **Sorghum** | | **Cassava** | |
| --- | --- | --- | --- | --- | --- | --- | --- | --- | --- | --- | --- | --- |
|  | **β** | **p** | **β** | **p** | **β** | **p** | **β** | **p** | **β** | **p** | **β** | **p** |
| LRI prevalence (%) (Q25) | +56·209 | 0·010 * | +54·438 | 0·014 * | +47·215 | 0·012 * | +38·664 | 0·074 † | +40·251 | 0·083 † | +40·296 | 0·057 † |
| LRI prevalence (Q50) | +68·178 | 0·002 ** | +60·980 | 0·002 ** | +66·114 | 0·003 ** | +60·293 | 0·007 ** | +54·451 | 0·012 * | +66·830 | 0·002 ** |
| LRI prevalence (%) (Q75) | +93·612 | 0·005 ** | +76·062 | 0·029 * | +84·049 | 0·025 * | +89·833 | 0·008 ** | +91·429 | 0·007 ** | +60·144 | 0·058 † |
| SDI (Q25) | −12·438 | 0·363 | −27·648 | 0·019 * | −23·016 | 0·028 * | −29·364 | 0·016 * | −31·242 | 0·015 * | −28·109 | 0·009 ** |
| SDI (Q50) | −32·611 | 0·076 † | −45·857 | 0·010 ** | −37·154 | 0·020 * | −39·965 | 0·014 * | −36·819 | 0·015 * | −33·158 | 0·046 * |
| SDI (Q75) | −25·071 | 0·158 | −29·032 | 0·126 | −27·953 | 0·154 | −24·761 | 0·200 | −22·498 | 0·211 | −45·778 | 0·011 * |
| Animal protein (g/d) (Q25) | −0·115 | 0·115 | −0·064 | 0·373 | −0·078 | 0·215 | −0·090 | 0·212 | −0·065 | 0·364 | −0·054 | 0·438 |
| Animal protein (Q50) | −0·039 | 0·689 | −0·065 | 0·464 | −0·014 | 0·878 | −0·049 | 0·609 | −0·037 | 0·687 | −0·030 | 0·755 |
| Animal protein (g/d) (Q75) | +0·009 | 0·938 | +0·042 | 0·736 | +0·050 | 0·675 | +0·045 | 0·696 | +0·045 | 0·697 | +0·108 | 0·380 |
| Total energy (kcal/d) (Q25) | −0·002 | 0·464 | −0·004 | 0·026 * | −0·003 | 0·066 † | −0·004 | 0·066 † | −0·004 | 0·089 † | −0·005 | 0·015 * |
| Total energy (Q50) | −0·004 | 0·227 | −0·003 | 0·265 | −0·004 | 0·142 | −0·003 | 0·211 | −0·005 | 0·039 * | −0·006 | 0·048 * |
| Total energy (kcal/d) (Q75) | −0·010 | 0·031 * | −0·012 | 0·003 ** | −0·010 | 0·015 * | −0·012 | 0·007 ** | −0·013 | 0·003 ** | −0·010 | 0·026 * |

Five sensitivity analyses were conducted for wasting: S1 excluding the five highest-wasting countries (South Sudan, India, Yemen, Sudan, Mauritius; n=122); S2 excluding 11 countries with non-FAOSTAT staple data (nine FAOSTAT-masked countries plus Eritrea and Equatorial Guinea; n=118); S3 restricted to Sub-Saharan and North African countries (n=47); S4 excluding countries above the Q75 HIV prevalence threshold (n=95); and S5 population-weighted analysis using UN IGME 2023 under-5 population weights (n=127). Wheat showed a borderline negative association in S1 (β=−0·037, p=0·082†) and S2 (β=−0·040, p=0·075†), a statistically significant negative association in S5 (β=−0·059, p<0·001***), and non-significant in S4. In S3, wheat showed a strongly positive association (β=+0·344, p<0·001***) — a reversal of direction explained by a suppressor variable effect: the raw Spearman correlation between wheat and wasting within S3 is −0·135 (negative), but wheat is strongly correlated with UHC (r=+0·395) and animal protein (r=+0·586) in this subset, both of which drive down wasting. After controlling for these covariates, the residual partial coefficient turns positive — a statistical artefact of multicollinearity in the geographically restricted subset, not a biological effect of wheat protein quality (see further discussion below). Rice showed a consistent positive association with wasting, statistically significant in S1 (β=+0·092, p=0·001**), S2 (β=+0·103, p<0·001***) and S5 (β=+0·044, p=0·028*), and of borderline significance in S4 (β=+0·068, p=0·062†); non-significant in S3. Maize showed a consistent negative association across all five analyses: statistically significant in S1 (β=−0·128, p=0·005**), S2 (β=−0·137, p=0·004**) and S5 (β=−0·131, p<0·001***), borderline in S4 (β=−0·130, p=0·069†), and non-significant in S3 (negative direction maintained; β=−0·172, p=0·112). Millet showed a borderline positive association with wasting in S1 (β=+0·191, p=0·055†) and S2 (β=+0·194, p=0·056†), a statistically significant positive association in S4 (β=+0·287, p=0·044*) and S5 (β=+0·208, p=0·001***), and non-significant in S3. Sorghum showed a statistically significant positive association across all five analyses: S1 (β=+0·221, p=0·038*), S2 (β=+0·217, p=0·046*), S3 (β=+0·404, p=0·008**), S4 (β=+0·462, p<0·001***) and S5 (β=+0·249, p<0·001***) — the most robust finding across all sensitivity specifications. Cassava showed no significant association in S1–S4; the association was positive and statistically significant only in S5 (β=+0·278, p<0·001***).

For stunting, wheat did not show a statistically significant association in any sensitivity analysis. Rice showed a negative statistically significant association with stunting in S3 (β=−0·428, p=0·014*) and S5 (β=−0·133, p=0·012*), consistent with the direction of the primary analysis. Maize showed a statistically significant positive association with stunting in S5 only (β=+0·195, p=0·022*); non-significant in all other specifications. Millet showed a non-significant association with stunting in S1–S4; in S5 it showed a strongly positive statistically significant association (β=+2·954, p<0·001***) — the largest coefficient in the entire study, reflecting the population-weighted dominance of high-millet, high-stunting Sahelian countries. Sorghum showed a statistically significant positive association with stunting in S3 (β=+0·655, p=0·032*) and S5 (β=+0·715, p<0·001***), borderline in S4 (β=+0·606, p=0·071†), and non-significant in S1–S2. Cassava showed a statistically significant positive association with stunting in S1 (β=+1·168, p=0·035*) and S5 (β=+1·134, p<0·001***), borderline in S2 (β=+0·984, p=0·089†), and non-significant in S3–S4, consistent with the borderline significance seen in the primary analysis. LRI prevalence showed a robust, consistent positive association with stunting across all staple models in S1–S2 (all p≤0·013) and S4–S5 (all p≤0·002); in S3 the association was significant for most staples (wheat, maize, cassava p<0·050) though borderline for rice and non-significant for millet and sorghum. For wasting, LRI showed a positive significant or borderline association with most staples in S1 (significant for wheat, sorghum, cassava; borderline for rice and maize; non-significant for millet) and S3 (significant for all six staples, p≤0·050 or borderline†). In S2, LRI was significant for wheat (p=0·022*) and sorghum (p=0·028*) only, non-significant for the remaining four staples. In S4, LRI was non-significant for all staples with wasting. In S5, LRI was significant for wheat (p<0·001***), rice (p=0·002**), sorghum (p=0·017*) and cassava (p<0·001***), and of borderline significance for maize (p=0·094†) and millet (p=0·083†).

**S Table 12. Sensitivity analyses — staple protein associations (complete)**

Robust MM regression. β = unstandardised coefficient; SE = standard error. Significance: ***p<0·001, **p<0·01, *p<0·05, †p<0·10.

S1: excludes five highest-wasting countries (South Sudan, India, Yemen, Sudan, Mauritius), n=122 wasting/n=121 stunting. S2: excludes eleven FAOSTAT-masked countries, n=118 wasting/n=117 stunting. S3: sub-Saharan Africa plus North Africa, n=47 wasting/n=47 stunting. S4: excludes countries above Q75 HIV prevalence (0·089%), n=95 wasting/n=94 stunting. S5: population-weighted (Under-5 population), n=127 wasting/n=126 stunting.

**S Table 12.Panel A. Wasting results from S1 to S5 (with no of countries in parentheses)**

| **Staple** | **Primary (n=127)** | | | **S1 (n=122)** | | | **S2 (n=118)** | | | **S3 (n=47)** | | | **S4 (n=95)** | | | **S5 wtd (n=127)** | | |
| --- | --- | --- | --- | --- | --- | --- | --- | --- | --- | --- | --- | --- | --- | --- | --- | --- | --- | --- |
|  | **β** | **SE** | **p** | **β** | **SE** | **p** | **β** | **SE** | **p** | **β** | **SE** | **p** | **β** | **SE** | **p** | **β** | **SE** | **p** |
| Wheat | −0·028 | 0·023 | 0·222 | −0·037 | 0·021 | 0·082 † | −0·040 | 0·022 | 0·075 † | +0·344 | 0·084 | <0·001 *** | −0·028 | 0·026 | 0·283 | −0·058 | 0·017 | <0·001 *** |
| Rice | +0·086 | 0·029 | 0·004 ** | +0·092 | 0·027 | 0·001 ** | +0·103 | 0·029 | <0·001 *** | −0·113 | 0·088 | 0·206 | +0·068 | 0·036 | 0·062 † | +0·044 | 0·020 | 0·028 * |
| Maize | −0·139 | 0·047 | 0·004 ** | −0·128 | 0·045 | 0·005 ** | −0·137 | 0·046 | 0·004 ** | −0·172 | 0·106 | 0·112 | −0·130 | 0·071 | 0·069 † | −0·131 | 0·031 | <0·001 *** |
| Millet | +0·190 | 0·105 | 0·073 † | +0·191 | 0·099 | 0·055 † | +0·194 | 0·100 | 0·056 † | +0·089 | 0·154 | 0·567 | +0·287 | 0·140 | 0·044 * | +0·208 | 0·059 | <0·001 *** |
| Sorghum | +0·361 | 0·100 | <0·001 *** | +0·221 | 0·105 | 0·038 * | +0·217 | 0·107 | 0·046 * | +0·404 | 0·144 | 0·008 ** | +0·462 | 0·121 | <0·001 *** | +0·249 | 0·069 | <0·001 *** |
| Cassava | +0·039 | 0·160 | 0·809 | +0·090 | 0·150 | 0·549 | +0·081 | 0·155 | 0·603 | +0·032 | 0·262 | 0·904 | −0·222 | 0·350 | 0·528 | +0·278 | 0·071 | <0·001 *** |

**S Table 12.Panel B. Stunting**

| **Staple** | **Primary (n=126)** | | | **S1 (n=121)** | | | **S2 (n=117)** | | | **S3 (n=47)** | | | **S4 (n=94)** | | | **S5 wtd (n=126)** | | |
| --- | --- | --- | --- | --- | --- | --- | --- | --- | --- | --- | --- | --- | --- | --- | --- | --- | --- | --- |
|  | **β** | **SE** | **p** | **β** | **SE** | **p** | **β** | **SE** | **p** | **β** | **SE** | **p** | **β** | **SE** | **p** | **β** | **SE** | **p** |
| Wheat | −0·039 | 0·066 | 0·556 | −0·065 | 0·064 | 0·312 | −0·031 | 0·062 | 0·618 | −0·004 | 0·158 | 0·981 | −0·022 | 0·069 | 0·754 | −0·059 | 0·040 | 0·143 |
| Rice | −0·073 | 0·084 | 0·384 | −0·053 | 0·080 | 0·514 | −0·087 | 0·081 | 0·281 | −0·388 | 0·158 | 0·018 * | −0·108 | 0·095 | 0·259 | −0·137 | 0·051 | 0·008 ** |
| Maize | +0·121 | 0·127 | 0·345 | +0·174 | 0·121 | 0·154 | +0·208 | 0·119 | 0·082 † | +0·063 | 0·185 | 0·736 | +0·281 | 0·191 | 0·145 | +0·192 | 0·082 | 0·021 * |
| Millet | −0·025 | 0·342 | 0·943 | −0·255 | 0·342 | 0·458 | −0·130 | 0·353 | 0·712 | +0·521 | 0·401 | 0·202 | +0·297 | 0·411 | 0·473 | +2·954 | 0·499 | <0·001 *** |
| Sorghum | +0·249 | 0·279 | 0·374 | −0·203 | 0·348 | 0·561 | −0·174 | 0·364 | 0·635 | +0·655 | 0·294 | 0·032 * | +0·606 | 0·331 | 0·071 † | +0·715 | 0·142 | <0·001 *** |
| Cassava | +0·975 | 0·573 | 0·092 † | +1·168 | 0·547 | 0·035 * | +0·984 | 0·573 | 0·089 † | +1·067 | 0·704 | 0·138 | −0·328 | 1·012 | 0·747 | +1·134 | 0·225 | <0·001 *** |

**Panel C. LRI prevalence robustness — wasting models**

| **Staple** | **Primary (n=126)** | | | **S1 (n=121)** | | | **S2 (n=117)** | | | **S3 (n=47)** | | | **S4 (n=94)** | | | **S5 wtd (n=126)** | | |
| --- | --- | --- | --- | --- | --- | --- | --- | --- | --- | --- | --- | --- | --- | --- | --- | --- | --- | --- |
|  | **β** | **SE** | **p** | **β** | **SE** | **p** | **β** | **SE** | **p** | **β** | **SE** | **p** | **β** | **SE** | **p** | **β** | **SE** | **p** |
| Wheat | +12·403 | 5·167 | 0·018 * | +12·106 | 4·839 | 0·014 * | +9·128 | 5·394 | 0·093 † | +16·357 | 12·564 | 0·200 | +9·091 | 6·339 | 0·155 | +14·553 | 3·345 | <0·001 *** |
| Rice | +8·974 | 4·956 | 0·073 † | +8·270 | 4·688 | 0·080 † | +6·262 | 5·185 | 0·230 | +15·838 | 12·184 | 0·200 | +6·264 | 6·116 | 0·309 | +9·978 | 3·144 | 0·002 ** |
| Maize | +8·953 | 5·182 | 0·087 † | +8·420 | 4·907 | 0·089 † | +6·229 | 5·408 | 0·252 | +15·203 | 12·442 | 0·228 | +4·774 | 6·458 | 0·462 | +4·940 | 3·134 | 0·118 |
| Millet | +11·773 | 5·157 | 0·024 * | +11·035 | 4·845 | 0·025 * | +10·520 | 5·047 | 0·039 * | +28·102 | 13·435 | 0·043 * | +8·990 | 6·398 | 0·164 | +11·030 | 2·928 | <0·001 *** |
| Sorghum | +13·553 | 5·259 | 0·011 * | +12·267 | 4·795 | 0·012 * | +11·742 | 4·998 | 0·021 * | +28·083 | 13·283 | 0·041 * | +10·773 | 6·346 | 0·093 † | +12·201 | 3·107 | <0·001 *** |
| Cassava | +12·055 | 5·354 | 0·026 * | +11·505 | 5·028 | 0·024 * | +10·920 | 5·262 | 0·040 * | +29·525 | 14·047 | 0·042 * | +10·082 | 6·696 | 0·136 | +10·416 | 2·644 | <0·001 *** |

**Panel D. LRI prevalence robustness — stunting models**

| **Staple** | **Primary (n=126)** | | | **S1 (n=121)** | | | **S2 (n=117)** | | | **S3 (n=47)** | | | **S4 (n=94)** | | | **S5 wtd (n=126)** | | |
| --- | --- | --- | --- | --- | --- | --- | --- | --- | --- | --- | --- | --- | --- | --- | --- | --- | --- | --- |
|  | **β** | **SE** | **p** | **β** | **SE** | **p** | **β** | **SE** | **p** | **β** | **SE** | **p** | **β** | **SE** | **p** | **β** | **SE** | **p** |
| Wheat | +66·263 | 15·367 | <0·001 *** | +65·670 | 14·676 | <0·001 *** | +51·712 | 15·002 | <0·001 *** | +56·446 | 29·230 | 0·060 † | +70·349 | 18·024 | <0·001 *** | +63·622 | 8·262 | <0·001 *** |
| Rice | +65·959 | 15·334 | <0·001 *** | +64·282 | 14·680 | <0·001 *** | +51·225 | 14·857 | <0·001 *** | +41·524 | 27·683 | 0·141 | +71·716 | 17·935 | <0·001 *** | +55·035 | 9·337 | <0·001 *** |
| Maize | +66·683 | 15·276 | <0·001 *** | +65·845 | 14·553 | <0·001 *** | +52·548 | 14·534 | <0·001 *** | +54·759 | 29·008 | 0·066 † | +77·831 | 18·416 | <0·001 *** | +56·211 | 8·049 | <0·001 *** |
| Millet | +64·985 | 15·589 | <0·001 *** | +62·388 | 14·831 | <0·001 *** | +59·926 | 16·019 | <0·001 *** | +71·444 | 30·420 | 0·024 * | +70·181 | 17·994 | <0·001 *** | +30·219 | 8·166 | <0·001 *** |
| Sorghum | +67·103 | 15·635 | <0·001 *** | +62·169 | 14·980 | <0·001 *** | +59·127 | 16·154 | <0·001 *** | +77·725 | 28·381 | 0·009 ** | +73·313 | 17·819 | <0·001 *** | +38·089 | 8·849 | <0·001 *** |
| Cassava | +63·464 | 15·133 | <0·001 *** | +61·888 | 14·478 | <0·001 *** | +59·030 | 15·517 | <0·001 *** | +69·321 | 29·725 | 0·025 * | +70·065 | 18·112 | <0·001 *** | +60·467 | 7·709 | <0·001 *** |

*S4 excludes countries above Q75 HIV prevalence (0·094%) to test HIV confounding; HIV coefficient reverses from negative to positive in S4, confirming geographic confounding (see S Table 13F below for supporting country-level detail). S3 includes all African UN subregions (n=52).*

HIV prevalence showed a strongly negative and statistically significant association with wasting in all primary staple models (β=−2·093 to −2·906, all p≤0·012). This is biologically implausible as a causal relationship. The pattern reflects geographic confounding: the highest HIV burden in the dataset is concentrated in maize-dominant countries of southern and eastern sub-Saharan Africa (Lesotho, Eswatini, Mozambique, Kenya, Tanzania, Zimbabwe, South Africa, Malawi; Spearman r=+0·30 between HIV prevalence and maize protein supply). These countries also have relatively low wasting prevalence. The combination of high HIV and low wasting in this group creates a spurious negative association between HIV and wasting when all countries are included together.

Among the six staple models, maize consistently shows the highest (least negative) HIV coefficient in primary analyses and through S1–S3,S5 (β=−1·813 to −3·073). This reflects the geographic co-localisation of high HIV burden and maize consumption in southern and eastern sub-Saharan Africa: 17 of the 32 countries excluded in S4 are maize-dominant. Because these high-HIV countries are also low-wasting, their inclusion partially counteracts the spurious negative HIV-wasting association in the maize model, producing the attenuated coefficient. The pattern across S1–S3,S5 confirms this is a structural feature of the maize model rather than a chance finding.

S4 excluded countries above the Q75 HIV prevalence threshold (0·089%), removing predominantly these maize-dominant southern African countries. On their removal, the HIV coefficient reversed to positive and non-significant across all six staple models (β=+7·633 to +12·318, p=0·34–0·56), confirming the primary negative coefficient is a geographic artefact.

**S Table 12. Panel E. HIV prevalence association with child wasting in primary and five sensitivity specifications**

| **Staple** | **Primary** |  | **S1 (n=122)** |  | **S2 (n=118)** |  | **S3 (n=47)** |  | **S4 (n=95)** |  | **S5 wtd** |  |
| --- | --- | --- | --- | --- | --- | --- | --- | --- | --- | --- | --- | --- |
|  | **β** | **p** | **β** | **p** | **β** | **p** | **β** | **p** | **β** | **p** | **β** | **p** |
| Wheat | −2·880 | <0·001 *** | −2·688 | <0·001 *** | −2·588 | <0·001 *** | −4·002 | <0·001 *** | +9·244 | 0·490 | −5·256 | <0·001 *** |
| Rice | −2·340 | 0·002 ** | −2·141 | 0·003 ** | −1·996 | 0·007 ** | −4·324 | 0·003 ** | +12·318 | 0·337 | −4·456 | <0·001 *** |
| Maize | −2·093 | 0·012 * | −1·948 | 0·013 * | −1·813 | 0·024 * | −2·541 | 0·087 † | +8·385 | 0·527 | −3·073 | 0·016 * |
| Millet | −2·788 | <0·001 *** | −2·578 | <0·001 *** | −2·477 | 0·001 ** | −3·719 | 0·007 ** | +10·004 | 0·447 | −4·461 | <0·001 *** |
| Sorghum | −2·845 | <0·001 *** | −2·634 | <0·001 *** | −2·530 | <0·001 *** | −3·659 | 0·007 ** | +7·633 | 0·559 | −4·675 | <0·001 *** |
| Cassava | −2·906 | <0·001 *** | −2·678 | <0·001 *** | −2·578 | 0·001 ** | −3·813 | 0·006 ** | +11·475 | 0·406 | −3·750 | <0·001 *** |

*S4 coefficients shown in red — reversal from negative to positive confirms geographic confounding. S3 includes all African UN subregions (n=52). S4 excludes countries above Q75 HIV prevalence threshold (0·089%). Negative coefficients in primary analyses reflect geographic co-localisation of high-maize consumption with high HIV burden in Southern and Eastern Africa.*

To clarify the basis of this reversal, we compared mean child stunting and wasting prevalence across three groups of countries in our analytical sample: the 34 countries excluded under the Q75 HIV prevalence threshold in S4, split by whether maize is their dominant staple, and the 100 countries retained in S4. Of the 34 countries excluded in S4, 17 (50%) are maize-dominant, a substantially higher proportion than for any other staple. The maize-dominant excluded countries showed the highest mean stunting (29·7%) but the lowest mean wasting (4·3%) of the three groups; the non-maize excluded countries showed intermediate stunting (23·5%) but the highest mean wasting (6·9%, strongly influenced by South Sudan, an outlier at 22·7%); and the 100 S4-retained countries showed the lowest stunting (18·1%) and intermediate wasting (5·2%) (S Table 13F). This pattern — stunting rising monotonically from S4-retained to non-maize-excluded to maize-excluded countries, while wasting is lowest specifically in the maize-excluded group — is consistent with the MUAC-predominant, WHZ-attenuated signature already described for maize-dominant countries in S Section F.1 and S Table 23: a population with high true undernutrition burden (reflected in high stunting and, by the Grellety-Golden cross-reference, high MUAC-based wasting) whose WHZ-based wasting prevalence is mechanically understated by shorter average stature. The exclusion of these countries in S4 therefore removes a population in which HIV prevalence and low WHZ-detected wasting co-occur for reasons unrelated to HIV itself, which is what drives the reversal of HIV’s coefficient; it does not indicate that maize’s own protective wasting association is artefactual, since that coefficient’s direction and magnitude are materially unchanged in S4 (β=−0·130 vs −0·139 in the primary model), with the loss of statistical significance (p=0·069) attributable to the reduced and disproportionately filtered maize sub-sample (9 of 26 maize-dominant countries retained) rather than to any genuine attenuation of the association.

**S Table 12. F Mean child stunting and wasting by HIV/staple group underlying the S4 exclusion**

| **Group** | **n** | **Mean stunting %** | **Mean wasting %** |
| --- | --- | --- | --- |
| S4-excluded, maize-dominant (Q75 HIV) | 17 | 29·7 | 4·3 |
| S4-excluded, non-maize-dominant (Q75 HIV) | 17 | 23·5 | 6·9 |
| S4-retained (below Q75 HIV) | 100 | 18·1 | 5·2 |

*Stunting and wasting are country-level JME prevalence values from our primary analytical dataset; HIV grouping and maize dominance are derived from the same FAOSTAT-based classification used throughout. South Sudan (sorghum-dominant, wasting 22·7%) is an outlier within the non-maize-excluded group; excluding it lowers that group’s mean wasting to approximately 6·3%.*

**S Section D.1. Dietary staple distribution across LMICs**

The following tables show the ten LMICs with highest per capita protein supply from each dietary staple (FAOSTAT 2019–2023 five-year means). Staple protein supply maps are shown alongside each table.

Sorghum is a drought tolerant cereal crop adapted to hot, semi-arid regions with low and erratic rainfall. Sorghum has a low DIAAS, low zinc and high phytate: zinc ratio as seen from S Tables 21 & 22. The nutritional profile of a sorghum based diet is reflected in the high to very high stunting and medium to very high wasting seen in the top ten countries. All the top 10 countries are peri-Sahelian as seen in S Fig 3.

**S Table 13. Top 10 LMIC countries consuming Sorghum protein (FAOSTAT 2019–2023)**

| **#** | **Country** | **Sorghum protein (g/d)** | **Stunting (%)** | **Wasting (%)** | **Survey year** | **Animal protein (g/d)** | **kcal<2100** |
| --- | --- | --- | --- | --- | --- | --- | --- |
| 1 | Sudan^‡^ | 15·69 | 38·2 | 16·3 | 2014 | 19·7 | — |
| 2 | South Sudan^‡^ | 12·13 | 31·3 | 22·7 | 2010 | 24·3 | — |
| 3 | Burkina Faso | 11·75 | 21·1 | 10·3 | 2021 | 18·2 | — |
| 4 | Niger | 11·59 | 47·7 | 10·9 | 2022 | 10·1 | — |
| 5 | Chad^‡^ | 10·73 | 31·9 | 7·8 | 2022 | 27·8 | — |
| 6 | Mali^‡^ | 8·95 | 25·1 | 5·4 | 2024 | 10·2 | — |
| 7 | Cameroon | 6·89 | 28·9 | 4·3 | 2018 | 13·7 | — |
| 8 | Ethiopia | 5·96 | 36·8 | 6·8 | 2019 | 7·0 | — |
| 9 | Nigeria | 5·87 | 33·8 | 11·5 | 2021 | 6·9 | — |
| 10 | Togo^‡^ | 5·27 | 23·8 | 5·7 | 2017 | 8·0 | — |

*† Total kcal/d below 2100 — energy deficiency threshold.*

*‡ FAOSTAT data masked/under review; values from earlier extraction (2018–2022).*

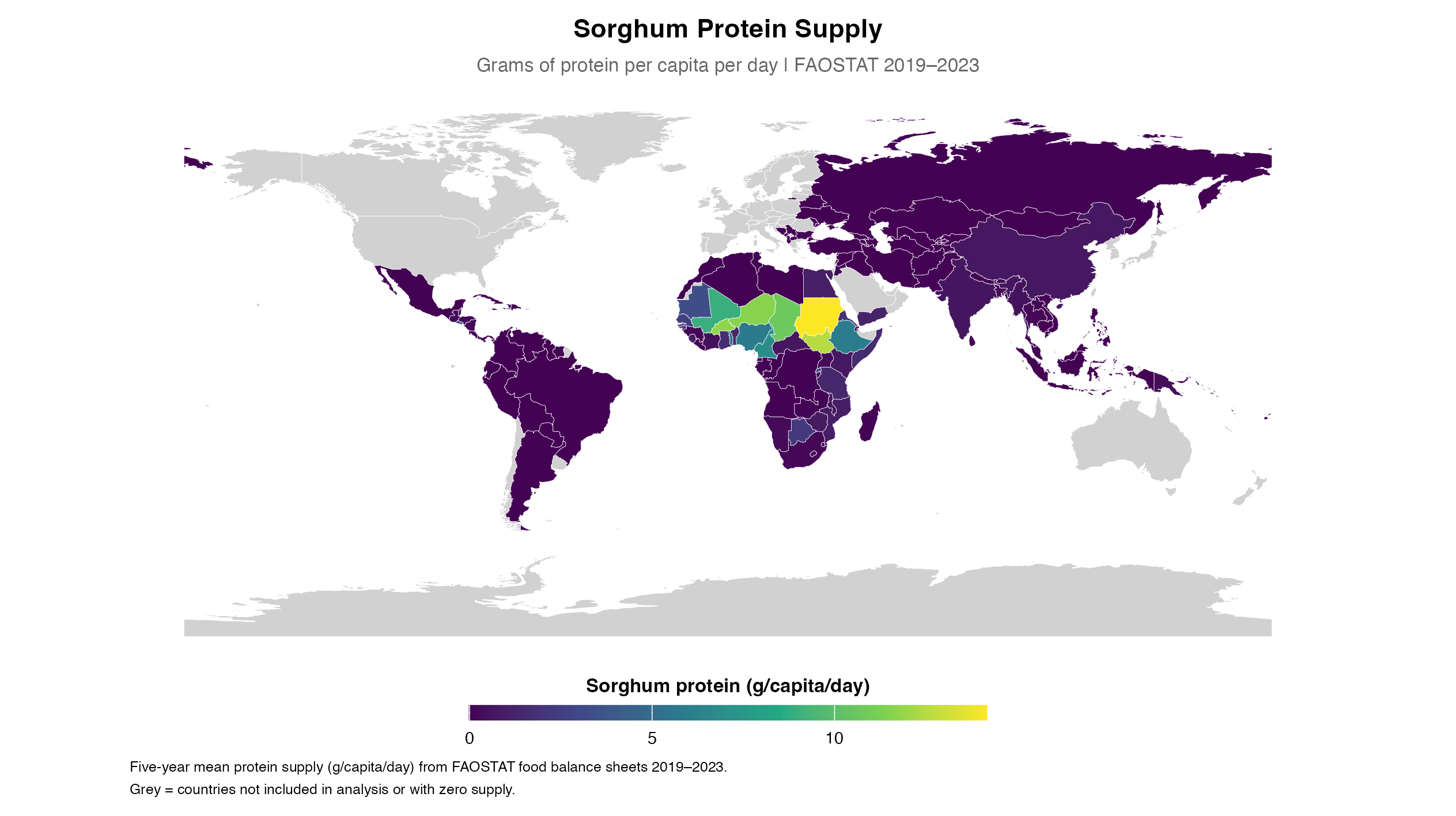

*S Figure 2. Sorghum protein supply across 127 LMICs, FAOSTAT 2019–2023 five-year mean (g/capita/day). Sudan (16·5 g/d) and South Sudan (12·1 g/d) highest consumers. Significant positive primary wasting association (β=+0·361, p<0·001). Grey=not in analysis or zero supply; Somalia excluded.*

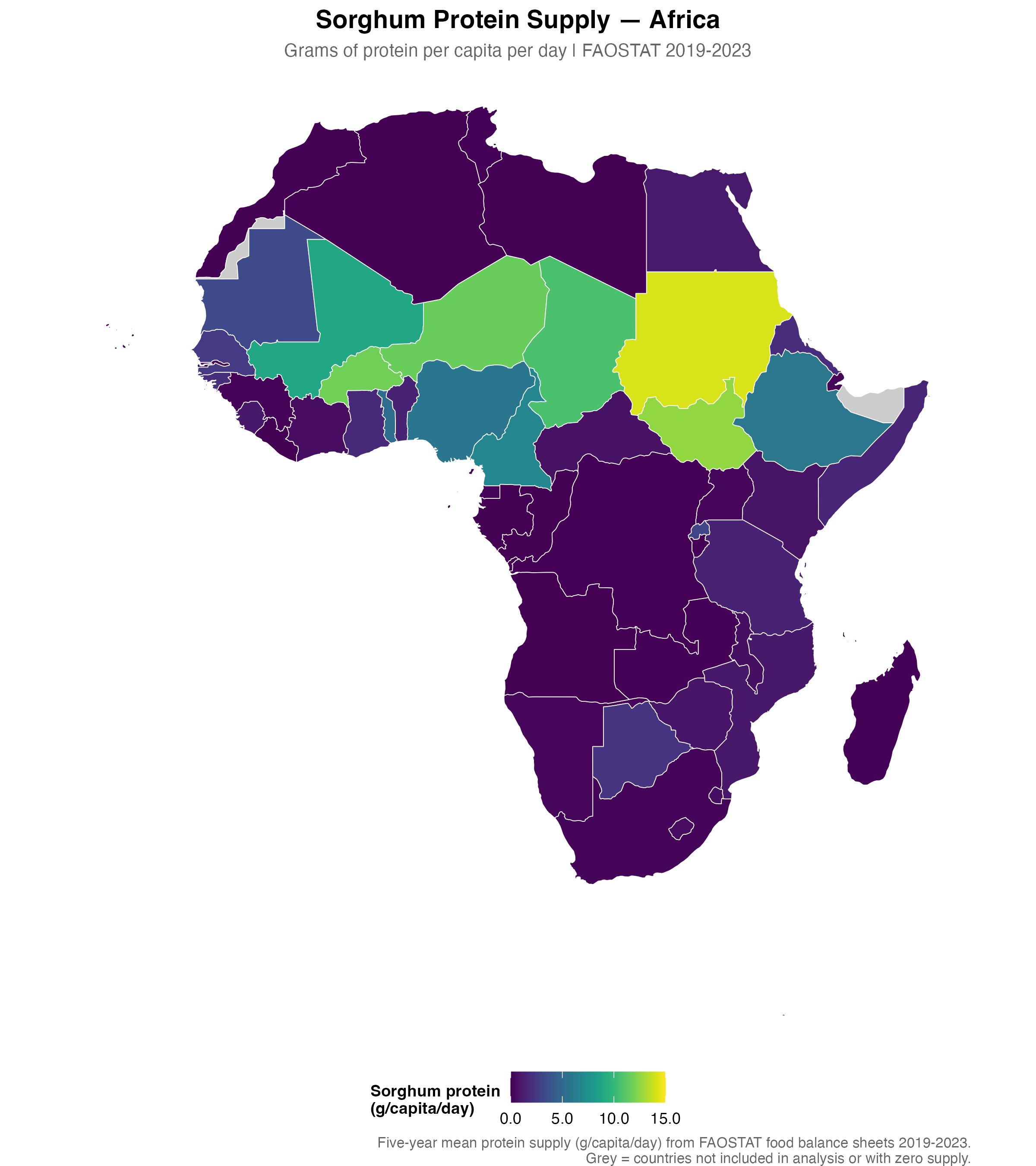

*S Figure 3. Sorghum protein supply across Africa, FAOSTAT 2019–2023 five-year mean (g/capita/day). Sahel belt dominates; Sudan highest consumer. Grey=no data.*

Millet referred to here is pearl millet( Pennisetum glaucum),since it is the most commonly consumed millet. Though as discussed earlier “millet” in FAOSTAT data is a generic term encompassing all millets. From our dataset , it is evident that the 9 of top ten countries consuming millets are from Africa. Among the top 10 countries consuming millet proteins, the highest was Niger with 18.29 gm per capita per day. The highest stunting prevalence was with 47.7 for Niger and the lowest was 17.5 for Gambia and Senegal. The highest wasting prevalence was 16.3 for Sudan and lowest 5.1 for Gambia and Guinea-Bissau. The values for both stunting and wasting range from medium to very high. Most countries are from the Sahel region of Africa as seen in the maps S Fig 4 & S Fig 5. The stunting and wasting prevalence ranges widely as per JME thresholds from medium to high and very high.

**S Table 14. Top 10 LMIC countries consuming Millet protein (FAOSTAT 2019–2023)**

| **#** | **Country** | **Millet protein (g/d)** | **Stunting (%)** | **Wasting (%)** | **Survey year** | **Animal protein (g/d)** | **kcal<2100** |
| --- | --- | --- | --- | --- | --- | --- | --- |
| 1 | Niger | 18·29 | 47·7 | 10·9 | 2022 | 10·1 | — |
| 2 | Mali^‡^ | 13·64 | 25·1 | 5·4 | 2024 | 10·2 | — |
| 3 | Chad^‡^ | 7·1 | 31·9 | 7·8 | 2022 | 27·8 | — |
| 4 | Burkina Faso | 6·43 | 21·1 | 10·3 | 2021 | 18·2 | — |
| 5 | Sudan^‡^ | 4·32 | 38·2 | 16·3 | 2014 | 19·7 | — |
| 6 | Senegal | 4·15 | 17·5 | 10·2 | 2023 | 16·1 | — |
| 7 | Gambia | 2·81 | 17·5 | 5·1 | 2020 | 19·7 | — |
| 8 | Namibia | 2·63 | 22·7 | 7·1 | 2013 | 22·5 | — |
| 9 | Nepal | 2·46 | 24·8 | 7·0 | 2022 | 17·2 | — |
| 10 | Guinea-Bissau | 2·03 | 28·1 | 5·1 | 2019 | 9·3 | — |

*† Total kcal/d below 2100 — energy deficiency threshold.*

*‡ FAOSTAT data masked/under review; values from earlier extraction (2018–2022).*

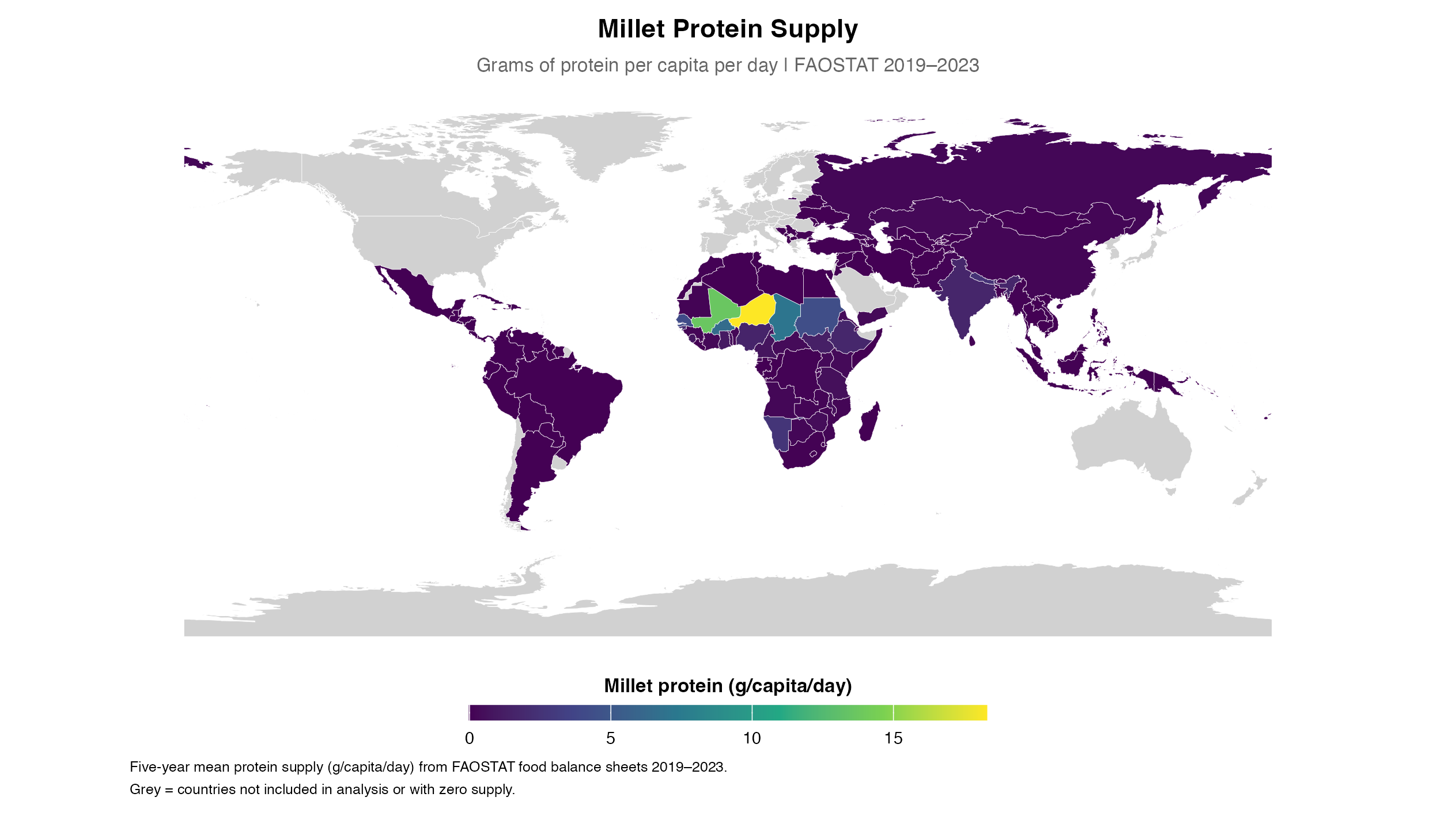

*S Figure 4. Millet protein supply across 127 LMICs, FAOSTAT 2019–2023 five-year mean (g/capita/day). Niger (18·3 g/d) and Mali (13·6 g/d) highest consumers. FAOSTAT millet is a single undifferentiated aggregate. Grey=not in analysis or zero supply; Somalia excluded.*

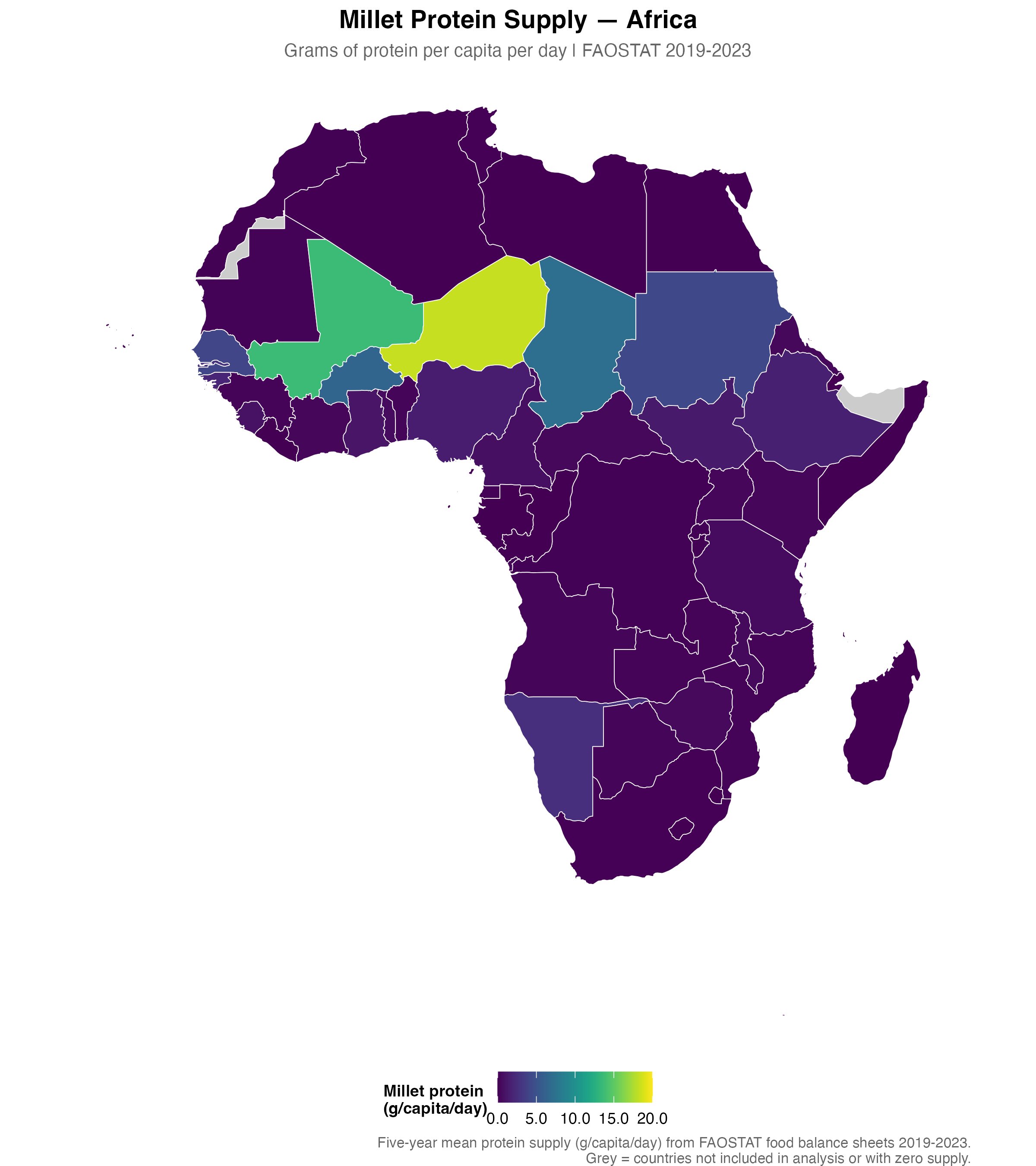

*S Figure 5. Millet protein supply across Africa, FAOSTAT 2019–2023 five-year mean (g/capita/day). Consumption declines sharply south of the Sahel. Grey=no data.*

The location of top countries consuming cassava is South of the Sahel in Africa. Four of top 10 Cassava consuming countries are from Central Sub-Saharan Africa. All other 6 among top 10 countries are from adjoining Eastern and Western Sub-Saharan Africa. The highest consumption of cassava protein was 10.72 of DRC, which was considerably lower than top consumers of other staples because of the very low protein content of cassava. The highest stunting prevalence was seen in Burundi with 52.8%(which was the highest among all countries as well). Of note, is that among these countries the lowest animal protein consumption was for Burundi at 3.1 gm per capita per day and DRC with 3.6 gm per capita per day. The medium to very high stunting and low to medium levels of wasting are reflected in countries in the top 10 countries with cassava protein availability. Though, from the previous secondary cross-reference analysis, a large proportion of MUAC based wasting will be missed in these countries if only weight for height is taken into consideration.

**S Table 15. Top 10 LMIC countries with Cassava protein availability (FAOSTAT 2019–2023)**

| **#** | **Country** | **Cassava protein (g/d)** | **Stunting (%)** | **Wasting (%)** | **Survey year** | **Animal protein (g/d)** | **kcal<2100** |
| --- | --- | --- | --- | --- | --- | --- | --- |
| 1 | Democratic Republic Of The Congo | 10·72 | 44·7 | 7·2 | 2023 | 3·6 | † |
| 2 | Ghana | 6·21 | 17·4 | 5·8 | 2022 | 17·8 | — |
| 3 | Equatorial Guinea^§^ | 4·85 | 26·2 | 3·1 | 2011 | 22·5 | — |
| 4 | Congo | 4·81 | 21·2 | 8·2 | 2014 | 29·5 | — |
| 5 | Burundi^‡^ | 4·81 | 52·8 | 8·0 | 2024 | 3·1 | † |
| 6 | Angola | 4·68 | 37·6 | 4·9 | 2015 | 14·9 | — |
| 7 | Côte D’Ivoire | 4·47 | 23·4 | 8·1 | 2021 | 16·1 | — |
| 8 | Mozambique | 4·44 | 36·7 | 3·8 | 2022 | 9·0 | — |
| 9 | Guinea | 3·67 | 26·1 | 6·4 | 2022 | 13·1 | — |
| 10 | Zambia | 3·62 | 34·6 | 4·2 | 2018 | 13·2 | — |

*† Total kcal/d below 2100 — energy deficiency threshold.*

*‡ FAOSTAT data masked/under review; values from earlier extraction (2018–2022).*

*§ FAOSTAT data never compiled; values imputed from GBD super-region mean.*

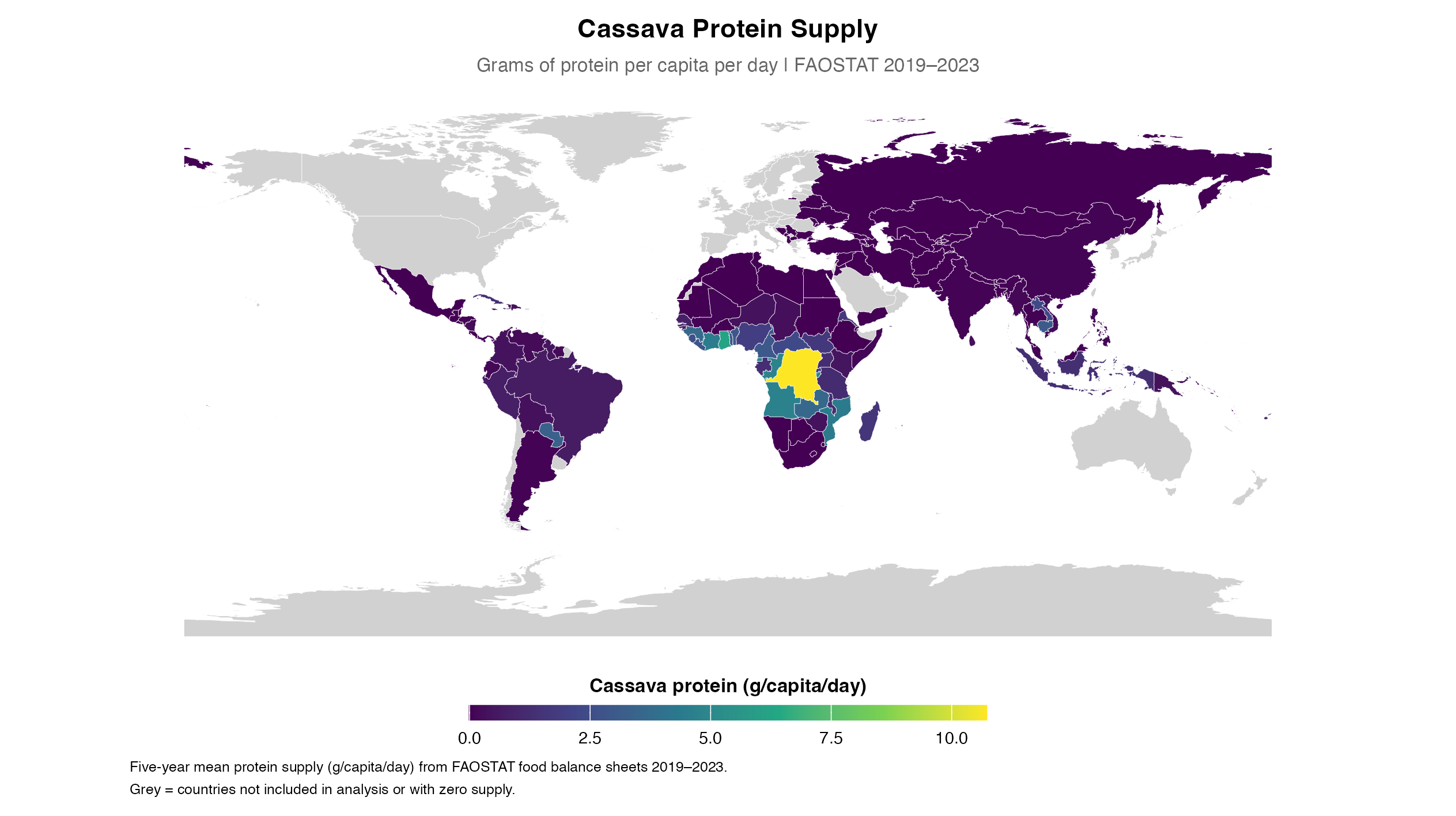

*S Figure 6. Cassava protein supply across 127 LMICs, FAOSTAT 2019–2023 five-year mean (g/capita/day). DRC (10·7 g/d) highest consumer. Lowest protein density of the six staples. Grey=not in analysis or zero supply; Somalia excluded.*

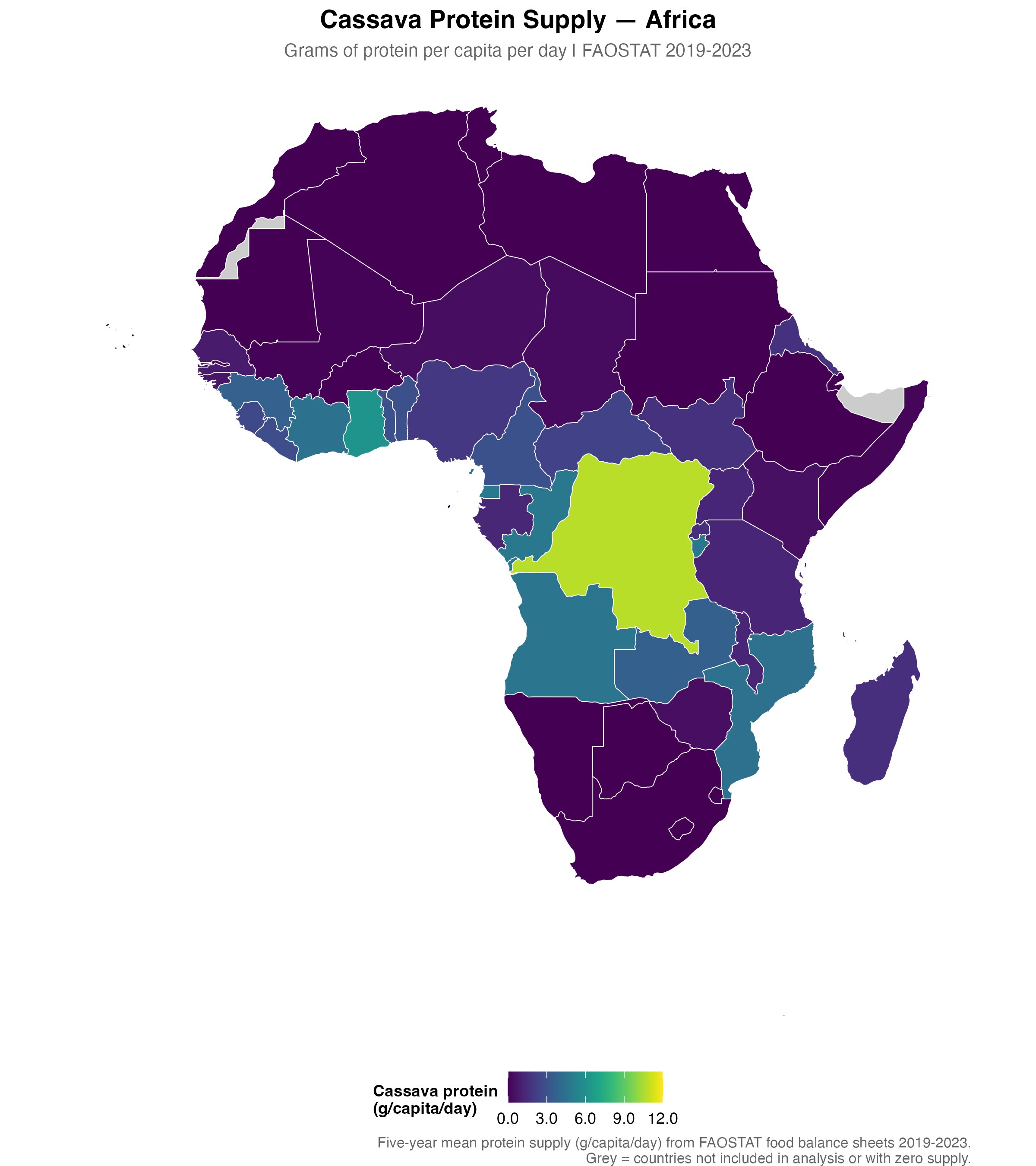

*S Figure 7. Cassava protein supply across Africa, FAOSTAT 2019–2023 five-year mean (g/capita/day). DRC dominates; cassava belt spans Central and West Africa. Grey=no data.*

Five of the top 10 maize consuming countries were located in Sub-Saharan Africa (Malawi, Lesotho, South Africa, Zambia, Eswatini) and five in Latin America (Mexico, Guatemala, Paraguay, El Salvador, Honduras). The highest stunting prevalence among them was Guatemala with a 46% prevalence and lowest was Paraguay at 5.6%.Wasting was consistently low ranging from Zambia with 4.2 which was the highest to Guatemala with 0.8, the lowest. This is consistent the low zinc and high phytate: zinc ratio of maize. The lower height could be contributing to lower wasting by virtue of the denominator being height in WHZ calculations of wasting. However, like cassava from our secondary cross-reference analysis in S Section F, it could be that wasting by MUAC may be missed in the above data. The contrast between the strong negative maize–wasting association (β=−0·139, p=0·004) and the weak positive maize–stunting association (β=+0·124, p=0·333) in the primary analysis reflects the dual nutritional profile of maize: moderate protein quality (DIAAS ~40) combined with a high phytate:zinc molar ratio (45·5). Maize-dominant diets appear to drive zinc deficiency–mediated linear growth failure (stunting) while the resulting short stature attenuates WHZ-based wasting estimates through the height denominator effect — the height denominator effect, consistent with the contrasting phenotypes observed by Garenne et al (2009) across sorghum/millet- and cassava/maize-consuming populations and further supported by the Grellety & Golden cross-reference, where maize-dominant countries cluster in the MUAC-predominant wasting zone. The heterogeneity of the top 10 maize consumers — spanning low-SDI SSA countries (Malawi, Zambia, Lesotho) and higher-SDI Latin American countries (Mexico SDI=0·69, Paraguay SDI=0·67) — explains why the maize–stunting ecological signal does not reach statistical significance after covariate adjustment, despite the biological plausibility being well supported by individual country profiles.

**S Table 16. Top 10 LMIC countries consuming Maize protein (FAOSTAT 2019–2023)**

| **#** | **Country** | **Maize protein (g/d)** | **Stunting (%)** | **Wasting (%)** | **Survey year** | **Animal protein (g/d)** | **kcal<2100** |
| --- | --- | --- | --- | --- | --- | --- | --- |
| 1 | Malawi | 24·59 | 35·5 | 2·6 | 2020 | 16·0 | — |
| 2 | Mexico | 20·88 | 12·5 | 1·0 | 2022 | 55·5 | — |
| 3 | Lesotho | 17·9 | 35·6 | 1·6 | 2024 | 16·2 | † |
| 4 | South Africa | 17·45 | 21·3 | 3·7 | 2017 | 38·1 | — |
| 5 | Zambia | 17·44 | 34·6 | 4·2 | 2018 | 13·2 | — |
| 6 | Guatemala | 16·36 | 46·0 | 0·8 | 2021 | 30·2 | — |
| 7 | Paraguay | 15·84 | 5·6 | 1·0 | 2016 | 30·5 | — |
| 8 | El Salvador | 15·83 | 10·0 | 2·9 | 2021 | 35·1 | — |
| 9 | Eswatini | 15·59 | 20·0 | 1·8 | 2021 | 17·8 | — |
| 10 | Honduras | 15·33 | 18·7 | 1·9 | 2019 | 25·1 | — |

*† Total kcal/d below 2100 — energy deficiency threshold.*

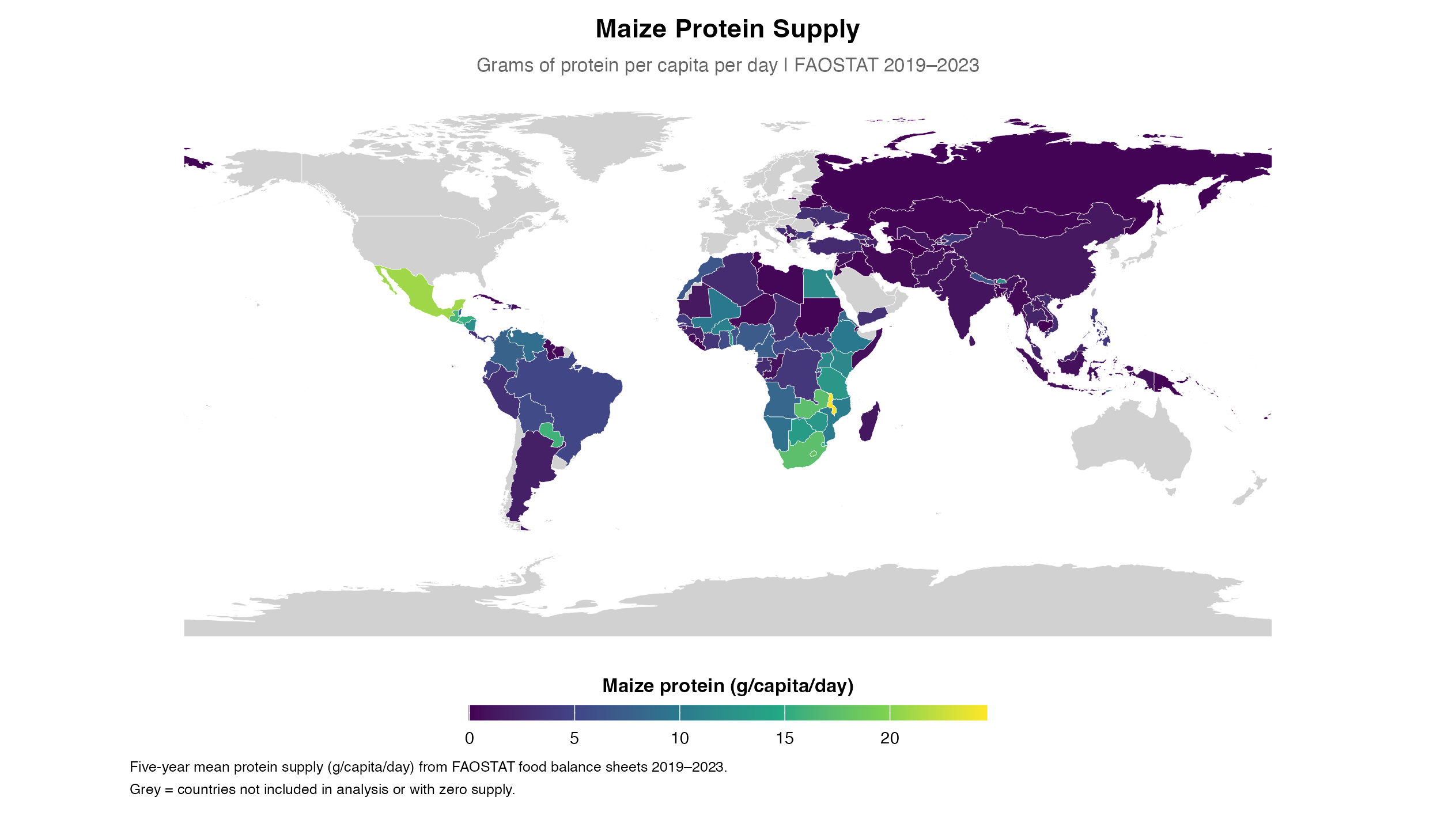

*S Figure 8. Maize protein supply across 127 LMICs, FAOSTAT 2019–2023 five-year mean (g/capita/day). Malawi (24·6 g/d) and Mexico (20·9 g/d) highest consumers. Significant negative primary wasting association (β=−0·139, p=0·004). Grey=not in analysis or zero supply; Somalia excluded.*

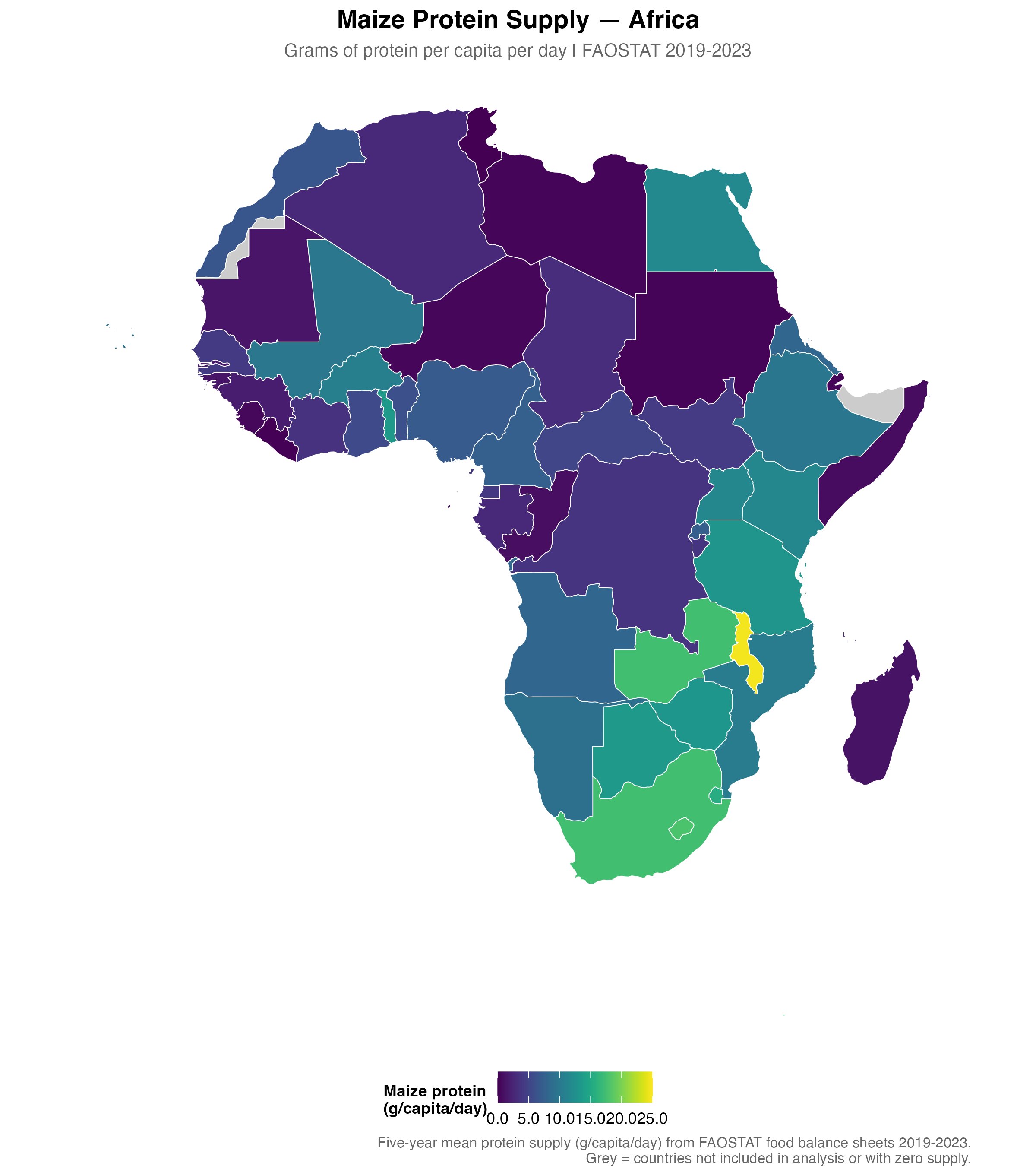

*S Figure 9. Maize protein supply across Africa, FAOSTAT 2019–2023 five-year mean (g/capita/day). Southern and East Africa dominate; Malawi highest. Grey=no data.*

All the top 10 rice consuming countries were from South Asia and South East Asia as seen in S Table 17 and S Fig 10, with the exception of Madagascar and Guinea-Bissau which were from Sub-Saharan Africa. The highest stunting prevalence among them was Madagascar at 39·8% and the lowest was Sri Lanka at 10·5%. Wasting prevalence ranged from a high of 10·7% for Bangladesh to a low of 4·4% for Vietnam. As per JME thresholds, wasting values for countries ranged from medium to high, while stunting also ranged from medium to high. The negative association of rice with stunting observed in the sensitivity analyses (S3 and S5) is consistent with the mechanistic role of animal protein in rice-dominant countries. In the primary rice model, animal protein showed a statistically significant negative association with stunting (β=−0·112, p=0·044*), whereas the rice protein coefficient itself was non-significant (β=−0·075, p=0·379). Rice-dominant countries in South and South East Asia — particularly Bangladesh, Vietnam, Cambodia and Myanmar — typically complement rice with small whole fish and other aquatic protein sources, providing complete protein, bioavailable zinc and vitamin A. It is this animal protein complementarity, not rice protein per se, that appears to protect against stunting in these settings. Rice maintains stature better than sorghum or millet (via its lower phytate:zinc ratio), and animal protein complementation further reduces stunting — together explaining the WHZ-predominant wasting pattern seen in rice-dominant countries: stature is relatively preserved, so lean mass depletion is detectable by WHZ. The negative association of rice with stunting observed in the quantile regression (S3 and S5) and sensitivity analyses may reflect the fact that rice protein, despite its moderate DIAAS, is better suited for dietary complementarity with other protein sources in the South and South East Asian context.

**S Table 17. Top 10 LMIC countries consuming Rice protein (FAOSTAT 2019–2023)**

| **#** | **Country** | **Rice protein (g/d)** | **Stunting (%)** | **Wasting (%)** | **Survey year** | **Animal protein (g/d)** | **kcal<2100** |
| --- | --- | --- | --- | --- | --- | --- | --- |
| 1 | Bangladesh | 32·98 | 23·6 | 10·7 | 2022 | 16·1 | — |
| 2 | Myanmar | 31·19 | 26·7 | 6·7 | 2018 | 27·6 | — |
| 3 | Lao People'S Democratic Republic | 28·72 | 32·8 | 10·7 | 2023 | 24·1 | — |
| 4 | Cambodia | 28·67 | 21·9 | 9·6 | 2021 | 20·3 | — |
| 5 | Viet Nam | 27·89 | 18·2 | 4·4 | 2023 | 37·7 | — |
| 6 | Sri Lanka | 25·46 | 10·5 | 9·3 | 2024 | 18·9 | — |
| 7 | Philippines | 24·76 | 26·7 | 5·4 | 2021 | 28·3 | — |
| 8 | Bhutan | 24·02 | 17·9 | 5·1 | 2023 | 17·3 | — |
| 9 | Madagascar | 23·92 | 39·8 | 7·2 | 2021 | 5·1 | † |
| 10 | Guinea-Bissau | 23·19 | 28·1 | 5·1 | 2019 | 9·3 | — |

*† Total kcal/d below 2100 — energy deficiency threshold.*

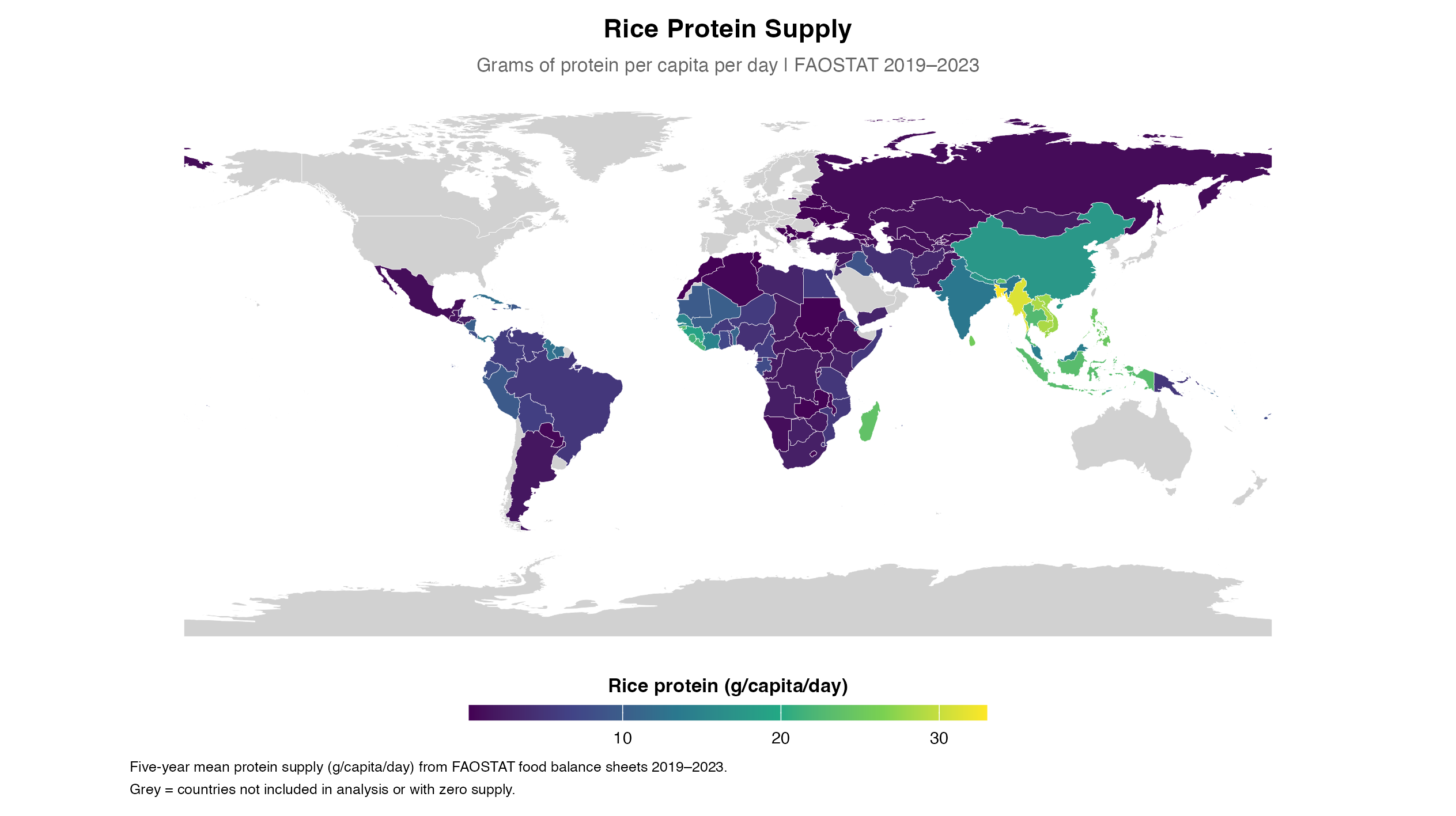

*S Figure 10. Rice protein supply across 127 LMICs, FAOSTAT 2019–2023 five-year mean (g/capita/day). Bangladesh (33·0 g/d) and Myanmar (31·2 g/d) highest consumers. Significant positive primary wasting association (β=+0·086, p=0·004). Grey=not in analysis or zero supply; Somalia excluded.*

All the top 10 wheat consuming countries were from North Africa and the Middle East as well as Central Asia, with the exception of Serbia which was from Central Europe, as seen in S Table 18 and S Fig 11. They all had low to very low wasting prevalence and low to medium stunting prevalence.

**S Table 18. Top 10 LMIC countries consuming Wheat protein (FAOSTAT 2019–2023)**

| **#** | **Country** | **Wheat protein (g/d)** | **Stunting (%)** | **Wasting (%)** | **Survey year** | **Animal protein (g/d)** | **kcal<2100** |
| --- | --- | --- | --- | --- | --- | --- | --- |
| 1 | Serbia | 48·92 | 5·4 | 2·6 | 2019 | 61·8 | — |
| 2 | Tunisia | 47·25 | 8·4 | 2·1 | 2018 | 32·9 | — |
| 3 | Azerbaijan | 43·23 | 6·6 | 3·5 | 2023 | 37·5 | — |
| 4 | Morocco | 42·85 | 14·2 | 2·4 | 2019 | 29·7 | — |
| 5 | Algeria | 42·73 | 9·8 | 2·7 | 2019 | 27·0 | — |
| 6 | Afghanistan | 40·48 | 44·6 | 3·6 | 2022 | 9·8 | — |
| 7 | Turkmenistan | 37·87 | 7·2 | 4·1 | 2019 | 53·9 | — |
| 8 | Uzbekistan | 37·61 | 6·5 | 2·4 | 2021 | 49·4 | — |
| 9 | Türkiye | 37·6 | 6·0 | 1·7 | 2018 | 45·0 | — |
| 10 | Iran (Islamic Republic Of) | 37·52 | 4·8 | 4·3 | 2017 | 25·9 | — |

*† Total kcal/d below 2100 — energy deficiency threshold.*

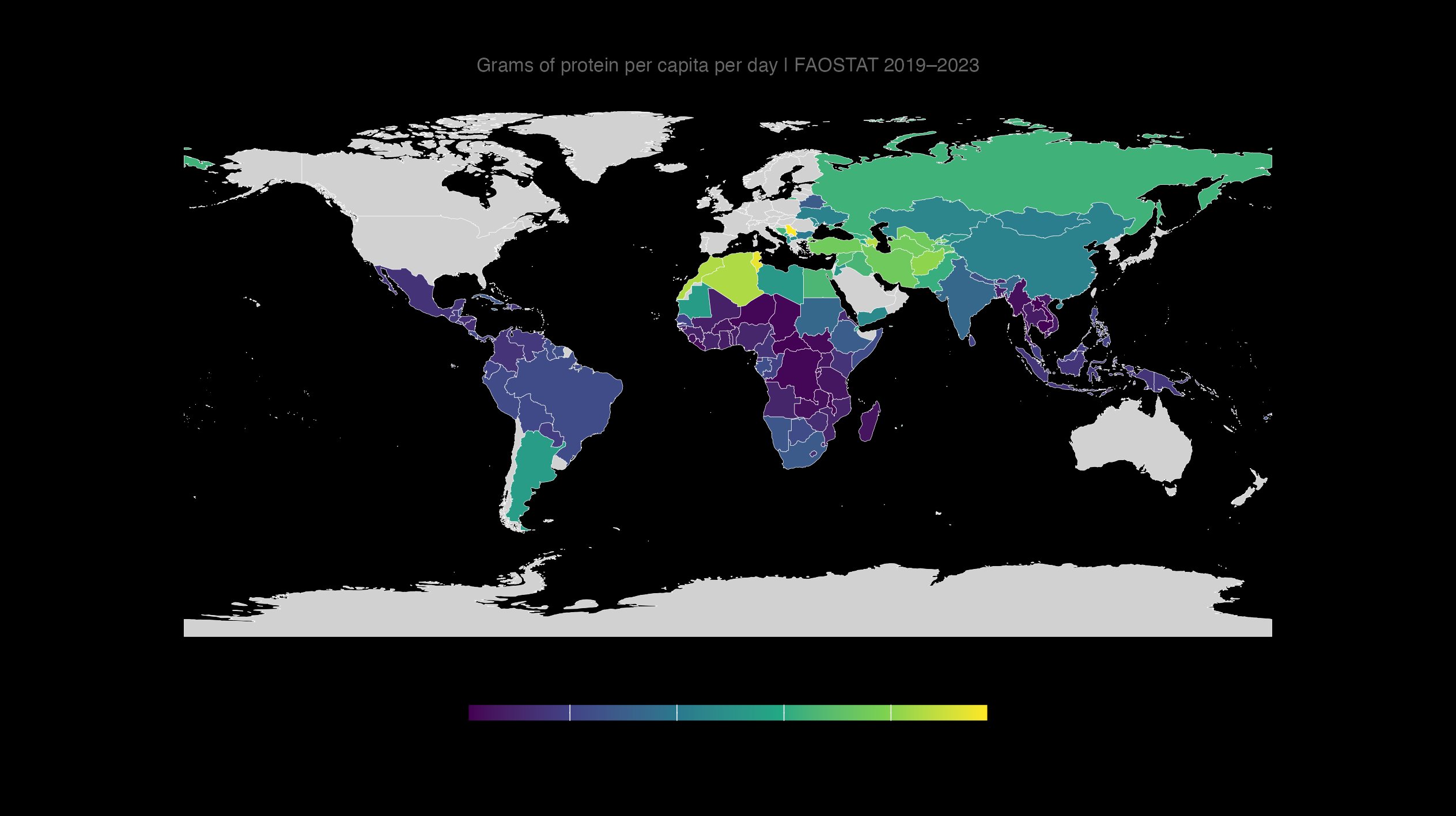

*S Figure 11. Wheat protein supply across 127 LMICs, FAOSTAT 2019–2023 five-year mean (g/capita/day). Serbia (48·9 g/d) and Tunisia (47·3 g/d) highest consumers. No significant primary wasting association. Grey=not in analysis or zero supply; Somalia excluded.*

**S Section E. Protein quality and zinc bioavailability of dietary staples**

A systematic search of PubMed was conducted using the terms: DIAAS, protein quality, protein digestibility, and each staple in turn (rice, wheat, maize, sorghum, millet, cassava). Most values are from animal models (pig, rat); human IAAO-based estimates are available only for pearl millet and sorghum. Values are relevant for age group specified. The DIAAS values are provided in S Table 19. Zinc content and phytate:zinc molar ratios were extracted from the Composite Nutrient Database reported in the supporting information of Wessells et al (Wessells KR, Singh GM, Brown KH. Estimating the Global Prevalence of Inadequate Zinc Intake from National Food Supply Data. J Nutr. 2012;142(7):1333–1343; Table S1). The zinc values and phytate:zinc ratios of discussed staples are provided in S Table 20.

**S Table 19. DIAAS reference values for staple cereals and tubers**

| **Staple** | **Protein (g/100g)** | **DIAAS (limiting AA, age)** | **Method** | **Caveat** | **Reference** |
| --- | --- | --- | --- | --- | --- |
| **Wheat** | 12·2 | **48 (Lys, 6m–3y)** | Pig model | *Average across published studies; cooked or raw not specified.* | 20 |
| **Rice** | 6·7 | **47 (Lys, 6m–3y)** | Pig model | *Average across published studies; cooked or raw not specified.* | 20 |
| **Maize** | 9·5 | **36 (Lys, 6m–3y)** | Pig model | *Average across published studies; cooked or raw not specified.* | 20 |
| **Finger millet** | — | **40 (Lys, 6m–3y)** | Human — dual isotope tracer | *Cooked millet pancake; Indian children 1–2 years. Method validated in children.* | 21 |
| **Pearl millet** | 9·7 | **68 (Lys, adults)** | Human — IAAO method | *Moist cooking method; healthy young adult males only. Not validated for children <3 years. Disputed — see footnote.* | 22 |
| **Sorghum (pig model)** | 10·1 | **29 (Lys, adults)** | Pig model | *Raw sorghum; applicable to older children/adults.* | 23 |
| **Sorghum (IAAO)** | 10·1 | **~59 (Lys, adults)** | Human — IAAO method | *Moist cooking method; young adult males. Higher than pig value — moist cooking effect. Not validated for children.* | 24 |
| **Cassava (De Vries)** | 0·9 | **17 (Leu, 6m–3y) 1st limiting** | In vitro/estimated | *Very low protein density (0·9 g/100g). Leucine is first limiting amino acid.* | 26 |
| **Cassava (Parikh)** | 2·1 | **~37 (Leu, 6m–3y) 1st limiting** | In vitro estimate | *Dry weight protein; Leucine 1st limiting, Isoleucine 2nd. No validated in vivo children's study available.* | 25 |
| **Brown rice** | — | **42 (Lys, 6m–3y)** | Rat model | Cooked | 21 |
| **Proso millet** | — | **7 (Lys, 6m–3y)** | Rat model | Cooked | 21 |
| **Foxtail millet** | — | **10 (Lys, 6m–3y)** | Rat model | Cooked | 21 |

*DIAAS: Digestible Indispensable Amino Acid Score. Lys=lysine; Leu=leucine. Age: 6m–3y = 6 months to 3 years; adults = >3 years (FAO 2013). Brown rice, proso millet and foxtail millet values from Han et al (rat model, cooked). Sorghum DIAAS estimated from true ileal amino acid digestibility data in Millward DJ, Jackson AA. Protein requirements and the indicator amino acid oxidation method. Am J Clin Nutr. 2012;95(6):1498–1501.*^23^ *These minor millet species are not individually reported in FAOSTAT food balance sheets.Reference number pertains to the no in bibliography.*

FAOSTAT reports millet as a single undifferentiated aggregate (Item Code 2517, "Millet and products"; Item Code 79 in FAOSTAT definitions and standards, CPC 0118) without species disaggregation — species composition is not documented in FAOSTAT definitions and standards. Millet encompasses nutritionally distinct species including pearl millet, finger millet, teff and other minor millets, which vary substantially in protein quality and geographic distribution; this limits interpretation of the FAOSTAT millet-wasting association. Pearl millet DIAAS (Fakiha et al 2020)^24^ is based on the IAAO method by moist cooking in healthy young adult males only — not validated for children under 3 years — and is disputed on methodological grounds (Millward & Jackson 2012).^25^ Sorghum value is higher by the IAAO and for young adults by moist cooking method than values in the pig model.^26^

Cassava DIAAS values are estimates from amino acid composition data — no validated in vivo children's study available.^27,28^

**S Table 20. Zinc content and phytate: zinc molar ratios for six staples**

| **Staple** | **Energy (kcal/100g)** | **Zinc (mg/100g)** | **Phytate (mg/100g)** | **Phytate:Zinc molar ratio** | **Notes** |
| --- | --- | --- | --- | --- | --- |
| **Wheat (whole grain flour)** | 340 | 2·6 | 777 | 29·6 | *High phytate:zinc ratio. Whole grain has higher zinc but also higher phytate than refined flour — FAOSTAT does not distinguish processing.* |
| **Maize (whole grain, white)** | 362 | 1·8 | 835 | **45·5** | *Second highest phytate:zinc ratio. Nixtamalisation substantially reduces phytate in Central America/Mexico but not in Southern/Eastern Africa (ugali, nshima).* |
| **Millet (pearl millet)** | 383 | 2·5 | 682 | 27·0 | *Similar phytate:zinc ratio to wheat. Fermentation and soaking reduce phytate — common in Sahelian preparation methods.* |
| **Sorghum** | 375 | 1·8 | 900 | **48·9** | *Highest phytate:zinc ratio of all six staples. Combined with lowest DIAAS (~29), sorghum presents the most adverse combined protein-zinc profile.* |
| **Cassava** | 160 | 0·3 | 39 | 16·9 | *Very low zinc content and low phytate:zinc ratio. Primary nutritional concerns are extremely low protein content and cyanogenic glycosides — not zinc.* |
| **Rice (white, raw)** | 361 | 1·1 | 226 | 20·1 | *Lower phytate:zinc ratio than cereals. Polishing reduces both zinc and phytate; refined rice has less zinc than brown rice but also less phytate inhibition.* |

Source: Composite Nutrient Database from Wessells KR, Singh GM, Brown KH. Estimating the Global Prevalence of Inadequate Zinc Intake from National Food Supply Data. J Nutr. 2012;142(7):1333–1343 (Table S1). Phytate:zinc molar ratio >15 considered inhibitory to zinc absorption; >25 associated with zinc deficiency risk (Wessells et al. 2012).^29^

*Red shading: phytate:zinc molar ratio >40 — sorghum (48·9) and maize (45·5).*

**S Section F. Secondary cross-reference: WHZ-MUAC discordance by dietary staple**

Our primary analysis uses wasting defined by weight-for-height Z-score (WHZ −2 SD), consistent with JME March 2025 methodology. In the course of our literature search, our findings on sorghum and millet associations with child undernutrition led us to the seminal work of Garenne et al (2009), who documented starkly contrasting patterns of child undernutrition — including stunting, WHZ-based wasting, MUAC-based wasting and oedema — across two populations in sub-Saharan Africa: one consuming millet and sorghum in Niakhar, Senegal, and one consuming maize and cassava in Bwamanda, Democratic Republic of Congo.

The divergent anthropometric phenotypes observed across these two staple-diet populations suggested that the dominant dietary staple may shape not only undernutrition prevalence but its phenotypic expression across different anthropometric indices. This led us to cross-reference the Grellety-Golden dataset (Grellety E, Golden MH. BMC Nutrition 2016;2:10, Table S2), which documents the proportion of wasting detected by WHZ only, MUAC only, and both criteria simultaneously across 47 nationally representative surveys, to examine whether staple-type consumption patterns were associated with WHZ–MUAC discordance at the population level. Applying a dietary staple lens to this dataset, we observed that maize- and cassava-dominant countries tended toward MUAC-predominant wasting, while sorghum- and millet-dominant countries tended toward WHZ-predominant wasting.

Critically, maize- and cassava-dominant countries also carry substantially higher stunting burdens — a pattern consistent with the Garenne et al (2009) findings comparing the Bwamanda (DRC) and Niakhar (Senegal) cohorts, where the population consuming maize and cassava showed higher stunting and MUAC-predominant wasting while the millet/sorghum-consuming population showed WHZ-predominant wasting. This is also consistent with the negative association between stunting prevalence and WHZ–MUAC discordance observed in our Spearman analysis (r=−0·463, p=0·001). This stunting-mediated mechanism warrants explicit explanation: in populations with high stunting prevalence, children have shorter stature for their age. Because WHZ is computed relative to the expected weight for a child’s measured height, a stunted child who has also lost lean mass may have a less severely depressed WHZ than a non-stunted child with equivalent lean mass depletion — because their lower height denominator partially compensates the weight deficit. MUAC, which directly measures mid-upper arm circumference independently of height, is therefore the more sensitive indicator of wasting in high-stunting maize- and cassava-dominant populations.

In maize- and cassava-dominant countries where stunting is prevalent, this artefactual attenuation of WHZ means that JME’s WHZ-based wasting estimates systematically undercount the true wasting burden. The reverse holds in sorghum- and millet-dominant countries: lower stunting prevalence means children maintain greater stature for their age. When lean mass is lost in a taller child, the weight deficit relative to their measured height is proportionally larger, producing a more pronounced WHZ deficit. This bidirectional mechanism parsimoniously explains why WHZ-based JME estimates may overestimate the true relative burden of wasting in sorghum- and millet-dominant countries compared with maize- and cassava-dominant ones, where the converse applies.

Applying this framework to the Grellety & Golden dataset yields a consistent pattern. Among countries with FAOSTAT 2019–2023 staple supply data, the dominant wasting indicator pattern by staple group is as follows. Sorghum-dominant countries show consistently WHZ-predominant wasting — more children detected by WHZ alone than by MUAC alone (Grellety: mean WHZ−MUAC discordance +39·7 percentage points of wasted children). Millet-dominant countries show the same direction at lower magnitude (Grellety: +22·5). Rice-dominant countries are predominantly WHZ-predominant (Grellety: +28·4, 80% of countries), consistent with rice-consuming populations generally maintaining better stature. Wheat-dominant countries show WHZ-predominant wasting in the majority of cases. By contrast, maize-dominant countries show a mixed or MUAC-predominant pattern: in the Grellety dataset, 73% of maize-dominant countries show MUAC-predominant wasting, reflecting the heterogeneity of maize-consuming populations across Sub-Saharan Africa and Latin America. Cassava-dominant countries consistently show MUAC-predominant or near-equal wasting, consistent with the high stunting burden and energy deficit characteristic of cassava-dominated diets.

These patterns are broadly consistent with the hypothesis that staple protein quality and zinc bioavailability shape not only wasting prevalence but its phenotypic expression across anthropometric indices. Sorghum and millet — with the lowest DIAAS — appear to produce a wasting phenotype detectable preferentially by WHZ, while maize and cassava — associated with higher stunting — produce a wasting phenotype where MUAC is the more sensitive indicator through the stunting-mediated height attenuation mechanism described above. Rice and wheat fall between these extremes, with WHZ predominating in most settings.

A similar staple-stratified pattern was observed in an independent analysis of 882 population-representative anthropometric surveys from 41 countries conducted in humanitarian settings (Leidman et al, BMC Nutrition 2019;5:39). In such settings, acute food deprivation would be expected to depress WHZ more severely than MUAC, biasing toward WHZ-predominant wasting detection and potentially obscuring any MUAC-predominant signal. The persistence of the staple-stratified pattern despite this directional bias lends additional support to the hypothesis that dietary staple type shapes anthropometric wasting phenotype independently of acute food security status.

This analysis is theory-driven, exploratory and hypothesis-generating. The sample is limited to 47 surveys and the staple classification is ecological. It is presented as a cross-reference to the primary findings rather than confirmatory evidence.

Spearman rank correlations between WHZ−MUAC difference and key variables across the 46 surveys are presented in S Table 21 Panel C. Higher sorghum, millet and rice protein supply were each positively associated with WHZ predominance (sorghum r=+0·295, p=0·047; millet r=+0·383, p=0·009; rice r=+0·443, p=0·002), consistent with the primary regression findings. Maize showed a negative correlation (r=−0·174, p=0·247) — not significant but in the expected direction of MUAC predominance. Wheat was non-significant (r=+0·238, p=0·112) with a positive trend. The negative association between stunting prevalence and WHZ−MUAC difference (r=−0·463, p=0·001) reflects the known anthropometric relationship — in high-stunting populations, the height denominator of WHZ is reduced, attenuating the WHZ deficit and causing MUAC to predominate as the more sensitive wasting indicator. Spearman r: rank correlation coefficient ranging from −1 (perfect inverse relationship) to +1 (perfect positive relationship); p-value indicates probability of observing this association by chance if no true relationship exists.

**S Section F.1. WHZ-MUAC discordance by dietary staple (Grellety-Golden cross-reference, n=47)**

*Data arrangement — Grellety-Golden cross-reference: Country-level WHZ-only, MUAC-only and concurrent WHZ+MUAC wasting percentages were extracted from Grellety and Golden (BMC Nutr 2016;2:10, Supplementary Table S2), which reported these values for 47 nationally representative DHS and MICS surveys. Stunting prevalence used in Spearman correlations was taken from the same Grellety dataset (their survey-derived stunting column), ensuring that the WHZ−MUAC discordance and stunting values derive from the same surveys and are temporally consistent. These Grellety stunting values are not from our JME March 2025 master dataset. Each of the 47 countries was then linked to FAOSTAT 2019–2023 five-year mean staple protein supply (g/capita/day) from our primary analysis dataset. Dominant staple was defined as the staple with the highest per capita protein supply. Second staple was the staple with the second highest value. WHZ−MUAC difference was calculated as WHZ-only% minus MUAC-only%, where positive values indicate WHZ predominance and negative values indicate MUAC predominance. Spearman rank correlations were computed between this difference and each staple protein supply variable and stunting prevalence using R version 4·5·3 (cor.test, method=‘spearman’).*

**S Table 21. Panel A.Secondary cross-reference: WHZ-MUAC discordance by dietary staple (Grellety-Golden, n=47)**

WHZ/MUAC data from Grellety and Golden (BMC Nutr 2016;2:10, Table S2). Dominant and second staple by FAOSTAT 2019–2023 five-year mean protein supply. Theory-driven secondary cross-reference analysis.

| **Country** | **Dominant staple** | **2nd staple** | **WHZ only %** | **MUAC only %** | **WHZ−MUAC diff** | **Stunt %** | **Wast WHZ%** | **Pattern** |
| --- | --- | --- | --- | --- | --- | --- | --- | --- |
| **Guatemala** | **Maize** | Wheat | 18.6 | 58.8 | **-40.2** | 46.0 | 0.8 | *MUAC strongly predominates* |
| **Malawi** | **Maize** | Wheat | 21.5 | 52.8 | **-31.3** | 34.9 | 2.6 | *MUAC strongly predominates* |
| **Tajikistan** | **Wheat** | Rice | 19.1 | 49.9 | **-30.8** | 13.7 | 6.4 | *MUAC strongly predominates* |
| Burundi | **Cassava** | Maize | 23.3 | 43.1 | -19.9 | 52.8 | 7.8 | *MUAC predominates* |
| Rwanda | **Maize** | Sorghum | 21.8 | 41.5 | -19.7 | 33.1 | 1.1 | *MUAC predominates* |
| Mozambique | **Maize** | Rice | 21.4 | 41.0 | -19.6 | 36.7 | 3.8 | *MUAC predominates* |
| Angola | **Maize** | Wheat | 23.2 | 42.6 | -19.4 | 37.6 | 4.9 | *MUAC predominates* |
| Uganda | **Maize** | Wheat | 24.1 | 42.8 | -18.6 | 24.4 | 3.2 | *MUAC predominates* |
| Afghanistan | **Wheat** | Rice | 23.0 | 41.3 | -18.3 | 44.6 | 3.6 | *MUAC predominates* |
| Zambia | **Maize** | Cassava | 27.4 | 44.6 | -17.2 | 34.6 | 4.2 | *MUAC predominates* |
| Sierra Leone | **Rice** | Cassava | 22.9 | 35.3 | -12.4 | 26.3 | 6.3 | *MUAC predominates* |
| Tanzania | **Maize** | Rice | 24.7 | 35.0 | -10.4 | 30.0 | 3.1 | *MUAC predominates* |
| DRC | **Cassava** | Maize | 32.0 | 40.8 | -8.8 | 44.7 | 7.2 | *MUAC predominates* |
| Eritrea | **Maize** | Wheat | 33.8 | 40.3 | -6.5 | 52.5 | 14.6 | *MUAC predominates* |
| Haiti | **Rice** | Wheat | 32.7 | 37.5 | -4.8 | 22.0 | 5.0 | *Near parity* |
| Madagascar | **Rice** | Wheat | 28.9 | 33.7 | -4.8 | 39.8 | 7.2 | *Near parity* |
| Ivory Coast | **Rice** | Wheat | 28.8 | 32.8 | -4.0 | 23.4 | 8.1 | *Near parity* |
| CAR | **Maize** | Cassava | 32.8 | 36.7 | -3.9 | 37.9 | 5.2 | *Near parity* |
| Liberia | **Rice** | Cassava | 32.4 | 30.8 | +1.6 | 29.8 | 3.4 | *Near parity* |
| Pakistan | **Wheat** | Rice | 36.1 | 33.6 | +2.6 | 37.6 | 7.1 | *Near parity* |
| Guinea | **Rice** | Wheat | 34.9 | 31.1 | +3.8 | 26.1 | 6.4 | *Near parity* |
| Myanmar | **Rice** | Wheat | 33.9 | 29.9 | +4.0 | 26.7 | 7.4 | *Near parity* |
| Zimbabwe | **Maize** | Wheat | 40.7 | 29.3 | +11.3 | 25.9 | 5.1 | *WHZ moderate predominance* |
| Nigeria | **Maize** | Sorghum | 38.6 | 27.0 | +11.6 | 33.8 | 11.6 | *WHZ moderate predominance* |
| Ethiopia | **Wheat** | Maize | 41.3 | 29.7 | +11.6 | 36.8 | 6.8 | *WHZ moderate predominance* |
| Nepal | **Rice** | Wheat | 38.8 | 26.2 | +12.6 | 24.8 | 7.0 | *WHZ moderate predominance* |
| Somalia^‡^ | **Wheat** | Rice | 47.3 | 26.4 | **+20.8** | 25.3 | 14.3 | *WHZ strongly predominates* |
| Bangladesh | **Rice** | Wheat | 46.6 | 24.9 | **+21.7** | 23.6 | 10.7 | *WHZ strongly predominates* |
| Niger | **Millet** | Sorghum | 43.5 | 21.0 | **+22.5** | 47.7 | 10.9 | *WHZ strongly predominates* |
| **Timor-Leste** | **Rice** | Maize | 51.0 | 20.5 | **+30.5** | 46.7 | 8.3 | *WHZ strongly predominates* |
| **Chad** | **Sorghum** | Millet | 49.9 | 18.2 | **+31.7** | 31.9 | 7.8 | *WHZ strongly predominates* |
| **Benin** | **Rice** | Maize | 51.5 | 19.0 | **+32.5** | 34.1 | 8.3 | *WHZ strongly predominates* |
| **South Sudan** | **Sorghum** | Maize | 51.4 | 18.4 | **+33.1** | 31.3 | 22.7 | *WHZ strongly predominates* |
| **Cameroun** | **Maize** | Wheat | 50.3 | 16.9 | **+33.4** | 28.9 | 4.3 | *WHZ strongly predominates* |
| **Mauritania** | **Wheat** | Rice | 58.7 | 22.1 | **+36.6** | 25.1 | 13.6 | *WHZ very strongly predominates* |
| **Sudan** | **Wheat** | Sorghum | 54.3 | 15.1 | **+39.1** | 38.2 | 16.3 | *WHZ very strongly predominates* |
| **Mali** | **Millet** | Rice | 55.5 | 16.3 | **+39.2** | 25.1 | 5.4 | *WHZ very strongly predominates* |
| **Burkina Faso** | **Sorghum** | Maize | 54.3 | 14.6 | **+39.7** | 21.1 | 9.3 | *WHZ very strongly predominates* |
| **Guinea-Bissau** | **Rice** | Wheat | 58.9 | 19.0 | **+39.9** | 28.1 | 5.1 | *WHZ very strongly predominates* |
| **India** | **Wheat** | Rice | 54.5 | 11.6 | **+42.8** | 35.5 | 18.7 | *WHZ very strongly predominates* |
| **Togo** | **Maize** | Sorghum | 63.4 | 13.1 | **+50.3** | 23.8 | 5.7 | *WHZ very strongly predominates* |
| **Kenya** | **Maize** | Wheat | 72.3 | 11.2 | **+61.1** | 17.6 | 4.5 | *WHZ very strongly predominates* |
| **Thailand** | **Rice** | Wheat | 71.7 | 10.1 | **+61.6** | 12.4 | 7.2 | *WHZ very strongly predominates* |
| **Gambia** | **Rice** | Wheat | 72.0 | 7.3 | **+64.8** | 17.5 | 5.1 | *WHZ very strongly predominates* |
| **Philippines** | **Rice** | Wheat | 75.5 | 7.4 | **+68.1** | 26.7 | 5.4 | *WHZ very strongly predominates* |
| **Sri Lanka** | **Rice** | Wheat | 76.5 | 7.8 | **+68.7** | 10.5 | 9.3 | *WHZ very strongly predominates* |
| **Senegal** | **Rice** | Wheat | 77.0 | 6.9 | **+70.1** | 17.5 | 10.2 | *WHZ very strongly predominates* |

^‡^ Somalia was excluded from the primary analysis on two independent grounds. First, FAOSTAT food balance sheet data for Somalia are currently unavailable and masked for data quality reasons (Filipczuk T, FAO Rome, personal communication, 17 November 2025). Second, the most recent JME wasting survey for Somalia dates from 2009—a 14-year temporal mismatch with the 2019–2023 FAOSTAT reference period, making it the oldest survey in the dataset. The staple values shown here are from an earlier FAOSTAT extraction (2018–2022) and indicate wheat as the dominant staple and rice as the second staple. However, the WHZ-strongly-predominates pattern shown above is more consistent with sorghum- and millet-consuming countries in the Sahel, suggesting possible underrepresentation of subsistence cereal consumption in the food balance data or a shift in dietary patterns since the 2009 survey.

**Panel B. Summary by dominant dietary staple (protein-based, n=47)**

| **Dom staple** | **WHZ only %** | **MUAC only %** | **WHZ−MUAC** | **N** | **Key observation** |
| --- | --- | --- | --- | --- | --- |
| **Cassava** | 27.6% | 42.0% | **-14.4** | 2 | *MUAC strongly predominates — DRC+Burundi; kwashiorkor and stunting denominator both operate; note cassava food volume substantially exceeds protein contribution in many additional Sub-Saharan African countries* |
| **Maize** | 34.3% | 35.6% | **-1.3** | 15 | *Near parity — Southern/Eastern Africa and Latin America; zinc-phytate stunting reduces WHZ denominator; Guatemala most MUAC-predominant (−40·2%)* |
| **Wheat** | 41.8% | 28.7% | **+13.0** | 8 | *WHZ moderate predominance — South Asia, MENA, Central Asia; Tajikistan outlier (−30·8) driven by high stunting* |
| **Rice** | 49.1% | 22.4% | **+26.7** | 17 | *WHZ moderate-strong predominance — SE Asia and West Africa; Timor-Leste (+30·5) driven by substantial maize second staple (7·12g/d)* |
| **Millet** | 49.5% | 18.6% | **+30.8** | 2 | *WHZ strongly predominates — Sahelian belt* |
| **Sorghum** | 51.9% | 17.1% | **+34.8** | 3 | *WHZ very strongly predominates — Sahelian core* |

**Panel C. Spearman correlations with WHZ−MUAC difference (n=47)**

| **Variable** | **Spearman r** | **p-value** | **Interpretation** |
| --- | --- | --- | --- |
| Stunting prevalence | **-0.459** | **0.001** | *Higher stunting → MUAC predominates (height denominator)* |
| Rice protein (g/d) | **+0.445** | **0.002** | *Higher rice → WHZ predominates* |
| Millet protein (g/d) | **+0.364** | **0.012** | *Higher millet → WHZ predominates* |
| Sorghum protein (g/d) | **+0.305** | **0.037** | *Higher sorghum → WHZ predominates* |
| Wheat protein (g/d) | +0.231 | 0.118 | *Not significant — positive trend in expected direction* |
| Maize protein (g/d) | -0.172 | 0.249 | *Not significant — positive trend in expected direction with MUAC predominance* |

*Colour coding — dominant staple: green=sorghum/millet; blue=maize; purple=cassava; yellow=rice; cream=wheat.*

*Notes: (1) Guatemala — maize dominant (16·36g/d), most MUAC-predominant country in entire dataset (−40·2%); WHZ-measured wasting 0·8% substantially underestimates true lean mass deficit. (2) Ivory Coast — rice dominant by protein but cassava food volume substantial; near parity (−4·0). (3) Tajikistan — wheat dominant, strongly MUAC-predominant (−30·8) driven by high stunting through height denominator. (4) Timor-Leste — rice dominant but maize second staple (7·12g/d) drives zinc-phytate stunting elevating WHZ (+30·5). (5) Cassava food volume substantially exceeds its protein supply contribution in many Sub-Saharan African countries — the FAOSTAT protein-based dominant staple classification therefore underestimates cassava dietary exposure in these settings, and the true cassava-associated MUAC predominance is likely larger than the n=2 cassava-dominant group suggests. (6) The WHZ-MUAC discordance pattern attributed to the dominant staple may be distorted by the mix of staples consumed within a country. Countries consuming two or more staples with opposing WHZ-MUAC effects — for example rice (WHZ-predominant) alongside cassava (MUAC-predominant) — may show intermediate discordance not reflecting either staple alone. The dominant staple classification represents the largest single protein contributor and does not capture the combined nutritional effect of the full staple mix. (7) Children identified by WHZ carry equivalent or higher mortality than those identified by MUAC (Grellety-Golden 2018); this analysis addresses differential geographic capture not clinical validity.*

**WHZ−MUAC discordance by dominant dietary staple: detection method breakdown (Grellety-Golden, n=46)**

A critical limitation of the primary analysis — and of global JME wasting estimates — is that wasting is defined exclusively by WHZ (<−2 SD), whereas MUAC-identified wasting represents a partially non-overlapping population. Grellety and Golden documented the percentage of wasted children identified by WHZ only, MUAC only, and both criteria simultaneously across 47 nationally representative surveys, permitting characterisation of the relative contribution of each detection method by dominant dietary staple. S Figure 13 presents the proportional breakdown of GAM detection across 46 surveys (Somalia excluded — absent from FAOSTAT dataset), grouped by primary dominant dietary staple.

To test whether WHZ−MUAC discordance differed systematically across staple groups, a Kruskal-Wallis test was applied to the 46 surveys classified into three groups: Millet/Sorghum (n=5), Maize/Cassava (n=17), and Rice/Wheat (n=24). The test indicated a statistically significant difference in discordance across the three groups (H=10·5, df=2, p=0·005). Post-hoc Mann-Whitney pairwise comparisons showed that Maize/Cassava-dominant countries had significantly lower WHZ−MUAC discordance — that is, a greater tendency toward MUAC-predominant wasting detection — than both Millet/Sorghum-dominant countries (U=72, p=0·019) and Rice/Wheat-dominant countries (U=96, p=0·004). Millet/Sorghum and Rice/Wheat countries did not differ significantly from each other (U=76, p=0·371), though this comparison was underpowered given only five Millet/Sorghum surveys in the dataset.

The direction of findings is consistent with the hypothesis: Millet/Sorghum countries showed WHZ-predominant wasting (median +33·1 pp); Maize/Cassava countries showed MUAC predominance (median −10·4 pp), with secondary staple patterns visible within this group; and Rice/Wheat countries were predominantly WHZ-predominant (median +17·1 pp). No correction for multiple comparisons was applied given the exploratory, hypothesis-generating nature of this analysis.

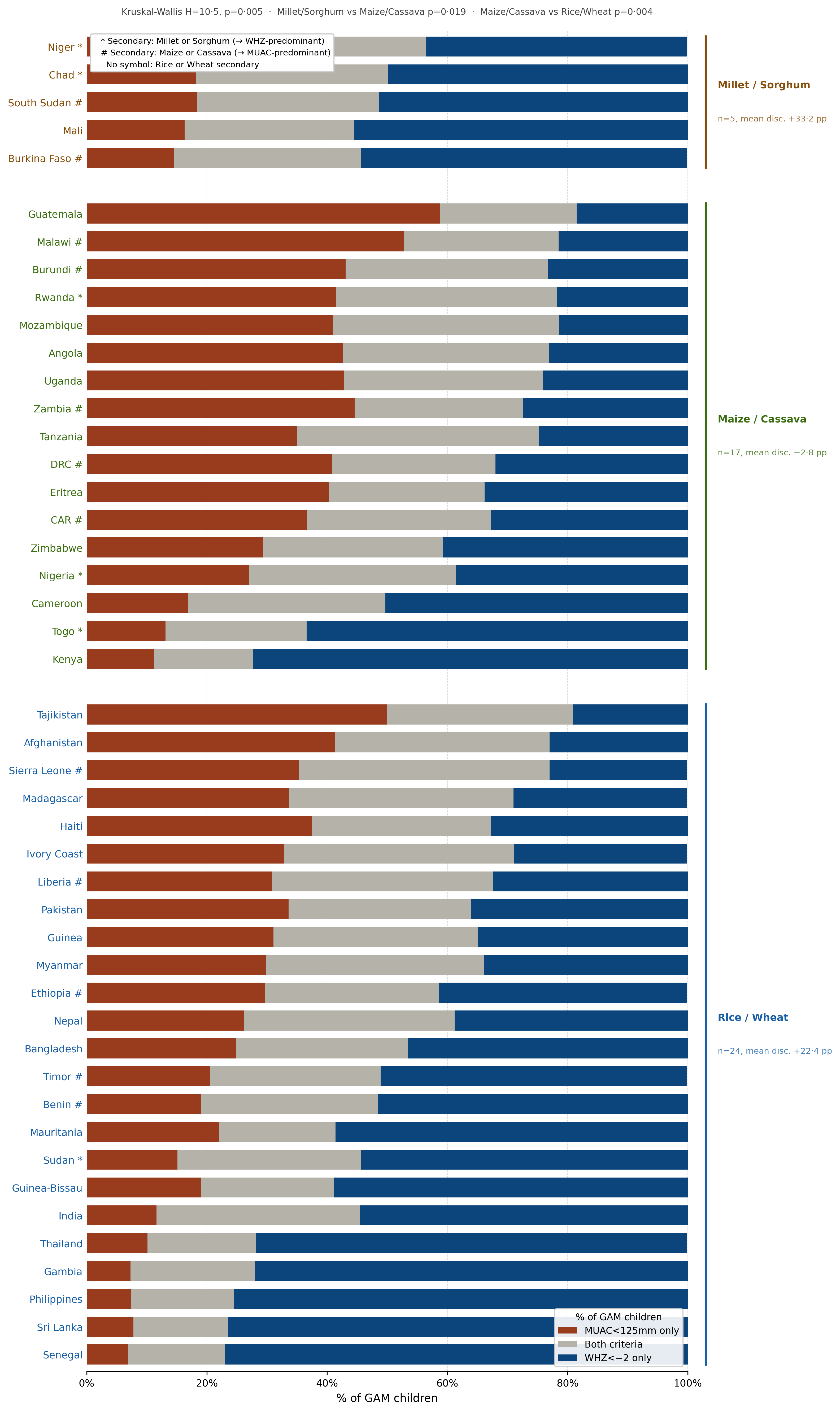

**S Figure 13.** *Proportion of GAM children detected by WHZ<−2 only, MUAC<125mm only, or both criteria simultaneously across 46 nationally representative surveys (Grellety E, Golden MH. BMC Nutrition 2016;2:10, Table S2). Countries grouped by primary dominant dietary staple protein supply (FAOSTAT 2019–2023 five-year mean) and sorted by WHZ−MUAC discordance (= %WHZ<−2 only minus %MUAC<125mm only) within each group. Somalia excluded (absent from FAOSTAT dataset; n=47→46). * Secondary staple is millet or sorghum (associated with WHZ predominance). # Secondary staple is maize or cassava (associated with MUAC predominance). No symbol = rice or wheat secondary staple. Kruskal-Wallis test across three groups: H=10·5, p=0·005. Mann-Whitney pairwise: Millet/Sorghum vs Maize/Cassava p=0·019; Maize/Cassava vs Rice/Wheat p=0·004; Millet/Sorghum vs Rice/Wheat p=0·371 (ns; n=5 limited power). Dominant staple = highest FAOSTAT protein supply contributor (g/capita/day).*

**Oedema prevalence by dominant dietary staple (Grellety-Golden cross-reference, n=42)**

Grellety and Golden reported the percentage of survey children excluded for oedema across 47 nationally representative DHS/MICS surveys. Oedema is a hallmark of kwashiorkor — oedematous protein-energy malnutrition characterised by bilateral pitting oedema — and is distinct from the lean mass depletion of marasmus. Children with oedema are excluded from WHZ-based wasting analyses because their true weight is artificially elevated by fluid retention, causing WHZ to underestimate true nutritional status. The oedema exclusion percentage therefore provides an indirect index of kwashiorkor burden in each survey population.

S Figure 14 shows oedema exclusion percentages by dominant dietary staple. Cassava-dominant countries had the highest mean oedema exclusion (Burundi 1·40%, DRC 1.12%), followed by maize-dominant countries (mean 0·67%; Mozambique 2·36%, Tanzania 1·40%, Rwanda 1·28%, Malawi 1·14%). Sorghum- and millet-dominant countries had consistently low oedema exclusion rates (sorghum mean 0·26%; millet mean 0·18%), as did rice-dominant countries (mean 0·19%). This pattern is consistent with the distinct malnutrition phenotypes proposed: cassava and maize protein quantity and quality deficit produces oedematous malnutrition (kwashiorkor), whereas sorghum and millet amino acid quality deficit produces lean mass depletion without oedema (marasmus or marasmic kwashiorkor). The higher oedema burden in cassava and maize populations further explains MUAC predominance in these countries — oedematous children have falsely elevated WHZ (oedema adds weight) but appropriately low MUAC (oedema does not affect mid-arm circumference), so MUAC detects oedematous wasting that WHZ misses entirely. These findings are theory-driven, ecological and hypothesis-generating.

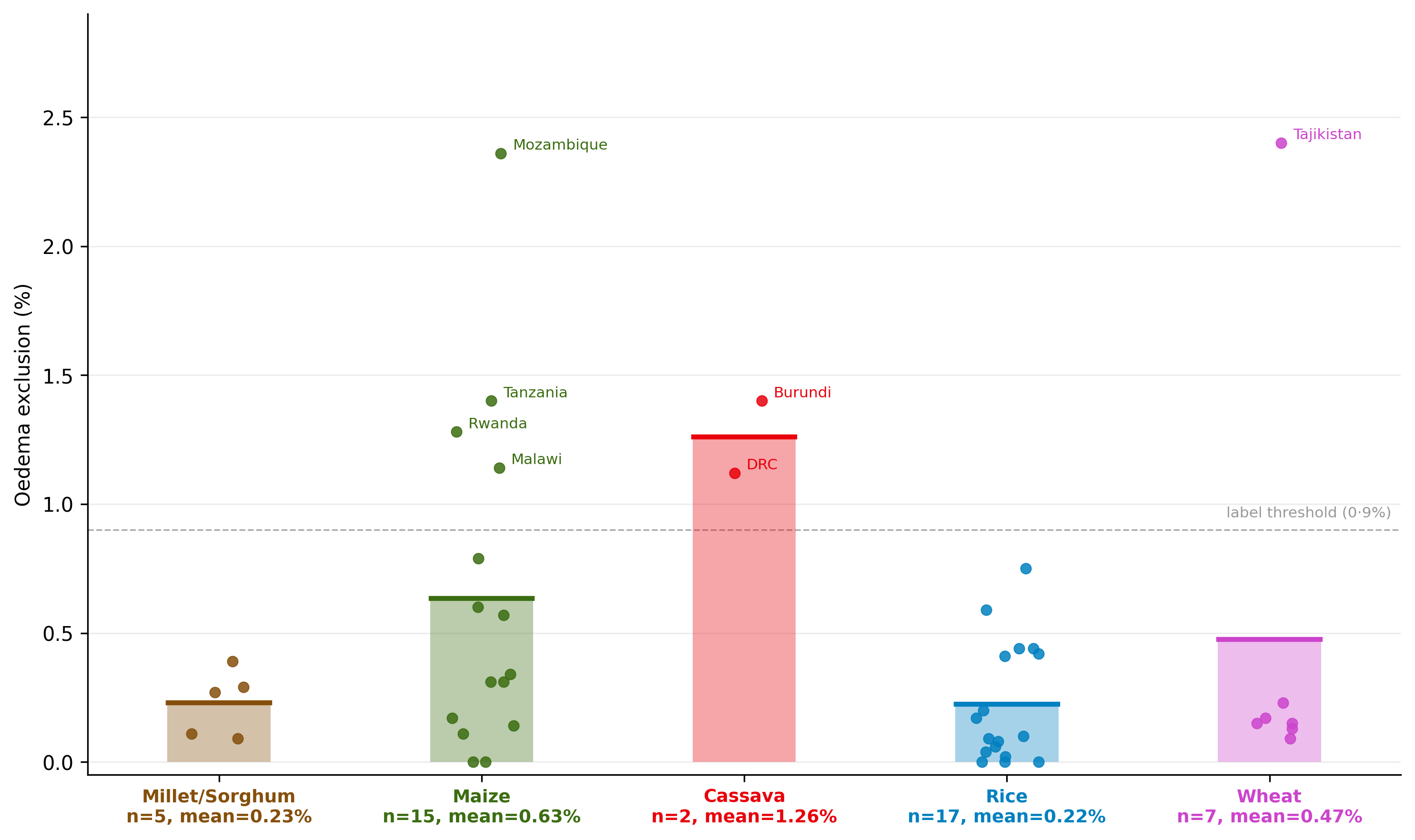

**S Figure 14.** *Oedema exclusion percentage by dominant dietary staple across 42 nationally representative surveys (Grellety and Golden, BMC Nutr 2016). Bars = group mean. Points = individual country surveys (jittered). Countries with oedema exclusion >0·9% labelled. Dominant staple assigned from FAOSTAT 2019–2023 protein supply (present study).*

**S Section F.2. WaSt in infants under 6 months by dietary staple (Kerac et al cross-reference, n=56)**

*Data arrangement — Kerac et al cross-reference: Country-level stunting, wasting and concurrent wasting-and-stunting (WaSt) prevalences in infants under 6 months were extracted from Kerac et al (BMJ Global Health 2025;10:e016121), which reported these for 56 LMIC DHS datasets. Each country was linked by name to FAOSTAT 2019–2023 five-year mean staple protein supply (g/capita/day) from our primary analysis dataset. All 56 countries were successfully matched. Dominant staple was defined as the staple with the highest per capita protein supply (g/capita/day) among wheat, rice, maize, millet, sorghum and cassava. The WaSt, stunting and wasting values used in all cross-reference analyses are exclusively from the Kerac et al dataset — not from our JME March 2025 master dataset — ensuring internal consistency with the Kerac survey-derived anthropometric data. Stunting values in the Grellety cross-reference similarly derive from Grellety’s own survey data, not from our JME March 2025 estimates. Animal protein supply from our primary FAOSTAT dataset was appended as a covariate for linear regression only. All analyses were conducted in R version 4·5·3 (lm() for regression; cor.test() method=‘spearman’ for correlations).*

Kerac et al (BMJ Global Health 2025;10:e016121) analysed anthropometric data from 56 LMIC DHS datasets in infants under 6 months of age.^30^ Since infants at this age have not yet begun complementary feeding, any association between country-level dietary staple type and malnutrition outcomes in this age group must operate more through maternal dietary protein quality transmitted in utero or via breast milk rather than the infant’s own food intake. We cross-referenced the Kerac dataset with FAOSTAT 2019–2023 five-year mean staple protein supply data for 56 countries present in both datasets to examine whether dominant dietary staple type predicts WaSt (concurrent wasting and stunting) prevalence in this critical early window.

**S Table 22. Summary of stunting, wasting and WaSt in infants under 6 months by dominant dietary staple (Kerac et al cross-reference)**

| **Dominant staple** | **n** | **Mean stunting <6m (%)** | **Mean wasting <6m (%)** | **Mean WaSt <6m (%)** | **Countries** |
| --- | --- | --- | --- | --- | --- |
| Sorghum | 2 | 11·0 | 20·8 | 1·80 | Burkina Faso, Chad |
| Millet | 2 | 14·8 | 15·1 | 1·85 | Mali, Niger |
| Wheat | 17 | 15·4 | 10·3 | 0·90 | Albania, Armenia, Congo, Egypt, Ethiopia, Gabon, India, Kyrgyz Republic, Maldives, Mauritania, Namibia, Pakistan, Papua New Guinea, Peru, Tajikistan, Türkiye, Yemen |
| Rice | 17 | 16·2 | 9·8 | 0·71 | Bangladesh, Benin, Cambodia, Comoros, Côte d’Ivoire, Dominican Republic, Gambia, Ghana, Guinea, Haiti, Liberia, Madagascar, Myanmar, Nepal, Senegal, Sierra Leone, Timor-Leste |
| Maize | 16 | 19·1 | 5·4 | 0·61 | Angola, Cameroon, Guatemala, Honduras, Kenya, Lesotho, Malawi, Mozambique, Nigeria, Rwanda, South Africa, Tanzania, Togo, Uganda, Zambia, Zimbabwe |
| Cassava | 2 | 22·0 | 8·1 | 0·90 | Burundi, DRC |

*Linear regression of WaSt on sorghum protein supply, millet protein supply and stunting prevalence in infants under 6 months showed directionally consistent positive associations for both sorghum (β=+0·062, p=0·252) and millet (β=+0·049, p=0·333), with stunting also showing a positive, statistically significant association (β=+0·042, p=0·012; overall model F p=0·005). Addition of animal protein as a covariate did not materially change the associations (sorghum β=+0·056, p=0·315; millet β=+0·049, p=0·336; stunting β=+0·040*, p=0·022; overall model F p=0·011). Full sample: n=56 countries.*

*Dominant staple = staple with highest per capita protein supply (g/capita/day) from FAOSTAT 2019–2023. Countries classified by dominant staple may include populations consuming other staples — see ecological misclassification note below.*

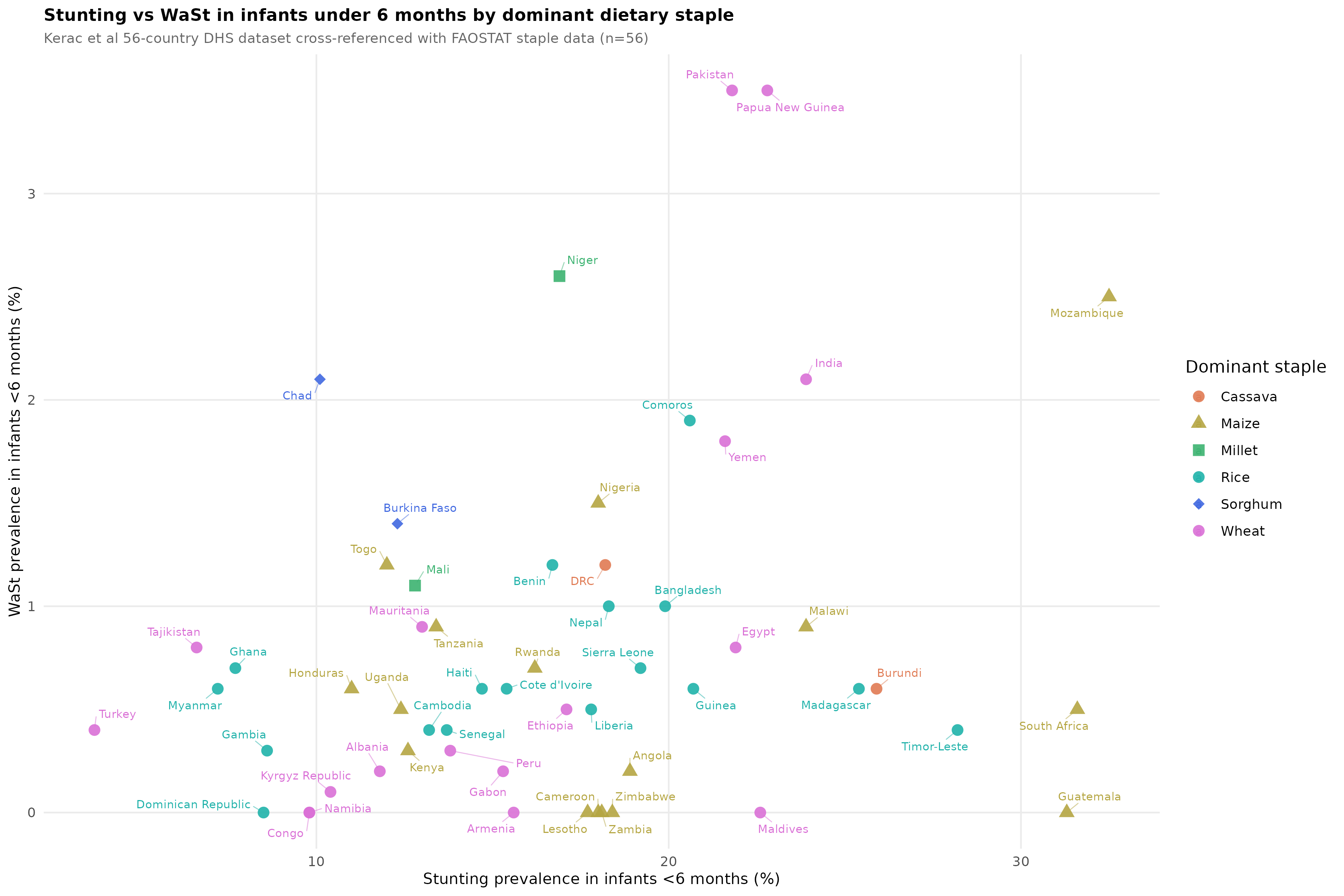

**S Figure 15: Stunting vs WaSt data cross-referenced with FAOSTAT data**

***S Figure 15. Stunting vs WaSt in infants under 6 months by dominant dietary staple (Kerac et al 56-country dataset cross-referenced with FAOSTAT staple data). Sorghum (blue) and millet (green) countries cluster at low stunting/high WaSt; cassava (salmon) and maize (olive) countries cluster at high stunting/low WaSt. Pakistan and Mozambique are outliers with high values on both axes.***

WaSt prevalence in infants under 6 months was more than double in sorghum- and millet-dominant countries (mean 1·80%) compared with cassava- and maize-dominant countries (mean 0·86%). Sorghum and millet dominant countries like Niger, Burkina Faso and Chad clustered in the upper-left quadrant of S Figure 15 — low stunting, high WaSt — while cassava and maize countries clustered in the lower-right — high stunting, low WaSt. This pattern is consistent with each staple driving a distinct growth phenotype determined by its specific nutritional profile: sorghum and millet protein quality deficit produces lean mass depletion and wasting despite relatively preserved linear growth; cassava protein quantity deficit preferentially impairs linear growth producing stunting; and maize-dominant populations show high stunting with suppressed WHZ-measured wasting consistent with the height denominator effect described in S Section F.1. Since infants under 6 months have not yet begun complementary feeding, this WaSt differential must reflect maternal dietary staple protein quality operating through pregnancy and lactation — suggesting the staple-specific growth phenotype is established from the earliest weeks of life. Pakistan is a notable outlier (stunting 21·8%, WaSt 3·5%) — possibly reflecting subnational variation from sorghum- and millet-consuming provinces rather than the national wheat-dominant diet. These analyses are theory-driven, ecological and hypothesis-generating.

Ecological misclassification note: Country-level dominant staple classification may misclassify large heterogeneous countries. India’s high WaSt likely reflects sorghum- and millet-consuming states (Maharashtra, Gujarat, Rajasthan) rather than rice-consuming coastal populations; Pakistan’s high WaSt may originate from sorghum- and millet-consuming provinces rather than the national wheat-dominant diet; Madagascar’s cassava classification could reflect southern regions rather than the rice-dominant north. This ecological misclassification biases the analysis toward the null — the true staple-WaSt association may be stronger than observed at country level. Subnational analysis linking district-level dietary staple data with DHS anthropometric surveys would be required to confirm this. Furthermore, country-level ecological analysis cannot account for genetic and epigenetic contributors to growth phenotype. The thin-fat Indian baby phenotype — characterised by low lean mass and relatively preserved adiposity at birth, documented by Yajnik and others — may independently contribute to high WaSt in India through intergenerational epigenetic programming of body composition, operating in parallel with or independently of dietary staple protein quality.^31,32^

**S Section F.3. Post-hoc cross-reference: Maternal short stature and thinness by dietary staple protein supply (DHS StatCompilerence, n=57)**

*Data arrangement — DHS cross-reference: Country-level prevalence of maternal short stature (height <145cm) and thinness (BMI<18·5) among women of reproductive age were extracted from the DHS Program StatCompiler (ICF, 2025; statcompiler.com), using the most recent available survey for each country as of June 2026. These DHS indicators were linked by country name to staple protein supply (g/capita/day, FAOSTAT 2019–2023 five-year means), total dietary energy availability (kcal/capita/day) and animal protein supply (g/capita/day) from our primary analysis dataset. Of 58 countries with DHS short-stature and thinness data available, 56 were matched to our primary dataset by country name (with manual correction for non-standard naming, e.g. “Congo Democratic Republic” for the Democratic Republic of the Congo); Turkey (DHS StatCompiler name) was matched to Türkiye (ISO 3166 official name) in our primary dataset by manual correction; dietary covariates were successfully linked. The final analytical sample was n=57 after exclusion of one further country with incomplete dietary covariate data. As in the Kerac and Grellety-Golden cross-references above, the maternal anthropometric values used here are exclusively from DHS StatCompiler, not from our JME March 2025 master dataset, and reflect women of reproductive age generally rather than specifically the mothers of children captured in the primary wasting and stunting analysis.*

This cross-reference was motivated by our results of maize being negatively associated with wasting in Primary analysis and the results are available in S Table 23. We wanted to establish whether this was linked to height-denominator mechanism or a true negative association with wasting. The height denominator effect is described in S Section F.1: because both WHZ and maternal body mass index (BMI) are computed as weight divided by a height-derived denominator, shorter stature could, in principle, mechanically inflate BMI for a given true body weight in the same way it attenuates WHZ for a given true lean-mass deficit. We have evidence from literature that maternal short stature is linked to child stunting.^33^ Similarly, maternal low BMI has been linked to both child wasting and stunting.^34^ So we reasoned that, staple-driven differences in zinc bioavailability and protein quality contribute to population-level differences in adult female stature, and also contribute to staple-associated differences in maternal BMI. We tested this using robust MM-estimation (robustbase, lmrob, KS2014 setting), consistent with the primary analysis, regressing each maternal anthropometric outcome on each staple’s protein supply (one staple per model, as in the primary analysis), with total dietary energy availability (kcal/capita/day) and animal protein supply (g/capita/day) included as covariates to test whether any staple association was attributable to overall energy or protein availability rather than to the staple itself.

For maternal short stature, no staple showed a robust, consistent association: rice showed a significant positive association (β=+0·109, p=0·007), the opposite direction to what the height-denominator hypothesis would predict for a WHZ-predominant staple; sorghum showed a borderline negative association (β=−0·175, p=0·073); wheat, maize, millet and cassava showed no significant association (S Table 24). For maternal thinness, by contrast, maize showed a strong, highly significant negative association (β=−0·422, p<0·001), indicating higher maize protein supply is associated with substantially lower prevalence of maternal thinness; sorghum and millet showed borderline positive associations (β=+0·528, p=0·066 and β=+0·396, p=0·086 respectively), indicating a trend toward higher maternal thinness with greater sorghum or millet supply. These associations for maize, sorghum and millet were materially unchanged by adjustment for total kcal/d and animal protein supply, both of which were themselves independently associated with lower maternal thinness in several models (e.g. animal protein β=−0·139, p=0·010 in the maize model).

This pattern argues against the height-denominator mechanism as the primary explanation for maize’s favourable maternal BMI profile. If shorter maternal stature in maize-dominant countries were mechanically inflating BMI and thereby masking true maternal undernutrition, the maize–thinness association would be expected to attenuate once stature-related confounding is addressed; instead, maize’s association with lower thinness was essentially unchanged in magnitude across unadjusted, kcal-adjusted and kcal-plus-animal-protein-adjusted models, and maize showed no significant association with short stature itself. The maize–thinness association is therefore more plausibly a genuine, independent nutritional association — consistent with maize’s comparatively higher DIAAS (~36) relative to sorghum (~29) and its broadly intermediate protein quality among the six staples (S Table 21) — than a measurement artefact of stature. This finding is consistent with, and lends independent corroboration to, maize’s robust negative (protective) association with child wasting in the primary analysis and across nearly all sensitivity specifications (S Table 13, Panel A), suggesting maize’s apparent protective effect against wasting in both mothers and children may share a common underlying nutritional, rather than purely artefactual, basis.

A secondary contribution of the height-denominator mechanism to maize’s wasting coefficient cannot be excluded, given maize’s high phytate:zinc ratio (45·5) and positive stunting associations in several specifications (S Table 12, Panel B); however, this is unquantified and the DIAAS-related component is more directly demonstrated. Because this is an ecological cross-sectional comparison, long-run genetic and epigenetic population differences in stature cannot be separated from current dietary effects — a limitation already noted above.

**S Table 23. Maternal short stature and thinness by staple protein supply, adjusted for total kcal/d and animal protein supply (robust MM-estimation, n=57)**

β = unstandardised coefficient per g/capita/day staple protein supply, from a model also including total kcal/d and animal protein supply (g/capita/day). Significance: ***p<0·001, **p<0·01, *p<0·05, †p<0·10. Outcomes are DHS StatCompiler indicators (most recent survey per country); see data arrangement note above.

| **Outcome** | **Staple** | **β (staple)** | **p (staple)** | **β (kcal)** | **p (kcal)** | **β (animal protein)** | **p (animal protein)** | **R²** |
| --- | --- | --- | --- | --- | --- | --- | --- | --- |
| Short stature (% <145cm) | Wheat | −0·037 | 0·165 | −0·0008 | 0·361 | +0·020 | 0·355 | 0·065 |
| Short stature (% <145cm) | Rice | +0·109 | 0·007 ** | −0·0022 | 0·028 * | +0·036 | 0·147 | 0·169 |
| Short stature (% <145cm) | Maize | −0·033 | 0·483 | −0·0014 | 0·121 | +0·009 | 0·685 | 0·054 |
| Short stature (% <145cm) | Millet | −0·122 | 0·112 | −0·0010 | 0·243 | −0·001 | 0·973 | 0·083 |
| Short stature (% <145cm) | Sorghum | −0·175 | 0·073 † | −0·0010 | 0·223 | −0·004 | 0·847 | 0·094 |
| Short stature (% <145cm) | Cassava | +0·180 | 0·138 | −0·0009 | 0·216 | +0·015 | 0·443 | 0·073 |
| Thinness (% BMI<18·5) | Wheat | +0·001 | 0·991 | −0·0031 | 0·230 | −0·101 | 0·116 | 0·200 |
| Thinness (% BMI<18·5) | Rice | +0·107 | 0·289 | −0·0037 | 0·139 | −0·078 | 0·211 | 0·215 |
| Thinness (% BMI<18·5) | Maize | −0·422 | 0·0003 *** | −0·0034 | 0·110 | −0·139 | 0·010 * | 0·388 |
| Thinness (% BMI<18·5) | Millet | +0·396 | 0·086 † | −0·0039 | 0·111 | −0·065 | 0·287 | 0·244 |
| Thinness (% BMI<18·5) | Sorghum | +0·528 | 0·066 † | −0·0035 | 0·143 | −0·065 | 0·292 | 0·253 |
| Thinness (% BMI<18·5) | Cassava | +0·252 | 0·540 | −0·0028 | 0·256 | −0·089 | 0·153 | 0·203 |

*Limitations specific to this cross-reference: DHS coverage was sparse and uneven across staples (e.g. only 1–2 of the ten highest sorghum- or millet-consuming countries in S Tables 15–16 had available DHS data), and several Sahelian countries central to the primary sorghum and millet wasting findings — Sudan, Burkina Faso and South Sudan — lacked DHS maternal anthropometric data entirely. This analysis should be regarded as exploratory and hypothesis-generating rather than confirmatory, in keeping with the other secondary cross-reference analyses in this section.*

S Table 24 below summarises the (hypothetical) effects of staples on child anthropometry in light of our analysis.

#### **S Table 24. Summary of nutritional profile and anthropometric associations across six dietary staples**

| **Staple** | **DIAAS** | **Protein (g/100g)** | **Zinc (mg/100g)** | **Phytate:Zn** | **Wasting (primary)** | **Overall anthropometric summary** |
| --- | --- | --- | --- | --- | --- | --- |
| **Wheat** | 48 good | 12·2 high | 2·6 ok | 29·6 high | −0·028 NS | Negative or null association with wasting and stunting throughout. Favourable protein quality and quantity. Despite an elevated phytate:zinc ratio (29·6), wheat shows no adverse anthropometric signal. |
| **Rice** | 47 good | 6·7 low | 1·1 low | 20·1 low | +0·086** | Positive wasting association, robust across S1/S2/S5. Negative stunting trend (S3/S5 significant), likely via animal-protein (fish) complementarity in South/South-East Asian diets. Stature relatively preserved, so lean-mass depletion is captured by WHZ — WHZ-predominant in Grellety cross-reference. |
| **Maize** | 36 ok | 9·5 ok | 1·8 ok | 45·5 high | −0·139** | Negative wasting, robust except in S3. Positive stunting trend (significant in S5; Q0·25 stunting QR). Consistent negative wasting association in primary and S1/S2/S4/S5. The negative WHZ-based association is consistent with MUAC predominance in Grellety cross-reference (73% of maize-dominant countries) and by a highly significant negative association between maize protein supply and BMI-for-age in the post-hoc cross-reference. HIV geographic confounding documented (S Table 21 Panel E); S4 reversal confirms artefactual HIV coefficient. Positive stunting in S5 and QR Q0·25. Likely protein quality issue than zinc-phytate(see discussion-main manuscript). |
| **Millet** | 7–68 wide range | 0·9–9·7 wide range | 2·5 ok | 27·0 high | +0·197 NS | No significant wasting association. Stunting direction inconsistent across analyses (positive at stunting QR Q0·25, negative in S5). FAOSTAT’s undifferentiated millet variable spans species with very different DIAAS (finger millet ~40 vs proso/foxtail 7–10), likely masking a true effect. Net signal unresolved. |
| **Sorghum** | 29 poor | 10·1 ok | 1·8 ok | 48·9 highest | +0·435*** | Strongest and most consistent positive wasting association of all six staples — significant in primary and all five sensitivity analyses including S3. Worst combined DIAAS/phytate:zinc profile. WHZ-predominant in Grellety/Leidman, consistent with true lean-mass depletion. Also positive for stunting in S5 — a "double burden" profile. |
| **Cassava** | 17–37 very poor | 0·9–2·1 lowest | 0·3 lowest | 16·9 low | +0·039 NS | No significant wasting association except positive in S5. Significant positive stunting in S1 and S5. Pattern resembles maize (low wasting via WHZ attenuation from stunting); cassava has both the lowest protein content and lowest zinc content of all six staples, so the relative contributions of protein-energy deficit versus zinc bioavailability to its stunting association cannot be disentangled. Consistent with maize, cassava-dominant countries show MUAC-predominant wasting in the Grellety & Golden cross-reference (−8·8) and Leidman (−1·6), supporting the interpretation that WHZ underestimates true wasting burden in cassava-dominant populations likely due to height denominator effect. |

*Significance: ***p<0·001, **p<0·01, *p<0·05, NS = not significant. Wasting column shows primary MM-estimation coefficients (n=127). The summary column synthesises primary, sensitivity (S1–S5), quantile regression and secondary cross-reference (Grellety & Golden, Kerac) findings.*

**Bibliography — References for Supplementary Sections**
